# Genetic regulation of the vascular endothelial growth factor receptor 1 during sepsis and association with ARDS susceptibility

**DOI:** 10.1101/2025.01.30.25321087

**Authors:** Eva Suarez-Pajes, Nick Shrine, Eva Tosco-Herrera, Tamara Hernandez-Beeftink, Luis A. Rubio-Rodríguez, M. Isabel García-Laorden, Almudena Corrales, Miryam Prieto-González, Aurelio Rodríguez-Pérez, Demetrio Carriedo, Jesús Blanco, Alfonso Ambrós, Elena González-Higueras, Elena Espinosa, Arturo Muriel-Bombin, David Domínguez, Abelardo García de Lorenzo, José M. Añon, Marina Soro, Jesús Villar, Martin D Tobin, Louise V. Wain, Olivia C. Leavy, Beatriz Guillen-Guio, Carlos Flores

## Abstract

**Background:** Acute respiratory distress syndrome (ARDS) is associated with high mortality in Intensive Care Units (ICU). A previous genome-wide association study (GWAS) identified the vascular endothelial growth factor receptor 1 (*VEGFR1*) gene in ARDS risk. We performed a GWAS on soluble VEGFR1 (sVEGFR1) levels to identify protein quantitative trait loci (pQTLs) and genes of interest for ARDS.

**Methods:** Serum samples (n=292) within the first 24 (T1), 72 hours (T2), and 7 days (T7) after sepsis diagnosis were collected while in the ICU. sVEGFR1 levels were measured and tested for association. We combined fine mapping, colocalisation and gene-set mapping analyses to prioritise genes and test low-frequency variation association with ARDS (n=822). We analysed the association of sVEGFR1 pQTLs with mortality and ARDS susceptibility using polygenic scores (PGS). Finally, the causality of VEGFR1 in ARDS was assessed using two-sample Mendelian randomisation (MR) analyses.

**Results:** We found a pQTL for T2 sVEGFR1 levels at *TCF20* (rs134871, *p*=4.66×10^-8^). Fine mapping prioritised rs762995 as a likely pathogenic variant and *CYP2D6* as the most likely functional gene. The locus colocalised with eQTLs for *TCF20* and *CYP2D6*. Low-frequency missense variation in *TCF20* was associated with sepsis-associated ARDS susceptibility (*p*=3.0×10^-3^). sVEGFR1 levels PGS were associated with decreased ARDS susceptibility and mortality. MR analyses did not evidence causality.

**Conclusions:** We identified biologically relevant pQTLs of VEGFR1 levels during sepsis in *TCF20* and identified *CYP2D6* as the gene more biologically implicated. Low frequency missense variation in *TCF20* and sVEGFR1 levels PGS models were associated with sepsis outcomes.

**What is already known on this topic:** Acute respiratory distress syndrome (ARDS) is an acute condition, characterised by respiratory failure, an acute inflammatory response and the development of non-cardiogenic oedema. There is a need for target pharmacological strategies and advances in the risk stratification methods that can improve patient management.

**What this study adds:** We performed the first GWAS of sVEGFR1 levels in patients with sepsis. The integration of different complementary genomic approaches has allowed us to reveal a regulatory variant of sVEGFR1 during sepsis, suggesting a role in ARDS susceptibility and mortality. In exome-wide low-frequency variation analyses, we identified *TCF20* as a novel gene of interest for ARDS.

**How this study might affect research, practice or policy:** This study emphasises the value of proteogenomic approaches in improving our understanding of ARDS pathogenesis and detecting novel risk genes to advance patient risk stratification.

## Introduction

Acute respiratory distress syndrome (ARDS) is a critical lung condition characterised by diffuse alveolar damage, bilateral pulmonary infiltrates, and acute hypoxaemia. The syndrome is heterogeneous and can be triggered by pulmonary and non-pulmonary insults, including sepsis, pneumonia, transfusion, aspiration, or trauma (The ARDS Definition Task Force*, 2012; Grasselli et al., 2023). These insults generate a dysregulated inflammatory response that affects the alveolar-capillary barrier and gas exchange, leading to the formation of non-cardiogenic oedema (Bos & Ware, 2022; Grasselli et al., 2023).

The current diagnosis of ARDS lacks specific tests and relies on clinical criteria such as physical examination, chest imaging, and oxygenation levels (Grasselli et al., 2023; The ARDS Definition Task Force*, 2012). Patient management involves life support strategies aimed at promoting oxygenation and reducing the inflammatory response, using mechanical ventilation, prone positioning, or corticosteroid administration (Gorman et al., 2022). Patients with ARDS require admission to intensive care units (ICUs) and have a high mortality rate of approximately 40%. Survivors commonly suffer physical and cognitive sequelae (Bellani et al., 2016; Gorman et al., 2022; Hendrickson et al., 2021).

Genetic studies of ARDS aim to understand disease susceptibility and identify novel pharmacological strategies and drug repositioning (Giannini & Meyer, 2021; Suarez-Pajes et al., 2023). A previous genome-wide association study (GWAS) of ARDS allowed us to identify a significant risk locus at the *FLT1* gene, encoding the vascular endothelial growth factor receptor 1 (VEGFR1) (Guillen-Guio et al., 2020). Results also revealed that blood *FLT1* gene expression was higher in ARDS patients than in other critically ill patients. VEGFR1 is a transmembrane tyrosine kinase receptor and one of the main ligands of VEGFA (also known as VEGF), which is critically involved in angiogenesis and vascular permeability (Shibuya, 2011). In addition, the soluble form of VEGFR1 (sVEGFR1), which is encoded as a splicing variant, appears to act as an antagonist by sequestering VEGFA and reducing its bioavailability (Failla et al., 2018; Saito et al., 2013).

The identification of biomarkers for ARDS aims to enhance diagnosis, predict mortality, and classify patients (Jabaudon et al., 2021; Villar et al., 2021). In this sense, circulating proteins are a valuable resource for understanding health status and disease progression. Large-scale initiatives have been undertaken to study the protein genomic regulation and identify protein quantitative trait loci (pQTLs). Proteogenomic-focused studies and statistical approaches such as Mendelian randomisation (MR) allow the use of genetic variants as instrumental variables to assess causality between different proteins and traits (Eldjarn et al., 2023; Sun et al., 2023; Zhu et al., 2024). This approach has been used to assess the causal effect of different proteins on ARDS susceptibility and outcomes (Dong et al., 2021; Jones et al., 2020; Reilly et al., 2018).

The biological context plays a critical role on genetic regulation, implying that many regulatory variants may be detectable only under specific conditions (Mostafavi et al., 2023) (Burnham et al., 2024). Therefore, here we performed a GWAS of sVEGFR1 protein levels after sepsis diagnosis to identify pQTLs that may be involved in the pathophysiology of ARDS, followed by complementary analyses including low-frequency variant analyses to identify genes of interest. Finally, we investigated the causal effect of VEGFR1 and VEGFA in sepsis-associated ARDS using MR.

## Material and methods

### Study design and sample description

We included serum samples taken at different times after sepsis diagnosis (**Figure 1**) from 292 patients from the GEN-SEP cohort, an observational, multicentre study conducted in Spanish ICUs. The different collection intervals were: i) within 24 hours after diagnosis (T1), ii) within 48-72 hours after diagnosis (T2), and iii) 7 days after diagnosis (T7). Further details of the study description are available in the **supplementary material**.

**Figure 1.**
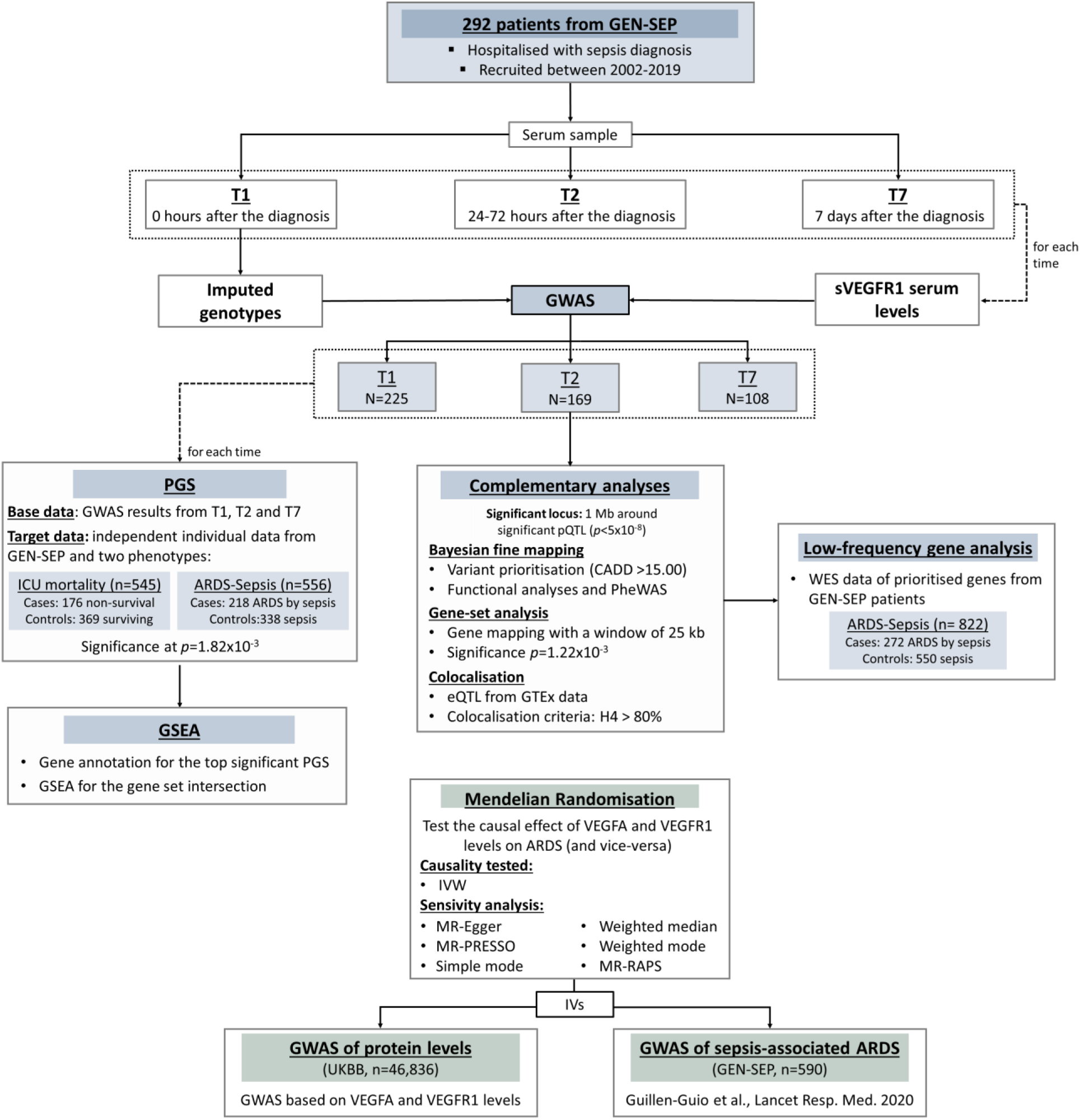
Flowchart of the study design. CADD: combined annotation dependent depletion score; eQTL: expression quantitative trait locus; GSEA: gene-set enrichment analysis; GWAS: genome-wide association study; IV: instrumental variable; ICU: intensive care unit; IVW: inverse variance-weighted; PGS: polygenic score; UKBB:UK biobank; WES: whole-exome sequencing.

Informed consent was obtained from all the patients. The study was performed according to The Code of Ethics of the World Medical Association (Declaration of Helsinki) and approved by the Ethics Committee for Drug Research from the Hospital Universitario de Canarias (Code: CHUNSC_2021-40).

### Serum VEGFR1 quantification

sVEGFR1 levels in serum samples were quantified using the VEGFR1/Flt-1 DuoSet ELISA kit and the DuoSet Ancillary Reagent kit2 (R&D Systems) according to the manufacturer’s instructions. Outliers and samples with concentrations below the limit of detection (62.5 pg/ml) were excluded from the analyses. Serum concentrations were normalised by Box and Cox transformations and normality of the data was assessed using the Lilliefors test.

### Single nucleotide polymorphism (SNP) level associations

We used genetic data of patients with sepsis from a previous published study (Hernandez-Beeftink et al., 2022). Genotyping, quality controls, and imputation procedures are described in the **supplementary material**. To identify pQTLs, we tested the association of the imputed genotypes with sVEGFR1 levels at each of the three collection intervals using a linear Wald test implemented in EPACTS v3.2.6. We included sex, age, and the Acute Physiology and Chronic Health Evaluation II (APACHE II) score as covariates following the previous model (Guillen-Guio et al., 2020). SNPs with a minor allele frequency (MAF) of less than 1% or with low imputation quality (Rsq<0.3) were excluded from the analyses. Significance was declared at the genome-wide threshold (*p*<5×10^-8^). The genomic inflation factor (λ) was calculated with the *gap* package for R (v3.6.0) to assess the inflation of results.

### Bayesian fine mapping, functional prioritisation, and colocalisation

We further explored the 1 Mb window around the significant variant to identify SNPs and genes of interest. We applied Bayesian fine mapping to identify the credible set of variants most likely to harbour the causal variant with 95% confidence. We included variants showing r^2^>0.1, based on GEN-SEP data, and the posterior probabilities (PP) were calculated using *corrcoverage* (Hutchinson et al., 2020), assuming a single causal variant per locus. We annotated using the Combined Annotation Dependent Depletion (CADD) score v1.6 to predict the biological impact of the variants that were included in the credible set. We performed further *in silico* analyses for the significant and prioritised pQTLs (CADD>15), and their regulatory role was evaluated using RegulomeDB v2.2 (Boyle et al., 2012) and Haploreg v4.2 (Ward & Kellis, 2012). We prioritised genes based on the functional evidence integrated into the Variant-to-Gene (V2G) score from Open Target Genetics (Ghoussaini et al., 2021). We conducted a phenome-wide association study (PheWAS) using data from Open Targets (Ghoussaini et al., 2021) to identify whether the significant and prioritised variant had previously been associated with phenotypes of interest with a *p*<0.005. We also performed a gene-set analysis mapping of the region using MAGMA v1.08, implemented in FUMA v1.5.2 (https://fuma.ctglab.nl/), to assist in variant-to-gene mapping for identifying the gene that most likely explain the signal. SNP localisation was used to map the association results to the genes in the region using a window size of 25 kb on each side.

We assessed the colocalisation between the significant locus and expression quantitative trait loci (eQTLs) data from GTEX v8 (https://gtexportal.org/) to detect whether the identified pQTLs also regulate the expression of other genes. For this, we selected the genes for which the significant variant is an eQTL (*p*<0.05) and evaluated the colocalisation using *coloc* (Giambartolomei et al., 2014) to assess the following hypotheses: H0: variants showed no association with sVEGFR1 levels, nor with eQTL data; H1: variants were only associated with sVEGFR1 levels; H2: variants were only associated with gene expression; H3: variants were associated with sVEGFR1 levels and gene expression, but by different causal SNP; H4: variants were associated with sVEGFR1 levels and gene expression by the same causal variant. We declared colocalisation when the PP of H4 was > 80%.

### Gene-based low frequency variant analysis on prioritised genes

We evaluated the gene-based association of low-frequency variants in the prioritised genes with sepsis-associated ARDS susceptibility. We accessed whole-exome sequencing (WES) data from 822 of the GEN-SEP patients, 272 cases with sepsis-associated ARDS and 550 sepsis patients defined as at-risk controls. We tested association using the optimal sequence kernel association test (SKAT-O) with Regenie v3.2.7 (Mbatchou et al., 2021). We tested three SNP categories: (i) variants with allele frequency (AF) <0.01; (ii) variants predicted as missense with AF<0.01; (iii) variants with AF<0.01 and CADD>15. Whenever a significant association was declared (*p*<0.008; corresponding to a Bonferroni correction for two genes by three SNP categories), a logistic regression in R (v4.0.0) was then used to estimate the effect of aggregated rare variation on ARDS susceptibility. Detailed information for sequencing procedures, variant calling, and annotation is available in the **supplementary material**.

### Polygenic score models

We developed and assessed polygenic scores (PGS) to determine whether sVEGFR1 pQTLs were associated with sepsis aggravation, specifically with ICU mortality or ARDS susceptibility. PRSice v2.3.5 (Choi & O’Reilly, 2019) was used to screen multiple PGS models by applying different *p*-value thresholds. We used the results of each collection interval GWAS as a base data. These PGS models were tested in a target cohort consisting of an independent subset of GEN-SEP patients to evaluate their association with susceptibility to sepsis-associated ARDS (218 cases and 338 controls) and ICU mortality (176 cases and 369 controls). Significance was declared at *p*=8.33×10^-3^ to correct for the number of independent tests. Details about the PGS procedures can be found in the **supplementary material**.

For the best significant fitting PGS, we extracted the SNPs included in the models and linked them to the annotated genes through functional regulatory data using GREAT (Tanigawa et al., 2022). Following this, in the common gene set, we performed gene set enrichment analysis using *enrichR* (Chen et al., 2013) was then used to assess whether there was an overrepresentation of genes in biological pathways of interest (based on Reactome 2022), in sepsis-related traits (based on the GWAS Catalog 2023), and in drug-related gene signatures (based on DSigDB).

### Mendelian Randomisation

To evaluate the causal effects of VEGFR1 levels on risk of sepsis-associated ARDS, we performed two-sample bidirectional MR analyses (**Figure S1**). We accessed UK Biobank (UKBB) data from 46,836 individuals from the general population with measurements of global VEGFR1 levels (accounting for the soluble and the membrane-bound forms). A GWAS was performed to determine the pQTLs associated with VEGFR1 levels in this cohort. We also assessed the causal effect of VEGFA (n=47,498) due to its direct binding to VEGFR1, its association with ARDS risk (Guillen-Guio et al., 2020), and its well-known role in lung permeability and pulmonary oedema (Lin et al., 2019; Tomita et al., 2020).

We evaluated causality using the inverse variance-weighted (IVW) estimate method and performed sensitivity analyses with five other methods, based on strong (F-stat>10) instrumental variables (IV) that met genome-wide significance (*p*<5×10^-8^) in the UKBB GWAS of circulating VEGFA and VEGFR1 protein levels, separately. For the opposite direction, we selected strong IVs that reached the suggestive threshold (*p*<5×10^-6^) declared at the discovery stage in our previous GWAS of sepsis-associated ARDS (Guillen-Guio et al., 2020). We evaluated the absence of pleiotropy by calculating the Q-statistic and we used MR-PRESSO to control for the effect of outliers. In addition, we used MR-RAPS to account for weak instruments (GWAS *p*<0.05) and pleiotropy. Further details on the UKBB GWAS and MR analyses can be found in the **supplementary material**.

## Results

After quality controls, serum sVEGFR1 levels from a total of 241 patients were obtained and normalised (**Figure S2**): 225 for T1, 169 for T2, and 108 for T7. Demographic and clinical features of the GEN-SEP patients retained for the study are available in the **supplementary material** (**Table S1**). Protein levels showed an increase over time (**Figure S3**), surpassing the statistical significance (*p*<0.05) when comparing T1 with T2 (*p*=4.28×10^-6^) or T1 with T7 (*p*=1.9×10^-7^) measurements. No significant differences were found between T2 and T7 (*p*=0.11).

In the GWAS of sVEGFR1 protein levels, there was no evidence of genomic inflation in the results (**Figure S4**). We found one pQTL at T2, located on chromosome 22, surpassing the statistical significance threshold (rs134871, *p*=4.66×10^-8^, beta= −0.23, standard error [SE]=0.04) (**Table 1**, **Figure 2**, **Figure 3**). The pQTL variant is intronic to the transcription factor 20 (*TCF20*) gene and the result was robust to model adjustments by covariates (**Table S2**). It also showed nominal significance in the GWAS of T1 (*p*=4.43×10^-3^) and of T7 (*p*=9.20×10^-3^), but was not associated with sepsis-induced ARDS (*p*=0.722) (Guillen-Guio et al., 2020) (**Table S3**). No genome-wide significant pQTLs were found for the other collection times. The most significant signal for T1 was located on chromosome 3 (rs59679804, *p*=1.78×10^-7^, beta= −0.16, SE=0.03), intergenic to ATP binding cassette subfamily C member 5 (*ABCC5*) and presenilin associated rhomboid like (*PARL*) genes (**Table 1, Figure S5**). This SNP was previously associated with blood cell traits, such as the monocyte percentage (*p*=4.2×10^-14^), mean platelet volume (*p*=1.2×10^-11^), platelet distribution width (PDW) (*p*=4.4 x10^-9^), and white blood cell count (*p*=3.9×10^-10^). For T7, the most significant association was found for rs72798674 (*p*=1.52×10^-6^, beta= −0.58, SE=0.11), which is intronic to the WD repeat domain 59 (*WDR59*) gene (**Table 1, Figure S5**). This variant was linked to type 1 diabetes, although did not reach genome-wide significance (*p*=8.7×10^-6^). The significant SNP reported in our previous GWAS of sepsis-associated ARDS (rs9508032, intronic to *FLT1),* (Guillen-Guio et al., 2020) did not show a significant association with sVEGFR1 levels (**Table S4**).

**Figure 2.**
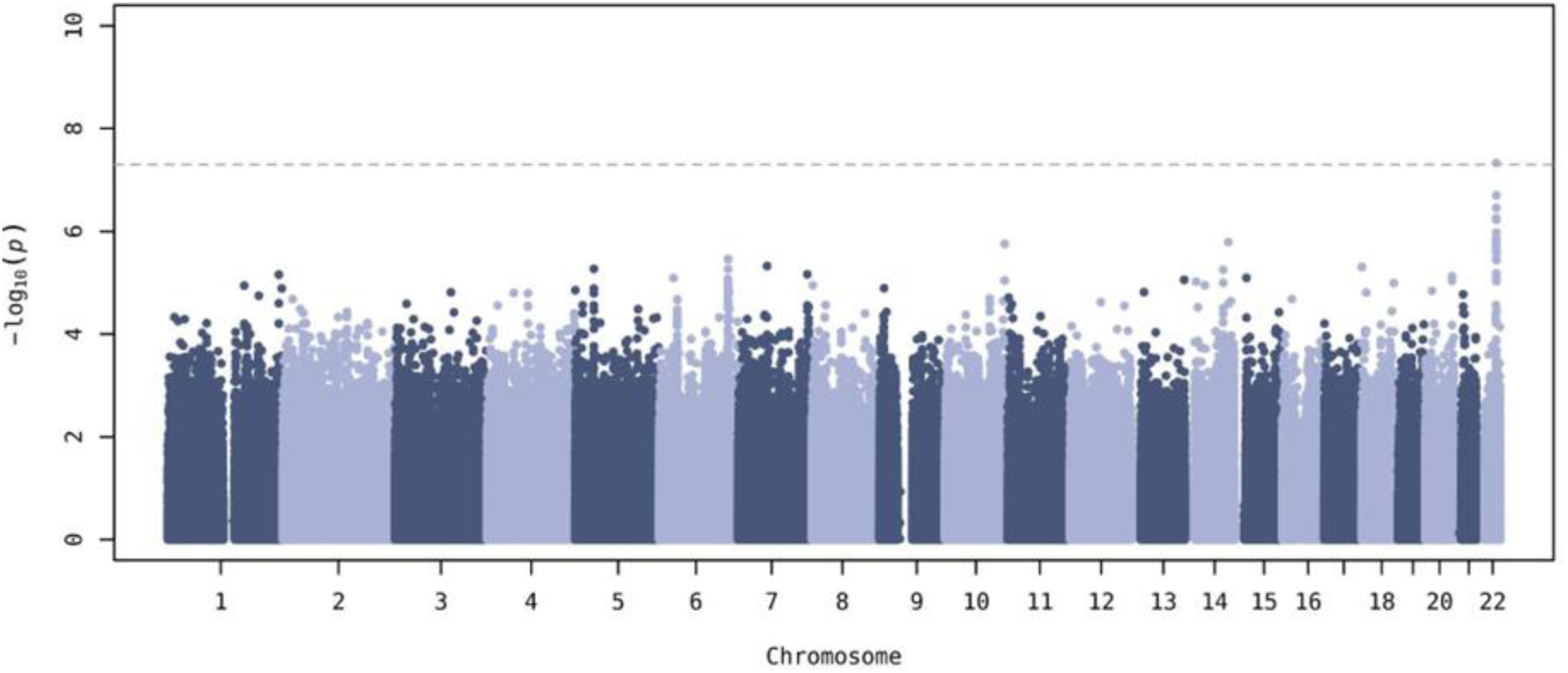
Manhattan plot of the genome-wide association study results for VEGFR1 levels at T2. The x-axis represents the chromosomal positions (GRCh37/hg19), and the y-axis shows the -log10(*p*-value). The horizontal dash line represents the genome-wide significance threshold (*p*-value=5.0×10^−8^).

**Figure 3.**
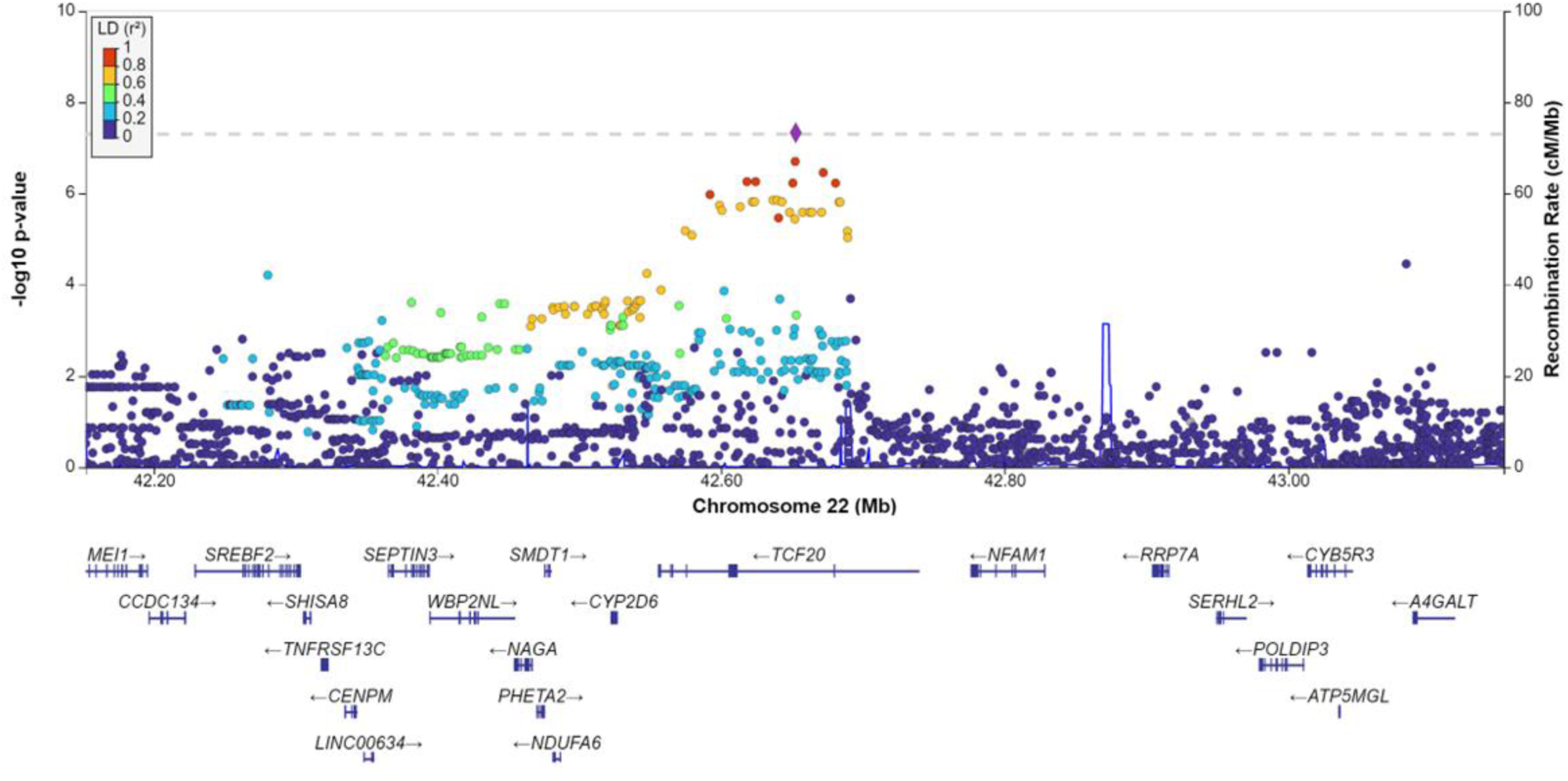
Regional plot of the association results for the genome-wide significant locus of VEGFR1 levels at T2. The x-axis represents the chromosomal positions (GRCh37/hg19), and the y-axis shows the -log10(*p*-value). The colour scheme of the top left legend represents the linkage disequilibrium (LD) values (r2) with the significant variant (rs134871) and is based on the European population data from The 1000 Genomes Project. The horizontal dash line indicates the genome-wide significance threshold (*p*-value=5.0×10^-8^). The plot was generated using LocusZoom (http://locuszoom.org/).

**Table 1.** Results of the top association signals with sVEGFR1 levels.

| GWAS | SNP | Position | NEA/EA | p-value | Beta | SE | EAF | Nearest gene |
| --- | --- | --- | --- | --- | --- | --- | --- | --- |
| T1 | rs59679804 | 3:183633094 | G/A | $1.78 \times 10^{-7}$ | -0.16 | 0.03 | 0.40 | *ABCC5/PARL* |
| T2 | rs134871 | 22:42652716 | T/C | $4.66 \times 10^{-8}$ | -0.23 | 0.04 | 0.46 | *TCF20* |
| T7 | rs72798674 | 16:74999522 | T/A | $1.52 \times 10^{-6}$ | -0.58 | 0.11 | 0.07 | *WDR59* |

EA: effect allele, NEA: non-effect allele, Position: chromosome and base pair based on GRCh37/hg19, SE: standard error, EAF: effect allele frequency.

The Bayesian fine mapping around the genome-wide significant pQTL identified 19 SNPs in the 95% credible set (**Figure S6, Table S5**). Out of those, we prioritised rs762995 intronic to *TCF20* based on the CADD score as a variant with a high biological impact in the credible set (*p*=3.49×10^-7^, beta= - 0.21, SE=0.04, CADD=22.1). The rs134871_C and rs762995_G alleles, linked to decreased sVEGFR1 levels, have previously been associated with an increase in PDW (*p*=2.9×10^-12^ and *p*=2.7×10^-8^, respectively), asthma risk (*p*=2.40×10^-3^ and *p*=2.00×10^-4^), and other hematopoietic traits (**Table S6**). Importantly, these two SNPs are functionally linked to the nearby Cytochrome P450 Family 2 Subfamily D Member 6 (*CYP2D6*) gene, widely involved in the metabolism of diverse drugs (Zanger & Schwab, 2013), according to the evidence integrated in the V2G score (**Table S5**).

Further *in silico* analyses highlighted additionally relevant functional consequences (**Table S7**) for the significant (rs134871) and prioritised (rs762995) pQTLs. These results support that they are located within regulatory elements (enhancer and promoter histone marks and DNase I hypersensitive sites), especially in primary T-cells, and lung and spleen tissues, among others. The MAGMA gene-set analysis of the 41 genes within the locus revealed that the strongest associations were for *TCF20* (*p*=5.18×10^-5^) and *CYP2D6* (*p*=1.17×10^-3^), both surpassing the Bonferroni significance (*p*=1.22×10^-3^) (**Table S8)**. Furthermore, the pQTL locus colocalised (PP(H4)>80%) with eQTLs of 11 of these genes in 25 tissues (**Table S9**), including *TCF20* (83.5%) in the suprapubic non-sun-exposed tissue and *CYP2D6* (96.1%) in the liver (**Table S9 and Figure S7**).

We then evaluated the association of low-frequency variants in the two prioritised genes, *TCF20* and *CYP2D6*, with sepsis-induced ARDS risk. SKAT-O tests revealed that low-frequency missense variants in *TCF20* were associated with ARDS (*p=*0.003) (**Table 2**). The association included 15 variants that met our criteria, and the functional annotation revealed a high biological impact for 10 of them based on their CADD score (CADD>15), most of them located in the first exon of the gene (**Table S10**). In addition, the different functional annotations revealed possible pathogenic variants. The small genetic damage index (GDI) calculated for *TCF20* (GDI: 7.94) suggested that the gene is less susceptible to carry deleterious variation compared to other genes, and it may be under stronger selection pressure. Therefore, the presence of these missense variants may have a greater impact on its function. Collectively, the result of the logistic regression model shows that *TCF20* missense rare variants were associated with reduced risk of ARDS among sepsis patients (OR [95% CI]=0.76 [0.63-0.92], *p*=0.005), and this result was robust to sex, age, and APACHE II adjustments (OR [95% CI]=0.76 [0.63-0.93], *p*=0.006).

**Table 2.** Association of low frequency variants in the prioritised genes with sepsis-associated ARDS.

| Gene | Location | Test | *p*-value |
| --- | --- | --- | --- |
| *CYP2D6* | chr22:42,522,501-42,526,908 | AF<0.01 | 0.664 |
|  |  | AF<0.01 & missense | 0.635 |
|  |  | AF<0.01 & CADD>15 | 0.672 |
| *TCF20* | chr22:42,556,019-42,739,622 | AF<0.01 | 0.519 |
|  |  | **AF<0.01 & missense** | **0.003** |
|  |  | AF<0.01 & CADD>15 | 0.07 |

Location: chromosome, start and end based on ensemble coordinates (GRCh37/hg19); AF: Allele frequency

The PGS models of the pQTLs of sVEGFR1 levels at each collection time were significantly associated with sepsis aggravation at T1 and T7. No models were significantly associated with sepsis aggravation at T2 (**Table S11**). At T1, the best PGS model resulted for 2,116 independent variants (threshold set at *p*=1.82×10^-3^) and was associated with ICU mortality (OR[95% CI]=0.65[0.54,0.80]; *p*=2.61×10^-5^). The best PGS at T7 integrated 3,094 variants (threshold set at *p*=2.84×10^-3^) and was associated with susceptibility to ARDS (OR[95% CI]=0.70[0.58,0.84]; *p*=1.04×10^-4^). In these two PGS models, patients with higher scores were associated with a better prognosis. That is, patients carrying variants associated with increased sVEGFR1 levels were associated with lower risk of sepsis-associated ARDS and lower ICU mortality (**Table S11**).

These two more significant PGS models included variants linked to a total of 2,666 genes in T1 and 3,438 genes in T7. To conduct a simplified gene set enrichment analysis on these, we opted to assess their intersection (1,350 shared genes). This revealed an enrichment of genes that serve as common targets for treatments that have shown efficacy in ARDS patients (**Table S12**), being trichostatin A the most significant one (*p*_-adjusted_=9.81×10^-58^). Significant treatment enrichments also included dexamethasone (*p*_-adjusted_=2.70×10^-3^) (Villar et al., 2020), rosiglitazone (*p*_-adjusted_=8.15×10^-4^) (He et al., 2019), and doxycycline (*p*_-adjusted_=0.03) (Sauer et al., 2021) (**Table S12**). Results also evidenced an enrichment of genes implicated in infectious-respiratory conditions, such as severe COVID-19 (*p*_-adjusted_=7.49 x10^-16^) and tuberculosis (*p*_-adjusted_=3.23×10^-5^), as well as lung function (FEV1/FVC) (*p*_-adjusted_=1.22×10^-4^) (**Table S13**). Finally, there was an enrichment in genes of signal transduction (*p*_-adjusted_=2.62×10^-6^), receptor tyrosine kinase signalling (*p*_-adjusted_=4.22×10^-4^), and cell adhesion pathways, including adherens junction interactions (*p*_-adjusted_=0.010) and cell-cell junctions (*p*_-adjusted_=0.040) (**Table S14**).

To understand the effect of VEGFR1 and VEGFA on the causality of sepsis-associated ARDS, we then conducted a two-sample bidirectional MR study. For instrumental variable selection, we performed an independent GWAS based on serum levels of VEGFR1 (n=46,836) and VEGFA (n=47,498), separately, in UKBB participants. Manhattan and QQ plots with the inflation factor of the VEGFR1 and VEGFA levels association are available in the **supplemental Figures S8 and S9**. The rs134871 association was not replicated in this population-based cohort (**Table S3**). For VEGFR1, we selected the 16 SNPs that met the criteria for strong IVs (independent variants with *p* <5×10^-8^ and F-statistic >10). The IVW-based results suggested that variants associated with VEGFR1 levels did not suggest a causal relationship with ARDS susceptibility (*p*=0.536; OR [95%CI]=0.33[0.01-10.72]. Sensitivity analyses with five other MR methods did not support causality (**Supplementary Table S15**). For VEGFA, we selected 36 strong IVs. However, neither the IVW analysis (*p*=0.992, OR [95%CI]=1.00[0.59-1.70]) or any of the other MR methods supported a causal effect of VEGFA on ARDS susceptibility (**Supplementary Table S15**). Neither the Q-statistic nor the Egger intercept evidenced the presence of pleiotropy, and no outliers were detected for VEGFR1 or VEGFA analyses.

Subsequently, we tested whether the variants that were associated with ARDS susceptibility have a causal effect on serum levels of VEGFR1 or VEGFA. We included 42 independent association signals with *p*<5×10^-6^ and F-statistic >10 as strong instruments for ARDS risk from the discovery stage of our previous GWAS of sepsis-associated ARDS (Guillen-Guio et al., 2020). The use of MR-RAPS did not improve the results in any of the cases (**Supplementary Table S16**). The Q-statistic suggested pleiotropy for VEGFA assessment, and one outlier was detected (**Supplementary Table S16**). These results suggest that no causality effect can be inferred between serum levels of VEGFA or VEGFR1 and ARDS, or vice versa.

## Discussion

Our previous GWAS implicated the *FLT1* gene variation in the sepsis-associated ARDS susceptibility. The study of biomarkers over time is key for understanding sepsis pathophysiology, inflammatory patterns, disease progression, and patient outcomes. Thus, here we screened the genetic variation associated with the serum sVEGFR1 levels in patients with sepsis at different time points during the acute systemic process. By integrating different genetic approaches, including common and low-frequency variant association analysis, to identify sVEGFR1 pQTLs with potential biological and clinical relevance, as well as a gene expression assessment, we uncovered *TCF20* as a novel risk gene for sepsis-associated ARDS.

We identified pQTLs associated with decreased sVEGFR1 levels located within the first intron of *TCF20*. *In silico* functional analysis and the colocalisation study suggest that these genetic variants have a potential regulatory role in many genes including *TCF20*. Moreover, low-frequency missense variants in *TCF20* were significantly associated with susceptibility to ARDS by sepsis and a functional annotation of these variants revealed potential pathogenic effects. *TCF20* is a transcription factor regulating the activity of different elements, many of them involved in immunity, and has been associated with developmental and cognitive disorders (Huang et al., 2023; Lyngsø et al., 2000; Rekdal et al., 2000; Vetrini et al., 2019). However, some patients with *TCF20* mutations show immunological abnormalities such as autoimmune hepatitis, hyper IgE syndrome, immune thrombocytopenic purpura, and wheezy bronchitis (Huang et al., 2023; Torti et al., 2019). Circular RNA of this gene has been shown to be downregulated in patients with sepsis-associated ARDS and in a mice model with sepsis-associated acute lung injury, suggesting its contribution to the pathophysiology of ARDS (Xu et al., 2024). Despite the lack of evidence linking TCF20 to VEGFR1 regulation, VEGFA has been shown to down-regulate TCF20 (Li et al., 2021). In addition, a Tcf20 deficient mice model developed liver fibrosis, mitochondrial metabolism abnormalities, extracellular matrix turnover, and increased levels of succinate (Córdoba-Jover et al., 2023). These findings are present also in patients carrying loss-of-function mutations in the gene (Córdoba-Jover et al., 2023). Succinate is involved in inflammatory processes, angiogenesis, and hypoxia (Huang et al., 2024). Increased succinate levels have been observed in patients with post-injury ARDS that may be involved in the pulmonary sequestration of neutrophils (Nunns et al., 2022). Moreover, succinate could be a biomarker of progression in idiopathic pulmonary fibrosis and may promote fibroblast activation (He et al., 2024). Taken together, this evidence supports that TCF20 may play a key role in the immune response and the accompanying fibrotic processes, during early stages of the exudative phase or in the fibroproliferative phase of ARDS (Blondonnet et al., 2016; Spadaro et al., 2019).

The significant and prioritised pQTLs have been previously associated with PDW. This platelet measure has been proposed as a clinical predictor of severity in various infectious-respiratory diseases, ICU mortality, or the need for mechanical ventilation (Białas et al., 2017; Dubey et al., 2021; Ligi et al., 2024). Studies in mice showed that the maturation of megakaryocytes, the precursor cells of platelets, is enhanced by VEGFA administration. This effect is mediated by VEGFR1, which contributes to the increase in platelet levels. Inhibition of VEGFR1 reversed the effect of VEGFA without affecting basal platelet levels, suggesting a role related to inflammatory rather than homeostatic processes (Pitchford et al., 2012). Although we did not detect association of these variants with ARDS susceptibility, platelets play a critical role in hemostasia, coagulation, thrombosis, as well as in vascular integrity, and oedema formation. Their contribution in sepsis and ARDS has been described (Middleton et al., 2016). Therefore, the study of platelets as immune effector cells is crucial for the comprehension of acute inflammatory responses, and the variants associated with platelet levels and their characteristics may be an important point in understanding their activity and contribution to disease progression.

We detected a significant increase in the sVEGFR1 levels over time after sepsis diagnosis. The soluble fraction of VEGFR1 has an anti-inflammatory role in the host response since this nonmembrane form can competitively bind to VEGFA inhibiting its activity and reducing the inflammatory response and angiogenesis. Increased sVEGFR1 levels have been associated with a worse prognosis and mortality (Shapiro et al., 2010; Yang et al., 2011). While its regulation and inflammatory role are not fully understood, it has been suggested that these observations may be due to the need for an intense anti-inflammatory response to combat sepsis (Greco et al., 2018). In addition, VEGFA has been shown to induce the expression of sVEGFR1 in vascular endothelial cells, which may act as negative feedback regulator (Saito et al., 2013). PGS estimates of the increased sVEGFR1 levels were associated with lower risk of ARDS by sepsis and with lower ICU mortality. Interestingly, genes contributing to the PGS models were enriched in targets for treatments of key interest for ARDS including trichostatin A, rosiglitazone, and dexamethasone. Bidirectional MR analyses did not provide evidence of causality of VEGFR1 levels on ARDS risk or vice versa, nor for analyses based on its main ligand VEGFA. Sepsis-specific eQTLs have been identified, which are undetectable in population controls (Burnham et al., 2024). Given the potential impact of acute systemic processes on gene regulation variation, further studies are needed to determine whether causality may be mediated by pQTLs specific to critically ill patients.

We found that the polygenic component of the sVEGFR1 levels was also associated with susceptibility to ARDS and with ICU mortality. Patients with higher PGS scores, i.e., carrying alleles associated with increased sVEGFR1 levels, were protected from ARDS and death in the ICU. This might be a consequence of the anti-inflammatory activity of sVEGFR1 described above. Most importantly, PGS models can be used to explore the genetic overlap between traits (Guillen-Guio et al., 2024; van Reij et al., 2020) and to reveal potential drug targets to enable drug repurposing (Wang et al., 2024). In this respect, our analyses on the gene lists linked to the PGS models of the sVEGFR1 levels identified genes associated with lung function, severe infectious-respiratory diseases, and of key treatments for ARDS, some of which are currently in use in the clinical practice. Trichostatin A was the most significant drug identified by this enrichment. Trichostatin A is known to inhibit class I and II histone deacetylases and VEGF expression under hypoxic conditions (Kang et al., 2012), and has been shown to reduce pulmonary fibrosis and ventilation-induced lung injury in mice models (Li et al., 2017). Despite the available evidence, there is lack of clinical trials testing the efficacy of the drug in ARDS patients.

To our knowledge, this is the first study that integrates different approaches to identify the regulatory mechanisms of the sVEGFR1 protein in sepsis. However, we recognise some limitations of our study. First, the sample size used in the study is limited, which reduces statistical power. In addition, a replication cohort is needed to validate our findings, and future studies should include participants of non-European ancestry to improve the transferability. Furthermore, although we analysed the effects of rare variants in the targeted genes of interest, genome-wide studies focused predominantly on common variants, overlooking the importance of rare variants. Finally, even though MR analyses suggested the absence of causality between serum levels of VEGFA or VEGFR1 and ARDS, and vice versa, instrumental variable selection in these studies did not involve critically ill patients, which may have important consequences for the regulatory mechanisms of VEGFR1. Additionally, it must be noted that the VEGFR1 measurements in UKBB result from an aggregate of both the soluble and membrane-bound fractions. Thus, possible functional differences between these forms should be carefully investigated, especially in the context of acute processes. Therefore, caution must be taken when interpreting the results, as this may have critically contributed to the lack of replication of the findings in the GEN-SEP study.

### Conclusion

We identified *TCF20* as a novel gene for ARDS. Although its role in sVEGFR1 regulation is unclear, we provide evidence to support its association with susceptibility to ARDS by sepsis. Subsequent analyses also pinpoint *CYP2D6*. Further proteogenomic studies in ever growing protein sets will help to improve our understanding of ARDS susceptibility.

## Data Availability

All data produced in the present study are available upon reasonable request to the authors

