## Supplementary material for "Genetic regulation of the vascular endothelial growth factor receptor 1 during sepsis and association with ARDS susceptibility"

GEN-SEP study

The GEN-SEP study is a Spanish multicentre network of post-surgical units and intensive care units (ICUs) enrolling unrelated adult patients (>18 years) with sepsis of European ancestry. The diagnosis of sepsis was clinically defined according to the Third International Consensus Definitions for Sepsis (Singer et al., 2016). For this study, 292 patients with sepsis were recruited between 2002 and 2019 (Hernandez-Beeftink et al., 2022) from the following hospitals: Hospital Universitario de Canarias, Tenerife; Hospital Universitario Nuestra Señora de Candelaria, Tenerife; Hospital Universitario Río Hortega, Valladolid; Hospital Universitario Dr. Negrin, Gran Canaria; Hospital General de Ciudad Real, Ciudad Real; Complejo Hospitalario Universitario de León, León; Hospital Virgen de la Luz, Cuenca; Complejo Hospitalario Universitario de Santiago de Compostela, Santiago de Compostela; Fundació Althaia-Manresa, Barcelona; Hospital Clinic, Barcelona; Hospital Clínico de Valladolid; Hospital La Paz, Madrid; Hospital Fundación Jiménez Díaz, Madrid; Hospital del Bierzo, Ponferrada; Hospital General Río Carrión, Palencia, and Hospital Virgen de la Concha, Zamora.

Four ml of peripheral blood were withdrawn at three times: T1 (time of inclusion into the study, within 24 hours after diagnosis), T2 (48-72 hours after diagnosis), and T7 (7 days after diagnosis). DNA extraction was performed on samples collected at T1 using Illustra™ blood genomicPrep Mini Spin Kit (GE Healthcare). The DNA concentration was measured on the Qubit 3.0 fluorometer with the dsDNA HS Assay kit (Thermo Fisher Scientific).

Genotyping, quality controls and variant imputation

Genotyping of GEN-SEP patients was carried out on samples collected at T1, for the detection of single nucleotide polymorphisms (SNPs) using the Axiom Genome-Wide CEU1 array (Affymetrix) in the National Genotyping Centre (CeGen), Universidad de Santiago de Compostela Node, Spain. Intensity data were processed for variant calling using AffyPipe v2.10.0 software following the manufacturer's recommendations. The quality controls were performed using R (v3.6.0) (R Core Team, 2022) and PLINK v1.9 (Chang et al., 2015). Variants were filtered to exclude if they had a genotyping rate < 95%, a minor allele frequency (MAF) <1%, or deviations from Hardy Weinberg equilibrium (HWE, $p<1.0 \times 10^{-6}$). Individuals with a high degree of relatedness (PIHAT> 0.2), deviations in heterozygosity or incomplete clinical data were also excluded from the study. The principal component analysis (PCA) was performed to assess genetic heterogeneity among patients and to exclude outliers (exceeding four standard deviations). Variant imputation was performed on the Michigan Imputation Server, using "Haplotype Reference Consortium" (HRC r1.1 2016 - GRCh37/hg19) as the reference panel.

Whole-exome sequencing and variant annotation

Whole-exome sequencing (WES) was obtained from 822 patients of the GEN-SEP cohort 272 with sepsis-associated ARDS and 550 at-risk controls with sepsis from the GEN-SEP study. Sequencing libraries were prepared using various NGS library preparation protocols, including DNA Prep with Enrichment kit (Illumina Inc.) and SureSelect XT HS2 DNA (Agilent Technologies Inc.), according to the procedures described elsewhere (Díaz-de-Usera et al., 2020). Library sizes and quantification were obtained with a TapeStation 4200 (Agilent Technologies Inc.) and the Qubit dsDNA HS Assay (Thermo Fisher Scientific). Sequencing was carried out at the Instituto Tecnológico y de Energías Renovables (ITER, Santa Cruz de Tenerife, Spain) on various Illumina platforms including Illumina NextSeq550, HiSeq4000, and NovaSeq6000, with paired end reads of 75 or 100 base in length, as determined by the sequencing instrument, with an average depth of 100X, and included a 1% PhiX V3 control (Illumina Inc.). Pre-processing of sequencing reads was conducted using bcl2fastq v2.18 for demultiplexing and BWA-MEM v0.7.15 (https://github.com/lh3/bwa) to align reads to the GRCh37/hg19 reference. SAMtools v1.3 (http://www.htslib.org) and Picard v2.10.10 (https://broadinstitute.github.io/picard) were used for BAM quality controls. Small germline variant calling (SNPs and indels up to 50 bp) was conducted with GATK HaplotypeCaller v3.8 (https://gatk.broadinstitute.org/hc/en-us/articles/360037225632-HaplotypeCaller), using a 100 bp padding around capture targets and following the GATK Best Practices workflow. Resulting variants were filtered with BCFtools v1.16 based on the "PASS" filter, missingness (FMISS) ≤0.05, genotype quality (GQ) ≥20, and depth of coverage (DP) ≥10.

Functional variant annotation was performed for the genes reported in this study, to include functional information for population allele frequency in 1000 Genomes Phase 3 Europeans population, and gnomAD v.2.1 non-Finnish Europeans populations, variant type, protein function, pathogenic potential, and the genetic damage index (GDI) using Ensembl VEP v105 and ANNOVAR v07.06.20 (https://annovar.openbioinformatics.org/en/latest/) based on different databases. All analyses were performed with the support of Teide-HPC Supercomputing facility (http://teidehpc.iter.es/en) at ITER.

Gene-based low frequency variant analysis

To assess the effect of exonic rare variation for the genes of interest detected in this study, we carried out a gene-based rare variation analysis. We performed a gene-based association using the Sequence Kernel Association Test - Optimal (SKAT-O) by Regenie v.3.2.7 (Mbatchou et al., 2021). The association was performed on 260 cases (sepsis-induced ARDS) and 537 controls with sepsis. Following the Regenie workflow, we performed a first step fitting a whole-genome model using common variation from the SNP array data. In the second step, input variants were filtered to be using different masks: (i) AF01: variants with allele frequency (AF) < 0.01 in 1000 Genomes Phase 3 in Europeans population, or gnomAD 2.1 Non-Finnish Europeans populations, or with missing data in both; (ii) AF01 and missense: variants matching as AF01 filtered and VEP v.105 consequence field matches “missense”; (iii) AF01 CADD: variants matching AF01 filter and with CADD v.1.6 phred score > 15.00.

Polygenic scores

We derived polygenic score (PGS) models to evaluate the association between pQTLs of sVEGFR1 and ARDS susceptibility, as well as ICU mortality. As base dataset, we used the results obtained for each one of the GWAS of soluble VEGFR1 (sVEGFR1) levels. We applied different *p*-value thresholds to screen multiple PGS models. These scores were evaluated in a target dataset, consisting of an independent cohort of individuals from the GEN-SEP study, to calculate individual PGS and test their association with the two phenotypes. The PGS were calculated using PRSice 2.3.5 (R 4.2.2) (Choi & O'Reilly, 2019), according to the following formula:

$$PGS_j = \sum_{i}^{n} \beta_i G_{ij}$$

where β is the logarithm of odd ratio and G is the genotype of variant i in individual j, and n is the number of SNPs includes in the score. We generated different PGS models, setting different *p*-value thresholds on the GWASs of sVEGFR1 levels at T1, T2, and T7, selecting independent SNPs (r2 = 0.1 in 250 kb windows). We excluded palindromic variants (A/T, C/G) of the analyses. The individual PGS were normalised by:

$$PGS_j = \frac{\sum_i(\beta ixGij) - Mean(PGS)}{SD(PGS)}$$

Subsequently, we tested whether the variants associated with sVEGFR1 levels could predict ARDS susceptibility or ICU mortality in patients with sepsis. To avoid bias, an independent cohort of individuals from the GEN-SEP study who were not included in the sVEGFR1 GWASs was used as a target to test PGS models. Susceptibility to ARDS was tested in 556 patients, 338 controls with sepsis and 218 cases with ARDS by sepsis. The diagnosis for ARDS patients was established according to the Berlin definition (Ferguson et al., 2012). For mortality, we included 545 patients, 369 surviving and 176 non-surviving patients who died while admitted to the ICU. The normalised PGSs were assessed using logistic regressions, adjusting for sex, age and APACHE II. The best models (i.e., the top-most significant) were selected for each collection time.

Mendelian Randomisation

We performed a GWAS of VEGFA and VEGFR1 serum levels separately in individuals from UKBB to identify protein quantitative trait loci (pQTLs). We included individuals in UK Biobank with both imputed genotype and Olink® (Thermo Fisher Scientific) protein data, restricted to samples from Europeans as previously defined (Shrine et al., 2023). Association was performed using Regenie v3.2.6. Age, sex, array version, first ten PC, and protein batch were included as covariates. The GWAS of

VEGFA levels comprised a total of 47,498 individuals, while the GWAS of VEGFR1 included 46,836 individuals. Only variants with MAF>1% and an imputation quality score >0.5 were considered in following Mendelian Randomisation (MR) analyses.

We performed bidirectional two-sample MR analyses to determine the causal effect of VEGFA and VEGFR1, separately, on sepsis-induced ARDS and vice versa. The instrumental variables (IVs) were obtained from summary data from the: i) GWAS based on VEGFA levels in 47,498 individuals from UKBB; ii) GWAS based on VEGFR1 levels in 46,836 individuals from UKBB; iii) Discovery stage of our prior GWAS of sepsis-associated ARDS (Guillen-Guio et al., 2020), which included 316 patients with sepsis and 274 patients with sepsis-associated ARDS from the GEN- SEP cohort.

We selected "strong" instruments with F-statistic>10 and significantly associated ($p<5x10^{-8}$) with VEGFA and VEGFR1 levels. Due to the absence of significant signals at genome-wide level on the discovery stage of the ARDS GWAS, the suggestive significance threshold at $p=5x10^{-6}$ defined in our previous work was used. To determine the causal effect, we used the inverse variance weight (IVW) model. Five other complementary approaches were tested as sensitivity analyses: MR Egger, MR-PRESSO (MR Pleiotropy RESidual Sum and Outlier), simple mode, weighted median, and weighted mode. For IVW and MR-Egger, we calculated the Q-statistic to assess the heterogeneity of the selected instruments. The effect of horizontal pleiotropy was also assessed using the Egger intercept. The effect of outliers was controlled through MR-PRESSO. We used MR-RAPS to evaluate causality using "weaker" instruments, selecting as IVs all variants with $p<0.05$.

For MR analysis, independent variants ($r^2> 0.001$) were extracted using PLINK 1.9 based on the European 1000 Genomes population and their effects were harmonised using the *TwoSampleMR* R package. We excluded palindromic variants (A/T, C/G) for which positive strand allele frequencies could not be inferred and lacked suitable proxies ($r^2>0.8$, based on LDlink (Machiela & Chanock, 2015) from the analyses. MR analyses were performed under R 4.0.0 version using *TwoSampleMR* for IVW, MR Egger, simple mode, weighted median, and weighted mode models (Hemani et al., 2018). The *MRPRESSO* package was used for MR-PRESSO method and *mr.raps* was used for MR-RAPS (Verbanck et al., 2018; Zhao et al., 2020).

### SUPPLEMENTARY RESULTS

**Table S1. Demographic and clinical features of GEN-SEP patients after quality controls**.

| | **Patients (n=241)** |
|---|---|
| **Gender** | |
| Female | 50.62 % |
| Male | 49.38 % |
| **Age, mean years ± SD** | 64.07 ± 16.05 |
| **BMI, mean kg/m2 ± SD (N=208)** | 27.38 ± 10.73 |
| **sVEGFR1 normal levels (pg/ml):** | |
| T1 (n=225) | 3.68 ± 0.32 |
| T2 (n=169) | 3.86 ± 0.41 |
| T7 (n=108) | 3.94 ± 0.46 |
| **APACHE II (8h), mean ± SD** | 18.80 ± 7.38 |
| **Comorbidities$ (N=240)** | 65.41% |
| **ARDS** | 34.85 % |
| **ICU mortality** | 18.67 % |
| **Days in hospital, mean ± SD (N=240)** | 33.12 ± 40.09 |
| **Sepsis of pulmonary origin, (N=236)** | 70.76 % |
| **Pathogen, (N=210)** | |
| Gram-positive | 20.00 % |
| Gram-negative | 35.71 % |
| Gram-positive and Gram-negative | 15.24 % |
| Others+ | 11.43 % |

$Comorbidities include cancer, age >80 years, hepatopathy, valvular disease, immunodeficiency, morbid obesity, chronic disease, autoimmune disease, ischemic cardiopathy, pneumonia, and serious recurrent infections. +Others include both fungi, virus and polymicrobial. APACHE II, Acute Physiology and Chronic Health Evaluation II; ARDS, Acute Respiratory Distress Syndrome; BMI, Body Mass Index; ICU, Intensive Care Unit.

**Table S2. Sensitivity analysis of the rs134871 association with serum sVEGFR1 levels obtained within 48-72 hours after sepsis diagnosis (T2).**

| Model | *p*-value | Beta(SE) | N |
|---|---|---|---|
| **Unadjusted*** | $2.58x10^{-7}$ | -0.22(0.04) | 169 |
| **Sex, age, APACHE II** | $4.66x10^{-8}$ | -0.23(0.04) | 169 |
| **First 5 PC** | $4.50x10^{-8}$ | -0.23(0.04) | 169 |
| **BMI** | $1.10x10^{-6}$ | -0.21(0.04) | 150 |
| **Sepsis of pulmonary origin** | $8.08x10^{-8}$ | -0.23(0.04) | 165 |
| **ARDS** | $6.43x10^{-8}$ | -0.23(0.04) | 169 |
| **ICU mortality** | $3.88x10^{-8}$ | -0.23(0.04) | 169 |
| **GRAM +** | $2.65x10^{-8}$ | -0.25(0.04) | 149 |
| **GRAM -** | $4.98x10^{-8}$ | -0.24(0.04) | 149 |
| **GRAM + & GRAM -** | $7.47x10^{-8}$ | -0.24(0.04) | 149 |
| **Other microorganisms** | $6.11x10^{-8}$ | -0.24(0.04) | 149 |

ARDS: acute respiratory distress syndrome; BMI: body mass index; ICU: intensive care unit, PC: principal components; SE: standard error. Other microorganisms include both fungi, virus and polymicrobial infections. *Model without adjusting for any covariates, the remaining models include sex, age and APACHE score as covariates.

**Table S3. Association results of rs134871 in GWAS of sVEGFR1 levels, VEGFA levels, and sepsis-induced ARDS susceptibility.**

| GWAS | EAF | *p*-value | Beta | SE | N | NEA/EA |
|---|---|---|---|---|---|---|
| **GWAS sVEGFR1 T1** (GEN-SEP) | 0.47 | $4.43x10^{-3}$ | -0.08 | 0.03 | 225 | T/C |
| **GWAS sVEGFR1 T2 (GEN-SEP)** | 0.46 | $4.66x10^{-8}$ | -0.23 | 0.04 | 169 | T/C |
| **GWAS sVEGFR1 T7** (GEN-SEP) | 0.48 | $9.20x10^{-3}$ | -0.16 | 0.06 | 108 | T/C |
| **GWAS ARDS susceptibility*** | 0.49 | 0.722 | -0.04 | 0.12 | 590 | T/C |
| **GWAS VEGFR1** (UKBB) | 0.52 | 0.101 | 0.003 | 0.002 | 46,836 | T/C |
| **GWAS VEGFA** (UKBB) | 0.52 | 0.856 | -0.001 | 0.004 | 47,498 | T/C |

EA: effect allele; EAF: effect allele frequency; NEA: non-effect allele; SE: standard error. T1: sVEGFR1 levels obtained within 24 hours after sepsis diagnosis; T2: sVEGFR1 levels obtained within 48-72 hours after sepsis diagnosis; T7: sVEGFR1 levels obtained 7 days after sepsis diagnosis. *Results for the discovery stage of Guillen-Guio et al. (Guillen-Guio et al., 2020) that includes 274 cases with ARDS and 316 controls with sepsis from GEN-SEP cohort.

**Table S4. Association results of the previously described significant sepsis-induced ARDS susceptibility variant (rs9508032) in the GWAS of sVEGFR1 serum levels.**

| GWAS | EAF | *p*-value | Beta | SE | N | NEA/EA |
|---|---|---|---|---|---|---|
| **GWAS sVEGFR1 T1** (GEN-SEP) | 0.28 | 0.76 | -0.01 | 0.03 | 225 | C/T |
| **GWAS sVEGFR1 T2** (GEN-SEP) | 0.28 | 0.78 | 0.01 | 0.05 | 168 | C/T |
| **GWAS sVEGFR1 T7** (GEN-SEP) | 0.29 | 0.92 | 0.01 | 0.07 | 108 | C/T |

EA: effect allele; EAF: effect allele frequency; NEA: non-effect allele; SE: standard error. T1: sVEGFR1 levels obtained within 24 hours after sepsis diagnosis; T2: sVEGFR1 levels obtained within 48-72 hours after sepsis diagnosis; T7: sVEGFR1 levels obtained 7 days after sepsis diagnosis. *Results for the discovery stage of Guillen-Guio et al. (Guillen-Guio et al., 2020) that includes 274 cases with ARDS and 316 controls with sepsis from GEN-SEP cohort.

**Table S5. Bayesian fine mapping results of the significant locus at chromosome 22 around rs134871.**

| rsID | NEA/EA | EAF | *p*-value | Beta | SE | Gene(s) | $R^2$ | CADD | Best score [V2G]* |
|---|---|---|---|---|---|---|---|---|---|
| rs134871 | T/C | 0.46 | $4.66x10^{-8}$ | -0.23 | 0.04 | *TCF20* | - | 3.37 | 0.36 |
| rs134870 | T/C | 0.46 | $2.00x10^{-7}$ | -0.22 | 0.04 | *TCF20* | 0.92 | 5.03 | 0.36 |
| rs762995 | A/G | 0.46 | $3.49x10^{-7}$ | -0.21 | 0.04 | *TCF20* | 0.91 | 22.10 | 0.36 |
| rs2143138 | T/C | 0.46 | $5.51x10^{-7}$ | -0.21 | 0.04 | *TCF20* | 0.87 | 0.38 | 0.37 |
| rs5758661 | A/C | 0.46 | $5.51x10^{-7}$ | -0.21 | 0.04 | *TCF20* | 0.88 | 3.59 | 0.37 |
| rs134866 | C/T | 0.45 | $5.93x10^{-7}$ | -0.21 | 0.04 | *TCF20* | 0.91 | 4.14 | 0.37 |
| rs86669 | C/T | 0.45 | $5.93x10^{-7}$ | -0.21 | 0.04 | *TCF20* | 0.91 | 0.55 | 0.36 |
| rs5751239 | C/T | 0.45 | $1.06x10^{-6}$ | -0.20 | 0.04 | *TCF20* | 0.76 | 2.85 | 0.37 |
| rs5751250 | T/G | 0.45 | $1.40x10^{-6}$ | -0.20 | 0.04 | *TCF20* | 0.72 | 2.75 | 0.39 |
| rs5758670 | T/C | 0.45 | $1.40x10^{-6}$ | -0.20 | 0.04 | *TCF20* | 0.72 | 0.11 | 0.39 |
| rs5758659 | C/T | 0.45 | $1.52x10^{-6}$ | -0.21 | 0.04 | *TCF20* | 0.73 | 0.46 | 0.39 |
| rs5758660 | C/A | 0.45 | $1.52x10^{-6}$ | -0.21 | 0.04 | *TCF20* | 0.73 | 8.12 | 0.39 |
| rs5758677 | A/C | 0.45 | $1.52x10^{-6}$ | -0.21 | 0.04 | *TCF20* | 0.74 | 4.64 | 0.19 |
| rs134900 | G/C | 0.45 | $1.55x10^{-6}$ | -0.21 | 0.04 | *TCF20\LINC01315* | 0.74 | 0.68 | 0.38 |
| rs134902 | G/A | 0.45 | $1.55x10^{-6}$ | -0.21 | 0.04 | *TCF20\LINC01315* | 0.74 | 2.21 | 0.38 |
| rs5758645 | T/G | 0.44 | $1.81x10^{-6}$ | -0.21 | 0.04 | *TCF20* | 0.68 | 4.95 | 0.39 |
| rs5758653 | G/T | 0.44 | $1.96x10^{-6}$ | -0.20 | 0.04 | *TCF20* | 0.70 | 7.48 | 0.39 |
| rs5751241 | G/A | 0.44 | $2.35x10^{-6}$ | -0.20 | 0.04 | *TCF20* | 0.70 | 0.15 | 0.39 |
| rs5751255 | C/T | 0.44 | $2.58x10^{-6}$ | -0.20 | 0.04 | *TCF20* | 0.72 | 0.34 | 0.39 |

*V2G prioritise *CYP2D6* gene as the gene with the highest scores in all cases. CADD: combined annotation dependent depletion score (scaled) v.1.6; EA: effect allele; EAF: effect allele frequency; NEA: non effect allele; Pos: chromosome position and base pair based on GRCh37/hg19; $R^2$: linkage disequilibrium between variant with genome wide significant pQTL (rs134871); SE: standard error; V2G: Variant-to-Gene.

**Table S6. PheWAS results of significant and prioritised pQTLs**.

| SNP | EA | PheWAS |
|---|---|---|
| rs134871 | C | Platelet distribution width (*p*=2.90x$10^{-12}$, +); Mean spheric corpuscular volume (*p*=4.80x$10^{-4}$, -); White blood cell count (*p*=5.76x$10^{-4}$, +); Lung function (FEV1/FVC) (*p*=1.04x$10^{-3}$, +); Other local infections of skin and subcutaneous tissue (*p*=1.18x$10^{-3}$, +); Mean platelet (thrombocyte) volume (*p*=1.27x$10^{-3}$, +); Mean reticulocyte volume (*p*=1.50x$10^{-3}$, -); Mean sphered cell volume (*p*=1.82x$10^{-3}$, -); Asthma (*p*=2.40x$10^{-3}$, +); Eosinophil count (*p*=2.48x$10^{-3}$, +); Infection of intervertebral disc (pyogenic) (p=3.02x$10^{-3}$, +); SIR2-like protein 2 levels (*p*=3.04x$10^{-3}$, -); Herpesviral keratitis and keratoconjunctivitis (*p*=3.21x$10^{-3}$, -); Other erythematous conditions (*p*=3.40x$10^{-3}$, -); Flu -influenza vaccine (*p*=3.53x$10^{-3}$, +); Osteomyelitis (*p*=3.69x$10^{-3}$, +); NF-kappa-B essential modulator levels (*p*=4.28x$10^{-3}$, -); Neutrophil count (*p*=4.79x$10^{-3}$, +) |
| rs762995 | G | Platelet distribution width (*p*=2.50x$10^{-8}$, +); Lung function (FVC) (*p*=2.30x$10^{-5}$, -); Eosinophil counts (*p*=9.55x$10^{-5}$, +); Asthma (*p*=2.00x$10^{-4}$, +); Lung function (FEV1/FVC) (*p*=5.50x$10^{-4}$, +); Respiratory or ear-nose-throat disease (*p*=8.70x$10^{-4}$, +); Unspecified osteomyelitis (*p*=9.81x$10^{-4}$, +); White blood cell count (*p*=1.15x$10^{-3}$, +); Infection of intervertebral disc (p=pyogenic) (*p*=1.67x$10^{-3}$, +); Mean platelet volume (*p*=1.70x$10^{-3}$, +); Type 1 diabetes (*p*=1.86x$10^{-3}$, -); Eosinophil percentage (*p*=2.03x$10^{-3}$, +); Lymphocyte counts (*p*=2.07x$10^{-3}$, +); Mean sphered cell volume (*p*=2.79x$10^{-3}$, -); Whooping cough (*p*=3.54x$10^{-3}$, +); NF-kappa-B essential modulator levels (*p*=4.13x$10^{-3}$, -); Anti-herpes simplex virus 2 IgG seropositivity (*p*=4.19x$10^{-3}$, +); Other erythematous conditions (*p*=4.41x$10^{-3}$, -); Other/unspecified cytomegaloviral diseases (*p*=4.61x$10^{-3,}$-); White blood cell count (*p*=4.90x$10^{-3}$,+) |

EA: Effect allele; PheWAS: phenome-wide association study. Effect alleles align with the reported in the sVEGRF1 association, and the effect size is represented with + (beta > 0) and - (beta< 0). Results from OpenTarget genetic results (*p*<0.005) for traits related to infections, respiratory system, immunity or blood cell characteristics. For duplicate traits we indicated the most significant result. More information is available on https://genetics.opentargets.org/.

**Table S7. Functional analysis of significant and prioritised pQTLs.**

| | rs134871 | rs762995 |
|---|---|---|
| **Chromosome location** | 22:42652716 | 22:42672124 |
| **$R^2$** | - | 0.91 |
| **Genomic context** | Intronic (*TFC20*) | Intronic (*TFC20*) |
| **Freq (EUR non-Finnsh)** | 0.5035 | 0.5185 |
| **Rank [regulomeDB]** | 3a | 4 |
| **Score [regulomeDB]** | 0.874 | 0.609 |
| **Enhancer histone marks [HaploReg]** | | |
| H3K4me1 | Fetal lung fibroblasts cells (line: IMR90), Embryonic stem cells (lines: ES-I3, HUES6, HUES48, HUES64, ES-UCSF4), Primary mononuclear cells from peripheral blood, Primary T cells from peripheral blood, Primary T cells effector/memory enriched from peripheral blood, Primary T cells from cord blood, Primary T regulatory cells from peripheral blood, Primary T helper cells from peripheral blood, Primary T helper naive cells from peripheral blood, Primary T helper cells PMA-I stimulated, Primary T helper 17 cells PMA-I stimulated, Primary T helper memory cells from peripheral blood 1, Primary T helper memory cells from peripheral blood 2, Primary T CD8+ memory cells from peripheral blood, Primary T CD8+ naive cells from peripheral blood, Fetal Lung, Ovary, Fetal Adrenal Gland, Pancreas, Cervical Carcinoma Cell Line, Leukemia Cells (line: K562), Epidermal Keratinocyte Primary Cells, Lung Fibroblast Primary Cells, Osteoblast Primary Cells | Embryonic stem cells (lines: ES-WA7, H9, ES-I3, HUES6, HUES48, HUES64, H1, ES-UCSF4), Primary T cells from cord blood, Primary T helper cells from peripheral blood, Primary T helper naive cells from peripheral blood, Primary T helper cells PMA-I stimulated, NH-A Astrocytes Primary Cells, Epidermal Keratinocyte Primary Cells |
| H3K27ac | Primary T regulatory cells from peripheral blood, Primary T helper naive cells from peripheral blood, Primary T helper cells PMA-I stimulated, Primary T helper 17 cells PMA-I stimulated, Fetal Adrenal Gland, Spleen, T Cell Leukemia (line: Dnd41) | Embryonic stem cells (lines: H1, H9, HUES48 Cells, HUES64 Cells) |
| **Promoter histone marks [HaploReg]** | | |
| H3K4me3 | ES-UCSF4 Cells, Primary T cells effector/memory enriched from peripheral blood, Fetal Lung | Spleen |
| H3K9ac | Primary mononuclear cells from peripheral blood | Embryonic stem cells (lines: H9, HUES6, H1) |
| **DNAse [HaploReg]** | Epidermal Keratinocyte Primary Cells | Embryonic stem cells (lines: H9, H1), Placenta, Epidermal Keratinocyte Primary Cells |
| **Altered regulatory motifs [HaploReg]** | Pou2f2_known2, Rhox11, TAL1_known2 | CTCF_disc5, CTCF_disc8, Irf_disc4, Zbtb3 |
| **Proteins bound [HaploReg]** | None | MAX, NANOG, P300, POU5F1, YY1 |

$R^2$: linkage disequilibrium between variant with genome wide significant SNP (rs134871). 3a rank: transcription factor binding, any motif and chromatin accessibility peak; 4 rank: transcription binding and chromatin accessibility peak. Score from RegulomeDB: when closer to 1, it indicates more probability to be a regulatory variant. More information available on RegulomeDB (https://regulomedb.org/ ) and HaploReg (https://pubs.broadinstitute.org/) websites.

**Table S8. Gene associations resulting from the gene-set analysis by MAGMA within 1 Mb around the significant variant.**

| Gene | Location | SNPs | Zstats | *p*-value |
|---|---|---|---|---|
| *TCF20* | 22:42531019-42764622 | 502 | 3.88 | $5.2x10^{-5}$ |
| *CYP2D6* | 22:42497501-42551908 | 146 | 3.04 | $1.2x10^{-3}$ |
| *SEPT3* | 22:42347276-42419225 | 183 | 2.65 | $4.0x10^{-3}$ |
| *WBP2NL* | 22:42369729-42479460 | 205 | 2.48 | $6.6x10^{-3}$ |
| *NAGA* | 22:42429358-42491846 | 105 | 2.20 | 0.014 |
| *C22orf46* | 22:42059943-42119140 | 138 | 2.13 | 0.017 |
| *MEI1* | 22:42070503-42220460 | 353 | 2.13 | 0.017 |
| *NHP2L1* | 22:42044934-42111508 | 146 | 2.10 | 0.018 |
| *CENPM* | 22:42309725-42368168 | 159 | 2.09 | 0.018 |
| *SREBF2* | 22:42204109-42328312 | 252 | 2.07 | 0.019 |
| *TEF* | 22:41738337-41820330 | 178 | 2.04 | 0.021 |
| *FAM109B* | 22:42445255-42500445 | 105 | 2.00 | 0.023 |
| *NDUFA6* | 22:42456529-42511959 | 120 | 1.96 | 0.025 |
| *CCDC134* | 22:42171683-42247303 | 166 | 1.93 | 0.027 |
| *SMDT1* | 22:42450695-42505288 | 112 | 1.91 | 0.028 |
| *SHISA8* | 22:42282297-42335570 | 120 | 1.86 | 0.031 |
| *ZC3H7B* | 22:41672526-41781151 | 185 | 1.86 | 0.032 |
| *TNFRSF13C* | 22:42296045-42347822 | 128 | 1.85 | 0.032 |
| *TOB2* | 22:41804496-41868027 | 131 | 1.83 | 0.033 |
| *POLR3H* | 22:41896808-41965610 | 119 | 1.79 | 0.037 |
| *CSDC2* | 22:41931767-41998745 | 137 | 1.78 | 0.038 |
| *XRCC6* | 22:41992123-42085044 | 204 | 1.74 | 0.041 |
| *PMM1* | 22:41947898-42010894 | 132 | 1.72 | 0.043 |
| *ACO2* | 22:41840129-41949993 | 168 | 1.71 | 0.044 |
| *PHF5A* | 22:41830721-41889729 | 88 | 1.49 | 0.068 |
| *AL035681.1* | 22:41660388-41710686 | 76 | 1.44 | 0.075 |
| *DESI1* | 22:41969032-42042100 | 158 | 1.28 | 0.100 |
| *ARFGAP3* | 22:43167508-43279112 | 385 | 0.98 | 0.163 |
| *CYB5R3* | 22:42988846-43070574 | 229 | 0.80 | 0.212 |
| *PACSIN2* | 22:43206418-43436151 | 668 | 0.77 | 0.220 |
| *NFAM1* | 22:42751416-42853401 | 275 | 0.76 | 0.222 |
| *ATP5L2* | 22:43010809-43061607 | 169 | 0.66 | 0.254 |
| *A4GALT* | 22:43063127-43142304 | 352 | 0.57 | 0.284 |
| *SERHL2* | 22:42924623-42995388 | 173 | 0.42 | 0.337 |
| *POLDIP3* | 22:42954727-43035968 | 204 | 0.29 | 0.386 |
| *RRP7A* | 22:42880974-42940808 | 167 | -0.07 | 0.526 |
| *MCAT* | 22:43503212-43564400 | 159 | -0.29 | 0.613 |
| *TTLL1* | 22:43410522-43510434 | 338 | -0.34 | 0.631 |
| *TTLL12* | 22:43537628-43608139 | 78 | -0.36 | 0.641 |
| *TSPO* | 22:43522520-43584248 | 110 | -0.48 | 0.685 |
| *BIK* | 22:43481754-43550718 | 186 | -0.62 | 0.733 |

Location: chromosome and base pair of gene start and end based on GRCh37/hg19.
SNPs: number of variants mapped in each gene.

**Table S9. Colocalisation results (PP(H4) > 80%) between GTEx expression data and the significant locus at *TFC20*.**

| Gene | Tissue | PP(H4) |
|---|---|---|
| *CYP2D6* | Liver | 0.961 |
| *TCF20* | Skin Not Sun Exposed Suprapubic | 0.835 |
| *DESI1* | Colon Sigmoid | 0.876 |
| *OGFRP1* | Ovary | 0.995 |
| | Adipose Subcutaneous | 0.992 |
| | Breast Mammary Tissue | 0.987 |
| | Esophagus Mucosa | 0.984 |
| | Skin Not Sun Exposed Suprapubic | 0.984 |
| | Thyroid | 0.984 |
| | Pancreas | 0.978 |
| | Pituitary | 0.971 |
| | Stomach | 0.971 |
| | Cells Cultured fibroblasts | 0.967 |
| | Artery Aorta | 0.966 |
| | Whole Blood | 0.960 |
| | Adipose Visceral Omentum | 0.958 |
| | Lung | 0.957 |
| | Colon Transverse | 0.952 |
| | Prostate | 0.948 |
| | Artery Tibial | 0.943 |
| | Skin Sun Exposed Lower leg | 0.926 |
| | Small Intestine Terminal Ileum | 0.904 |
| | Colon Sigmoid | 0.845 |
| | Adrenal Gland | 0.835 |
| | Nerve Tibial | 0.813 |
| *CYP2D7* | Liver | 0.979 |
| *OLA1P1* | Brain Cerebellum | 0.969 |
| RP4-669P10.19 | Brain Hypothalamus | 0.819 |
| Z83851.4 | Thyroid | 0.945 |
| NDUFA6-DT | Brain Nucleus accumbens basal ganglia | 0.842 |
| RP1-257I20.14 | Esophagus Muscularis | 0.943 |
| | Brain Frontal Cortex BA9 | 0.860 |
| CTA-989H11.1 | Esophagus Mucosa | 0.937 |
| | Skin Sun Exposed Lower leg | 0.83 |

PP(H4): posterior probability that variants are associated with sVEGFR1 levels and gene expression by the same causal variant.

**Table S10. Annotation of missense variants in *TCF20* included in low frequency analysis on the prioritised genes.**

| Variant ID | Rs | AF | AF gnomAD | CADD | PolyPhen | MutationTaster | primateAI3D | ClinVar | Exon | HGVSp | Codons |
|---|---|---|---|---|---|---|---|---|---|---|---|
| **chr22:42564685:T:A** | rs146374327 | 0.0007 | 0.0003 | 26.1 | Possibly damaging (0.541) | D | 0.71 | Benign | 4/5 | p.Ser1953Cys | Agc/Tgc |
| **chr22:42564717:G:T** | rs144341537 | 0.0007 | 0.0002 | 24.1 | Benign (0.219) | D | 0.61 | Benign | 4/5 | p.Pro1942His | cCc/cAc |
| **chr22:42605821:T:C** | rs145292779 | 0.0015 | 0.0002 | 19 | Benign (0.046) | D | 0.36 | Like Benign Benign | 1/4 | p.Thr1831Ala | Act/Gct |
| **chr22:42607000:G:A** | rs771336001 | 0.0007 | 0.00005 | 26.8 | Possibly damaging (0.731) | D | 0.50 | VUS | 1/4 | p.Arg1438Cys | Cgt/Tgt |
| **chr22:42607018:T:C** | rs779029341 | 0.0007 | - | 14.77 | Benign (0) | N | 0.40 | Benign | 1/4 | p.Arg1432Gly | Agg/Ggg |
| **chr22:42607338:C:T** | rs17002888 | 0.0044 | 0.002 | 8.192 | Benign (0) | N | 0.27 | Benign | 1/4 | p.Ser1325Asn | aGt/aAt |
| **chr22:42607438:C:T** | rs775825597 | 0.0007 | 0.000009 | 0.202 | Benign (0.001) | N | 0.23 | VUS | 1/4 | p.Ala1292Thr | Gct/Act |
| **chr22:42607817:C:T** | rs17002890 | 0.0029 | 0.0005 | 22.6 | Benign (0.007) | N | 0.40 | Benign | 1/4 | p.Met1165Ile | atG/atA |
| **chr22:42608005:C:T** | rs762509400 | 0.0007 | 0.000009 | 14.43 | Benign (0) | N | 0.15 | - | 1/4 | p.Gly1103Ser | Ggt/Agt |
| **chr22:42608191:C:G** | rs1450225029 | 0.0007 | 0.000009 | 25.6 | Possibly damaging (0.756) | D | 0.66 | VUS | 1/4 | p.Glu1041Gln | Gag/Cag |
| **chr22:42608224:G:T** | rs200309070 | 0.0007 | 0.00002 | 19.55 | Benign (0.029) | D | 0.52 | - | 1/4 | p.Pro1030Thr | Cct/Act |
| **chr22:42608742:C:G** | rs1453276331 | 0.0007 | 0.000009 | 23.1 | Possibly damaging (0.741) | D | 0.39 | - | 1/4 | p.Gly857Ala | gGg/gCg |
| **chr22:42609192:T:C** | rs764736571 | 0.0007 | 0.00006 | 25.9 | Probably damaging (0.964) | D | 0.66 | VUS | 1/4 | p.Tyr707Cys | tAt/tGt |
| **chr22:42610139:C:A** | rs746200891 | 0.0007 | - | 23.2 | Possibly damaging (0.655) | N | 0.48 | - | 1/4 | p.Gln391His | caG/caT |
| **chr22:42610929:T:C** | rs748499160 | 0.0007 | 0.00006 | 12.29 | Benign (0.003) | N | 0.25 | VUS Like Benign | 1/4 | p.Asn128Ser | aAt/aGt |

Variant ID: chromosome, base pair, reference and alternative allele based on GRCh37/hg19; AF: allele frequency; AF gnomAD: allele frequency in non-Finish Europeans; D: disease causing, probably deleterious; N: polymorphism probably harmless; VUS, variant of uncertain significance. Gene damage index for *TCF20*: 7.94.

**Table S11. Results of the best PGS prediction models for sepsis-associated ARDS and ICU mortality in the sVEGFR1 levels GWAS.**

| | Sepsis induced ARDS | | | | ICU mortality | | | |
|---|---|---|---|---|---|---|---|---|
| **GWAS** | ***p*-value threshold** | **SNPs** | ***p*-value** | **OR[95%CI]** | ***p*-value threshold** | **SNPs** | ***p*-value** | **OR[95%CI]** |
| T1 (N=225) | $6.56x10^{-5}$ | 89 | 0.072 | 0.85[0.71-1.01] | $1.82x10^{-3}$ | 2,116 | $2.61x10^{-5}$ | 0.65[0.54-0.80] |
| T2 (N=169) | $1.05x10^{-4}$ | 118 | 0.054 | 1.19[1.00-1.43] | $6.85x10^{-6}$ | 6 | 0.033 | 1.23[1.02-1.49] |
| T7 (N=108) | $2.84x10^{-3}$ | 3,094 | $1.04x10^{-4}$ | 0.70[0.58-0.84] | 0.100 | 70,598 | 0.125 | 0.86[0.71-1.04] |

T1: sVEGFR1 levels obtained within 24 hours after sepsis diagnosis; T2: sVEGFR1 levels obtained within 48-72 hours after sepsis diagnosis; T7: sVEGFR1 levels obtained 7 days after sepsis diagnosis. Sepsis-associated ARDS: 338 controls with sepsis and 218 cases with ARDS-sepsis. ICU mortality: 369 surviving controls with sepsis and 176 non-survival patients with sepsis who died in ICU after diagnosis of sepsis. CI: confident interval; OR: odd ratio; *p*-value threshold: *p*-value threshold at which the most significant model is obtained; SNPs: number of variants included in the score.

**Table S15. Results from the Mendelian Randomization analysis evaluating whether VEGFA and VEGFR1 levels have a causal effect on ARDS susceptibility.**

| Exposure/outcome | IVs | Model | *p*-value | OR[95%CI] | Complementary test |
|---|---|---|---|---|---|
| **VEGFA/ARDS-Sepsis** | 36 | IVW | 0.992 | 1.00[0.59-1.70] | Q-statistic *p*=0.478 |
| | | MR Egger | 0.940 | 0.97[0.49-1.94] | Q-statistic *p*=0.431 Egger intercept=0.004 (SE=0.03; *p*=0.895) |
| | | MR-PRESSO | 0.993 | 1.00[0.59-1.70] | Global test *p*=0.528 |
| | | Simple mode | 0.351 | 0.42[0.07-2.55] | |
| | | Weighted median | 0.782 | 1.09[0.58-2.08] | |
| | | Weighted mode | 0.860 | 1.05[0.60-1.84] | |
| | 1,635 | MR-RAPS | 0.402 | 0.85[0.58-1.25] | |
| **VEGFR1/ARDS-Sepsis** | 16 | IVW | 0.924 | 0.87[0.05-14.68] | Q-statistic *p*=0.933 |
| | | MR Egger | 0.434 | 0.16[0.002-14.40] | Q-statistic *p*=0.940 Egger intercept=0.055 (SE=0.06; *p*=0.356) |
| | | MR-PRESSO | 0.896 | 0.87[0.11-6.64] | Global test *p*=0.931 |
| | | Simple mode | 0.865 | 0.57[0.001-355.84] | |
| | | Weighted median | 0.528 | 0.33[0.01-9.98] | |
| | | Weighted mode | 0.645 | 0.42[0.01-15.08] | |
| | 1,636 | MR-RAPS | 0.729 | 1.20[0.43-3.29] | |

CI: confidence interval; IV: instrumental variable; IVW: inverse variance-weighted; OR: Odd Ratio; SE: standard error.

**Table S16. Results from the Mendelian Randomization analysis evaluating whether ARDS susceptibility has a causal effect on VEGFA and VEGFR1 levels.**

| Exposure/outcome | IVs | Model | *p*-value | Beta | SE | Complementary test |
|---|---|---|---|---|---|---|
| **ARDS-Sepsis/VEGFA** | 42 | IVW | 0.487 | 0.001 | 0.002 | Q-statistic $p$=3.97x10$^{-8}$ |
| | | MR Egger | 0.631 | -0.004 | 0.008 | Q-statistic $p$=3.43x10$^{-8}$ Egger intercept=0.003 (SE=0.005, *p*=0.520) |
| | | MR-PRESSO | 0.889* | 0.0002 | 0.001 | Global test *p*<0.001 |
| | | Simple mode | 0.373 | 0.003 | 0.003 | |
| | | Weighted median | 0.345 | 0.001 | 0.002 | |
| | | Weighted mode | 0.356 | 0.003 | 0.003 | |
| | 1,443 | MR-RAPS | 0.732 | 0.0001 | 0.0003 | |
| **ARDS-Sepsis/VEGFR1** | 42 | IVW | 0.168 | -0.0007 | 0.0005 | Q-statistic *p*=0.256 |
| | | MR Egger | 0.943 | 0.0002 | 0.002 | Q-statistic *p*=0.228 Egger intercept=-0.0006 (SE=0.0015, *p*=0.704) |
| | | MR-PRESSO | 0.094 | -0.0008 | 0.0005 | Global test *p*=0.237 |
| | | Simple mode | 0.960 | 0.0001 | 0.001 | |
| | | Weighted median | 0.838 | -0.0001 | 0.0006 | |
| | | Weighted mode | 0.988 | 0.00002 | 0.001 | |
| | 1,444 | MR-RAPS | 0.746 | -0.00004 | 0.0001 | |

IV: instrumental variable; IVW: inverse variance-weighted; SE: standard error. MR-PRESSO global test *p*-value after excluding an outlier.

**Figure S1. Scheme of the Mendelian randomisation approach to assess the causal effect of VEGFA (A) and VEGFR1 (B) on ARDS susceptibility.**

A.

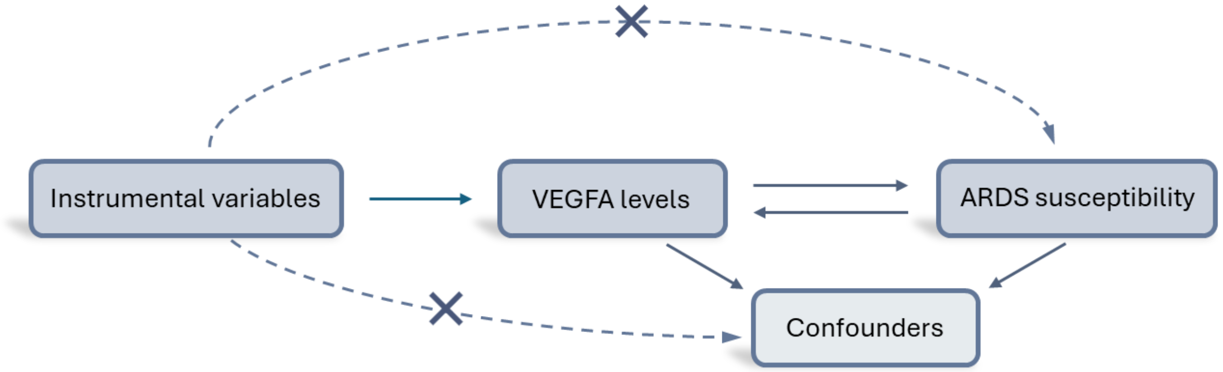


B.

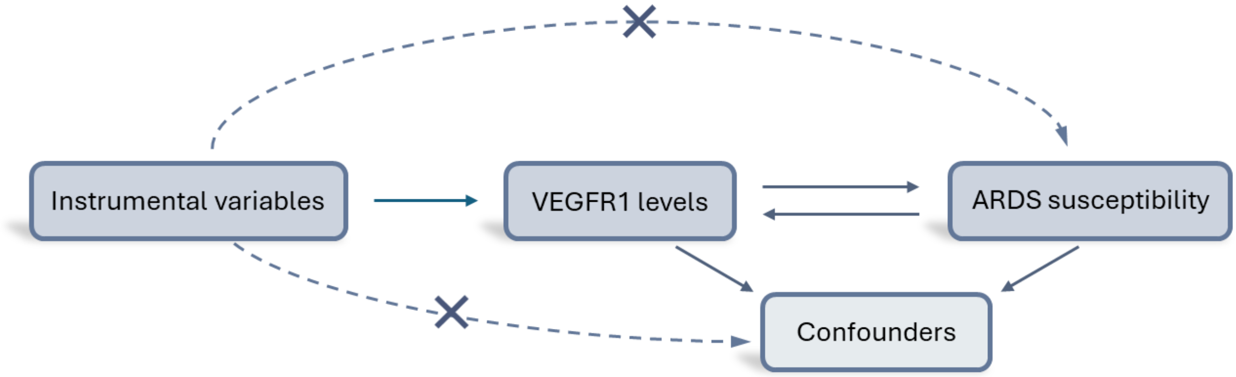

**Figure S2. Quantile-Quantile (Q-Q) plots.** sVEGFR1 levels (pg/ml) observed versus expected after normalisation at T1, T2 and T7. The p-values show the results derived from the Lilliefors test. T1: sVEGFR1 levels obtained within 24 hours after sepsis diagnosis; T2: sVEGFR1 levels obtained within 48-72 hours after sepsis diagnosis; T7: sVEGFR1 levels obtained 7 days after sepsis diagnosis.

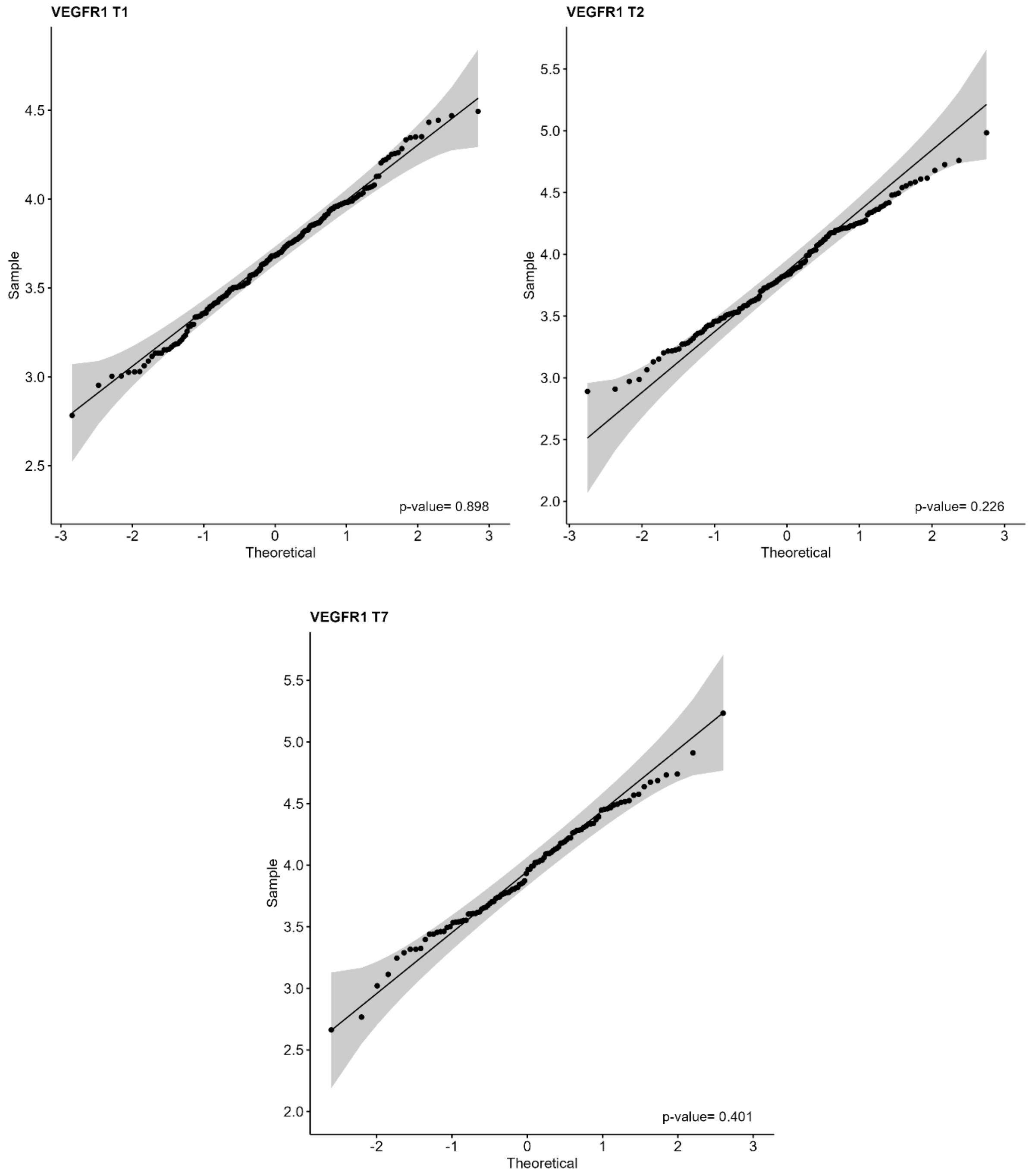

**Figure S3. Box plot of normalised serum sVEGFR1 levels (pg/ml) at T1, T2 and, T7.** The *p*-values show the results derived from the t-test. T1: sVEGFR1 levels obtained within 24 hours after sepsis diagnosis; T2: sVEGFR1 levels obtained within 48-72 hours after sepsis diagnosis; T7: sVEGFR1 levels obtained 7 days after sepsis diagnosis.

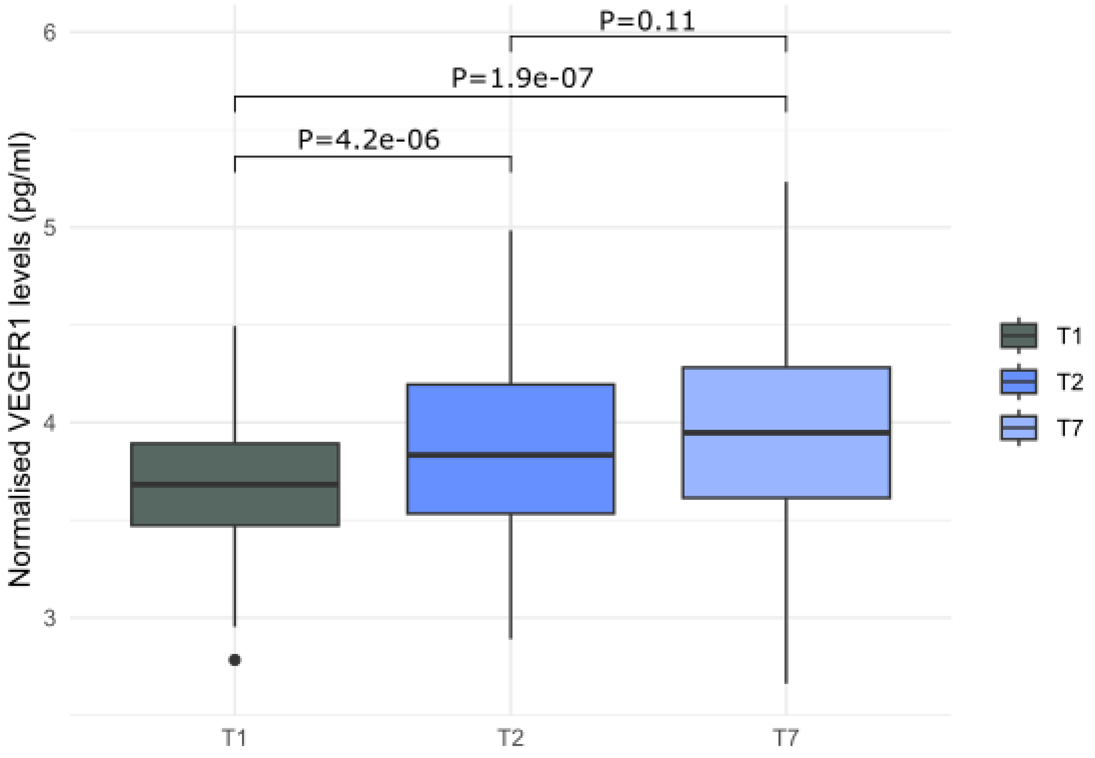

**Figure S4. Quantile-Quantile (Q-Q) plots.** Observed versus expected -log10 *p*-values for the GWAS results of serum sVEGFR1 levels at T1, T2, and T7. T1: sVEGFR1 levels obtained within 24 hours after sepsis diagnosis; T2: sVEGFR1 levels obtained within 48-72 hours after sepsis diagnosis; T7: sVEGFR1 levels obtained 7 days after sepsis diagnosis.

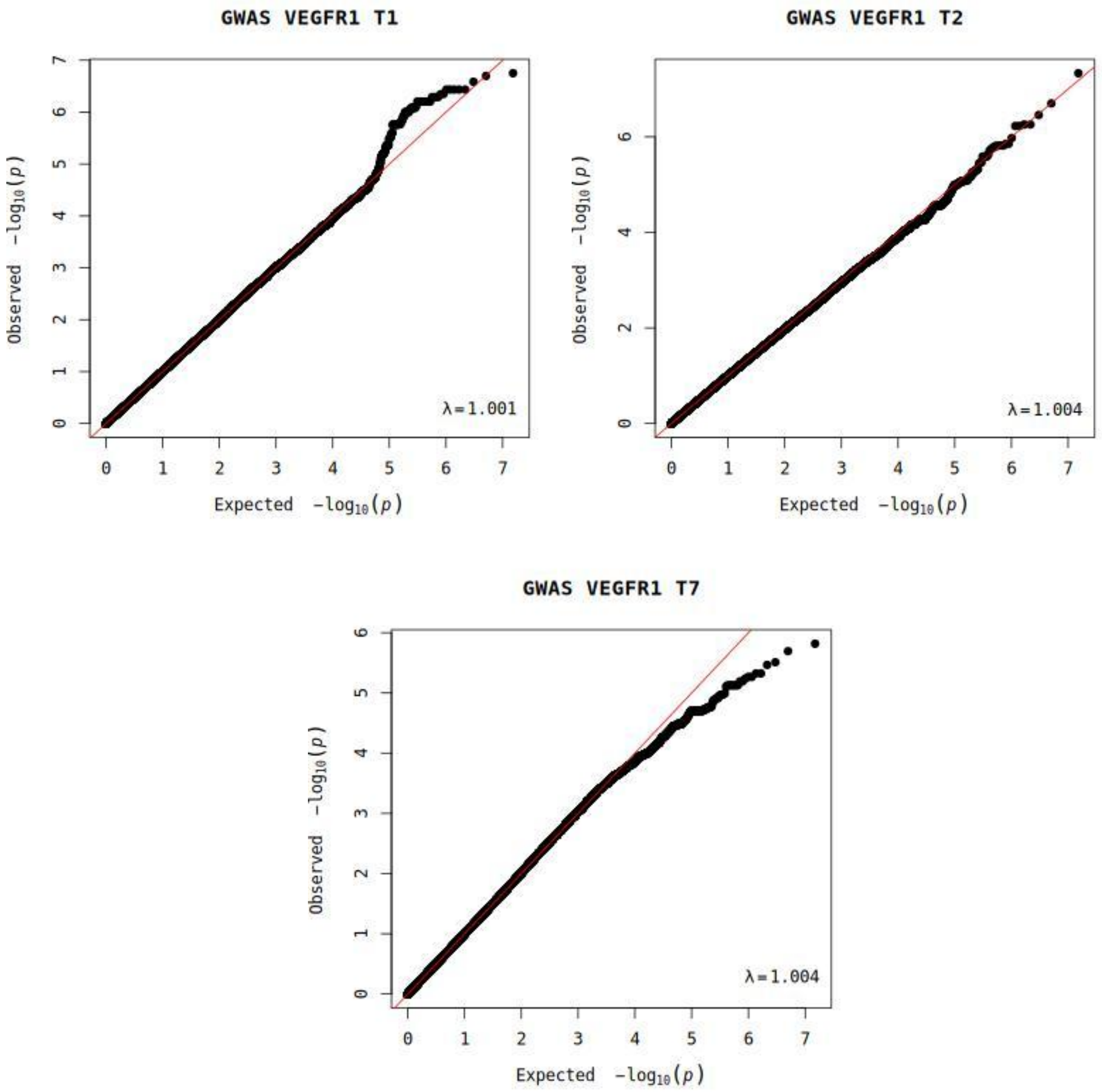

**Figure S5. Manhattan plot of the genome-wide association study results for sVEGFR1 levels at T1 (N=225), T2 (N=169), and T7 (N=108).** The x-axis represents the chromosomal positions (GRCh37/hg19) and the y-axis shows the -log10(*p*-value). The horizontal dash line represents the genome-wide significance threshold (*p*-value=$5.0\times10^{-8}$). T1: sVEGFR1 levels obtained within 24 hours after sepsis diagnosis; T2: sVEGFR1 levels obtained within 48-72 hours after sepsis diagnosis; T7: sVEGFR1 levels obtained 7 days after sepsis diagnosis.

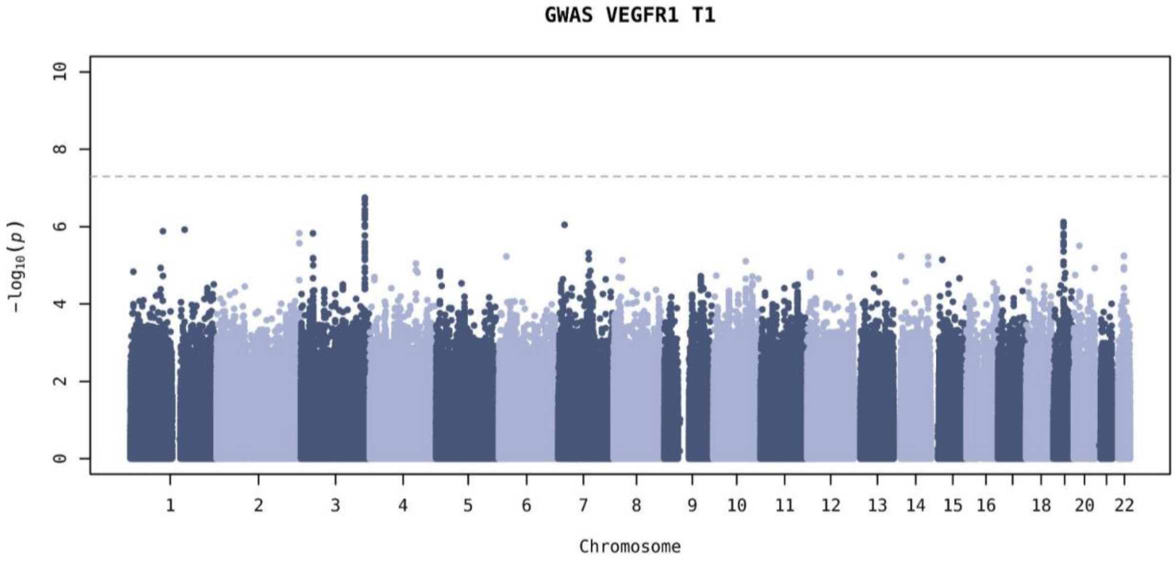


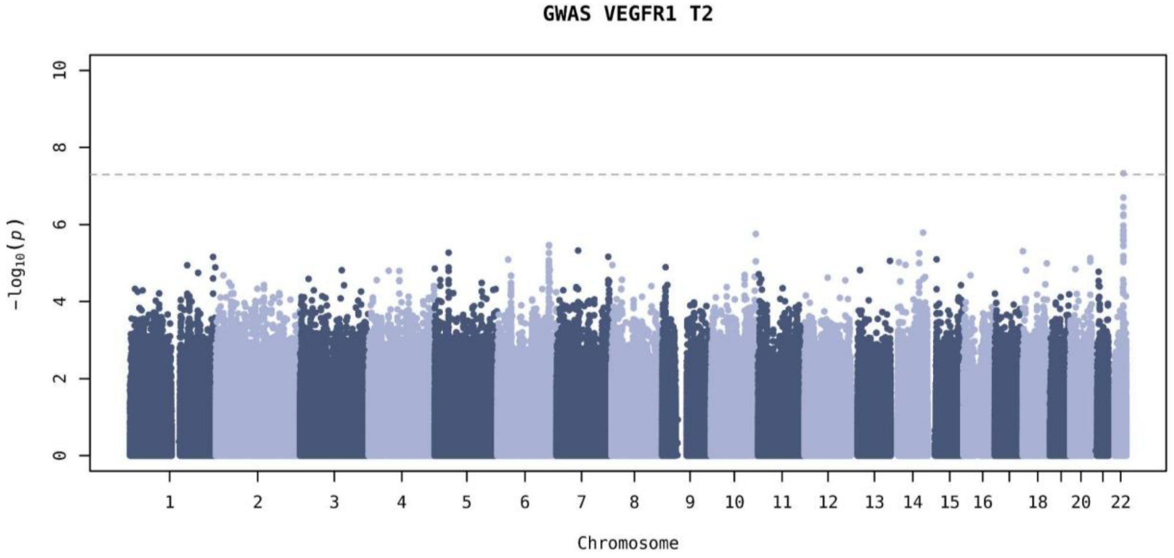

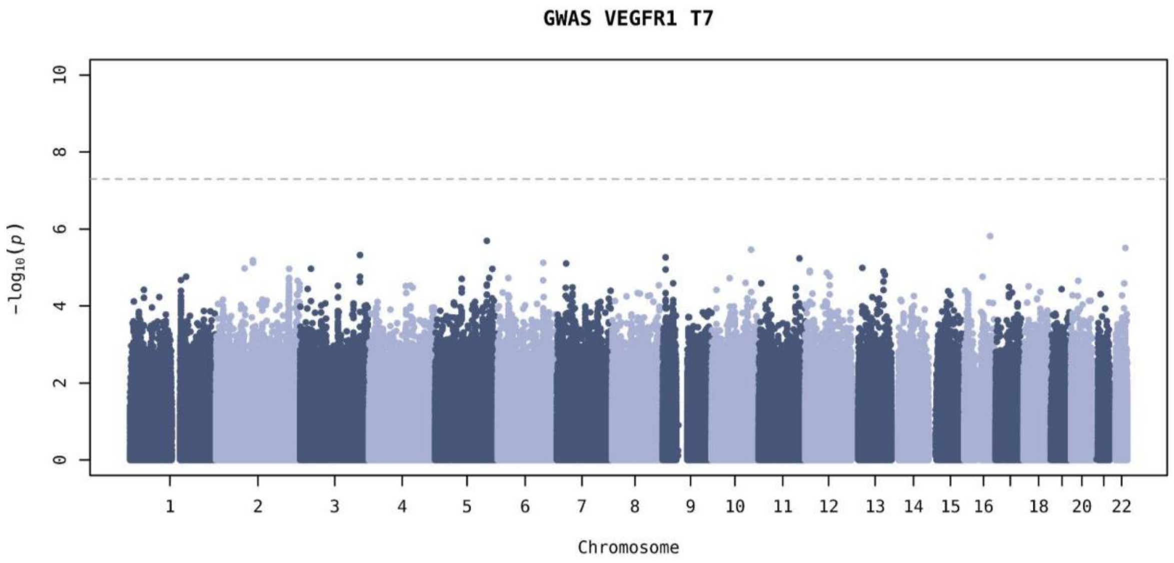


**Figure S6. Regional plot from the GWAS of sVEGFR1 levels at T2 for the significant locus at 22q13.2.** The x-axis represents the chromosomal positions (GRCh37/hg19), and the y-axis shows the -log10(*p*-value). The horizontal dashed line represents the genome-wide significance threshold (p-value=$5.0\times10^{-8}$). The sVEGFR1 significant pQTL (rs134871) is highlighted in orange and the pQTLs included in the credible set with 95% of confidence are in green. T2: sVEGFR1 levels obtained within 48-72 hours after sepsis diagnosis.

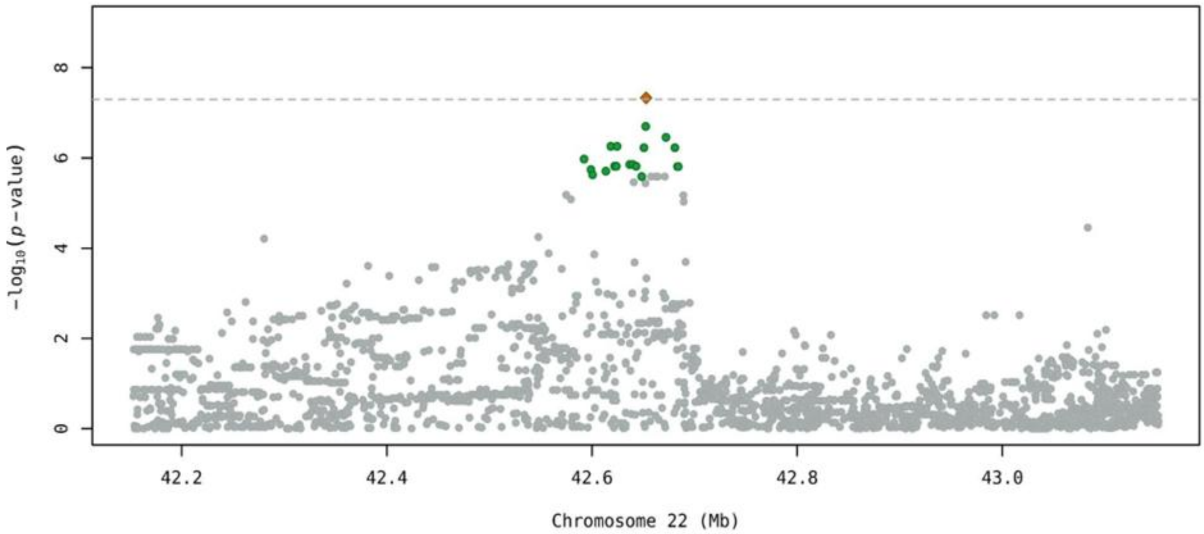

**Figure S7. Mirror plots of the significant locus at 22q13.2 for GWAS of sVEGFR1 levels at T2 vs. eQTL results.** The x-axis represents the chromosomal positions (GRCh38/hg38). The top y-axis shows the -log(*p*-value) of the GWAS of sVEGFR1 at T2, the bottom y-axis shows the -log(*p*-value) from the GTEx v8 database. The horizontal red line represents the genome-wide significance threshold (*p*-value=$5.0\times10^{-8}$). The sVEGFR1 significant pQTL (rs134871) is highlighted in blue the colour of the remaining variants represents their linkage disequilibrium with the significant variant (red: $r2 \geq 0.8$, orange: $0.6 \leq r2 < 0.8$, yellow: $0.4 \leq r2 < 0.6$, light yellow: $0.2 \leq r2 < 0.4$, and grey: $r2 < 0.2$). Diamonds indicate the variants in the 95% credible set obtained with *Corrcoverage*. T2: sVEGFR1 levels obtained within 48-72 hours after sepsis diagnosis.

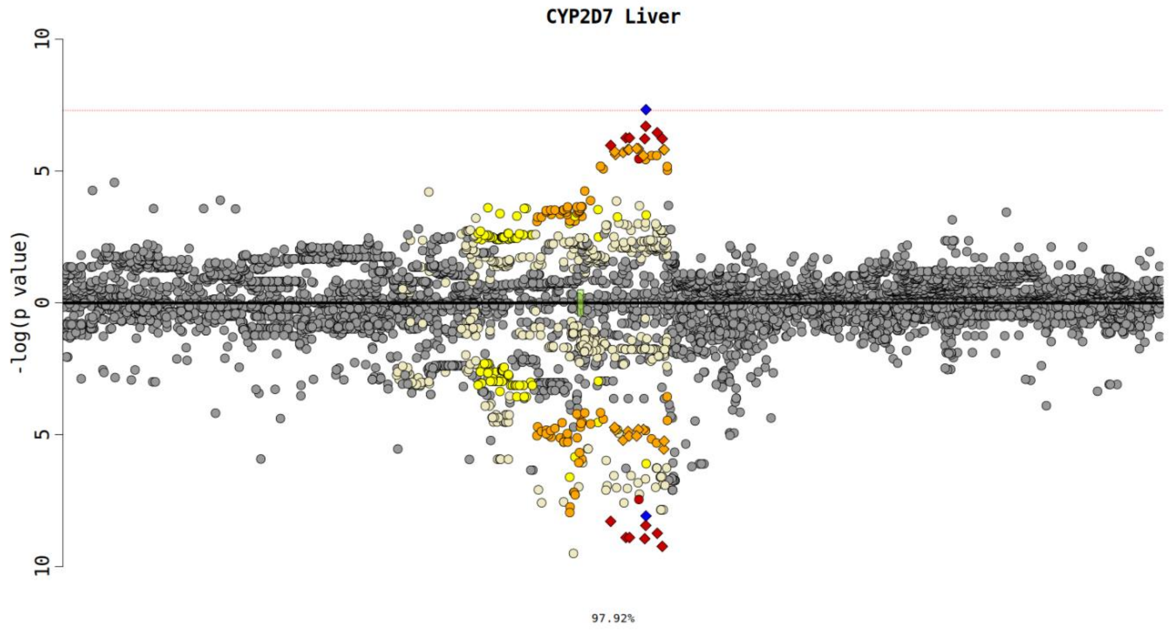


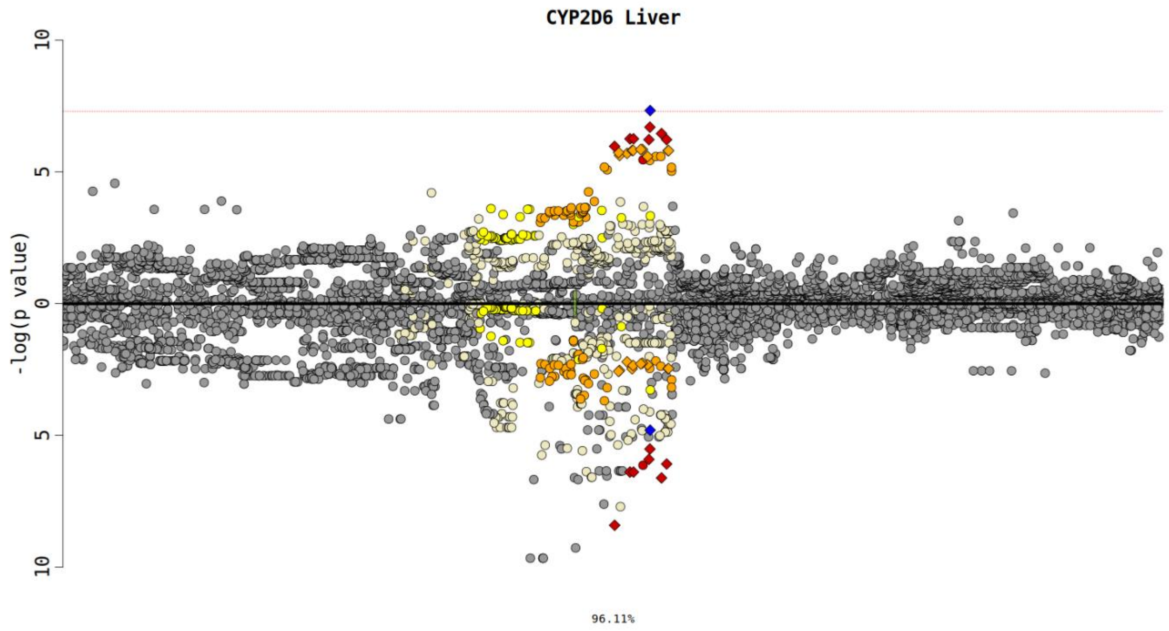

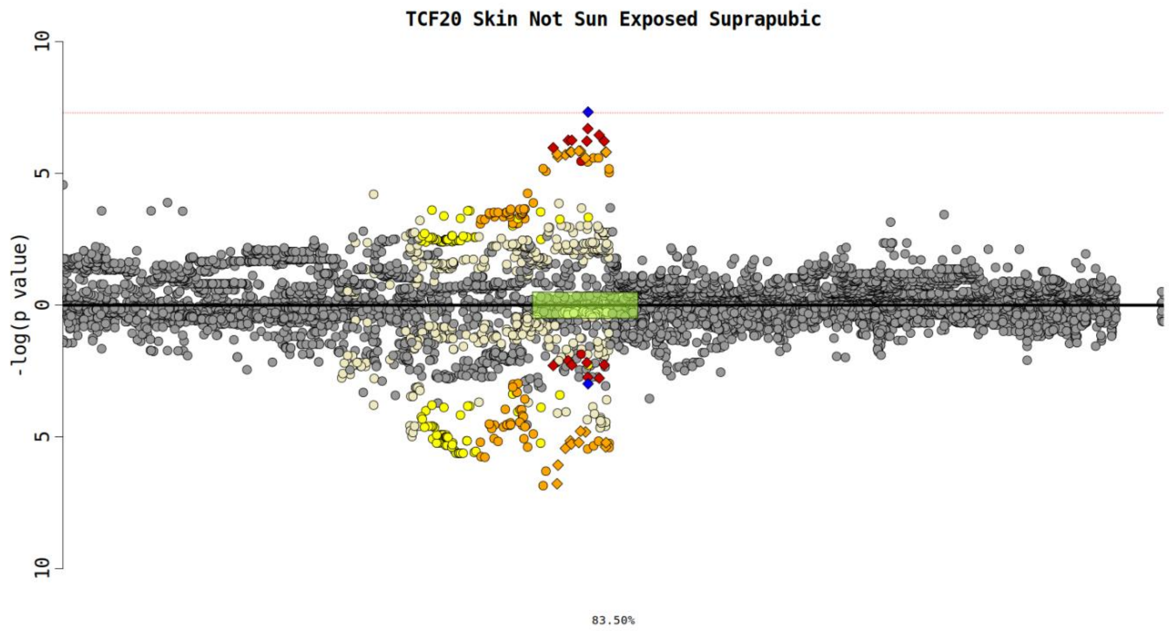


**Figure S8. Manhattan plot of the genome-wide association study results of VEGFR1 levels from UKBB (N=46,836).** The x-axis represents the chromosomal positions (GRCh37/hg19), and the y-axis shows the -log10(*p*-value). The horizontal dash line represents the genome-wide significance threshold (*p*-value=$5.0\times10^{-8}$).

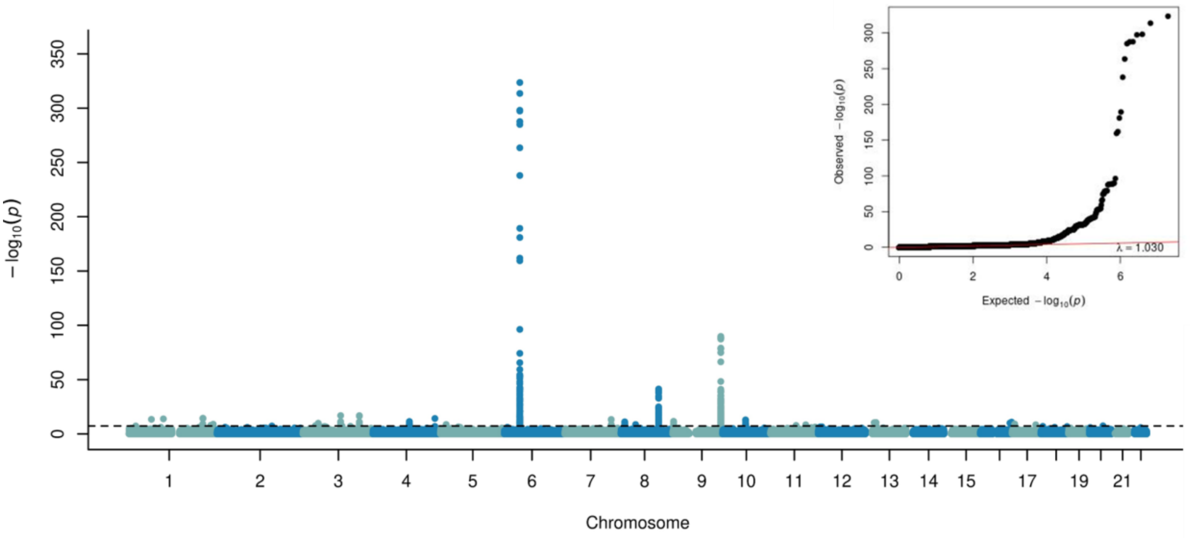

**Figure S9. Manhattan plot of the genome-wide association study results of VEGFA levels from UKBB (N=47,498).** The x-axis represents the chromosomal positions (GRCh37/hg19), and the y-axis shows the -log10(*p*-value). The horizontal dash line represents the genome-wide significance threshold (*p*-value=$5.0\times10^{-8}$).

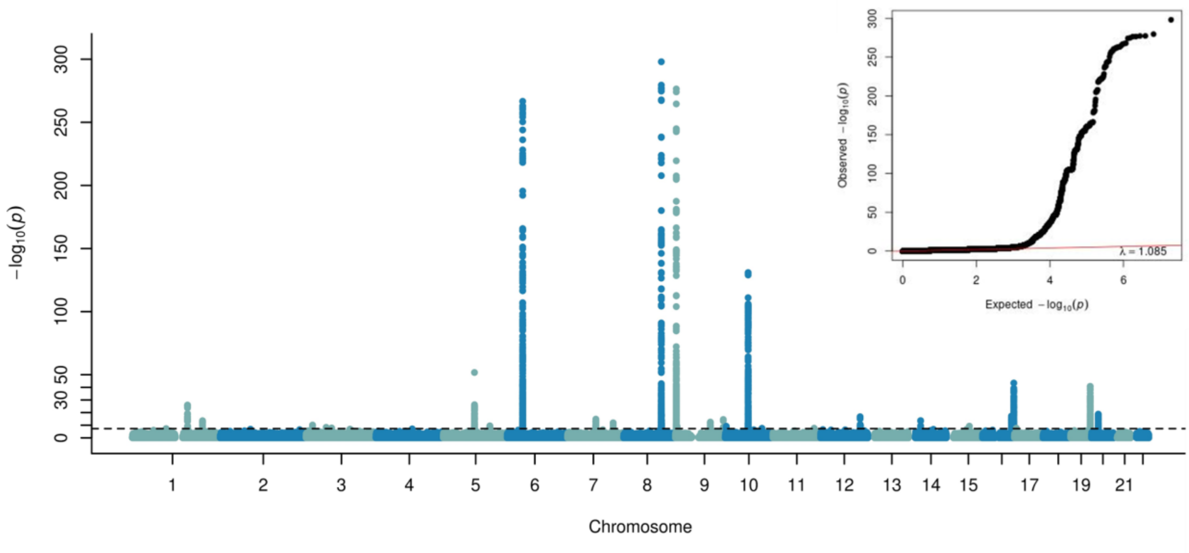
