## Supplementary Table S12 for "Genetic regulation of the vascular endothelial growth factor receptor 1 during sepsis and association with ARDS susceptibility"

**Table S12. Significant results of gene set enrichment analysis on the DSigDB dataset for the list of common genes obtained from the best PGS models at T1 and T7.**

| Term | Overlap | P-value | Adjusted P-value | Odds Ratio | Combined Score |
|---|---|---|---|---|---|
| trichostatin A CTD 00000660 | 487/3584 | 2.67E-61 | 9.81E-58 | 2.83 | 395.27 |
| Retinoic acid CTD 00006918 | 484/4258 | 2.06E-37 | 3.78E-34 | 2.20 | 186.10 |
| VALPROIC ACID CTD 00006977 | 771/8312 | 8.14E-33 | 9.98E-30 | 1.96 | 144.94 |
| benzo[a]pyrene CTD 00005488 | 432/4424 | 2.26E-18 | 2.08E-15 | 1.73 | 70.21 |
| MS-275 PC3 UP | 82/431 | 6.95E-18 | 5.11E-15 | 3.39 | 133.98 |
| arsenite CTD 00000779 | 168/1300 | 7.55E-17 | 4.63E-14 | 2.20 | 81.65 |
| 8-Bromo-cAMP, Na CTD 00007044 | 104/652 | 1.17E-16 | 6.15E-14 | 2.76 | 101.14 |
| progesterone CTD 00006624 | 217/1915 | 4.36E-15 | 2.00E-12 | 1.91 | 63.23 |
| Caspan CTD 00000180 | 116/808 | 5.13E-15 | 2.09E-12 | 2.44 | 80.27 |
| Tetradioxin CTD 00006848 | 362/3768 | 6.46E-14 | 2.37E-11 | 1.64 | 49.80 |
| melphalan CTD 00006262 | 54/279 | 1.60E-12 | 5.34E-10 | 3.41 | 92.68 |
| raloxifene CTD 00007367 | 95/686 | 1.39E-11 | 4.26E-09 | 2.31 | 57.83 |
| irinotecan MCF7 DOWN | 147/1272 | 5.16E-11 | 1.46E-08 | 1.90 | 45.09 |
| tamoxifen CTD 00006827 | 104/802 | 7.58E-11 | 1.99E-08 | 2.15 | 50.03 |
| estradiol CTD 00005920 | 389/4336 | 1.06E-10 | 2.59E-08 | 1.51 | 34.63 |
| POTASSIUM CHROMATE CTD 00001284 | 198/1897 | 1.67E-10 | 3.84E-08 | 1.71 | 38.60 |
| (-)-Epigallocatechin gallate CTD 00002033 | 215/2114 | 2.49E-10 | 5.39E-08 | 1.67 | 36.95 |
| vorinostat PC3 UP | 64/416 | 4.74E-10 | 9.68E-08 | 2.59 | 55.54 |
| irinotecan PC3 DOWN | 119/999 | 6.82E-10 | 1.32E-07 | 1.95 | 41.20 |
| AFLATOXIN B1 CTD 00007128 | 289/3081 | 8.09E-10 | 1.49E-07 | 1.55 | 32.39 |
| calcitriol CTD 00005558 | 198/1958 | 2.36E-09 | 4.13E-07 | 1.65 | 32.77 |
| formaldehyde CTD 00006001 | 294/3192 | 3.77E-09 | 6.30E-07 | 1.51 | 29.35 |
| parthenolide MCF7 DOWN | 32/162 | 3.30E-08 | 5.28E-06 | 3.46 | 59.58 |
| rifabutin PC3 UP | 29/151 | 2.69E-07 | 4.13E-05 | 3.33 | 50.43 |
| trichostatin A PC3 UP | 56/415 | 5.44E-07 | 7.75E-05 | 2.20 | 31.80 |
| 17-Ethynyl estradiol CTD 00005932 | 45/304 | 5.48E-07 | 7.75E-05 | 2.45 | 35.30 |
| thiostrepton MCF7 DOWN | 10/22 | 5.80E-07 | 7.90E-05 | 11.59 | 166.45 |
| piperlongumine HL60 DOWN | 53/389 | 8.08E-07 | 1.06E-04 | 2.23 | 31.25 |
| BW-B70C MCF7 UP | 14/47 | 1.58E-06 | 1.95E-04 | 5.91 | 78.99 |
| TERBUFOS CTD 00000658 | 82/715 | 1.59E-06 | 1.95E-04 | 1.84 | 24.57 |
| camptothecin PC3 DOWN | 147/1494 | 1.73E-06 | 2.05E-04 | 1.57 | 20.83 |
| lomustine PC3 DOWN | 22/109 | 3.08E-06 | 3.48E-04 | 3.53 | 44.85 |
| METHAMPHETAMINE CTD 00006286 | 21/101 | 3.13E-06 | 3.48E-04 | 3.67 | 46.49 |
| doxorubicin PC3 DOWN | 107/1023 | 3.48E-06 | 3.77E-04 | 1.67 | 20.94 |
| Fonofos CTD 00005884 | 80/712 | 4.67E-06 | 4.82E-04 | 1.80 | 22.04 |
| anisomycin PC3 UP | 96/899 | 4.76E-06 | 4.82E-04 | 1.70 | 20.85 |
| daunorubicin PC3 DOWN | 50/381 | 4.85E-06 | 4.82E-04 | 2.13 | 26.05 |
| parathion CTD 00006472 | 85/776 | 6.40E-06 | 6.20E-04 | 1.75 | 20.88 |

| | | | | | |
|---|---|---|---|---|---|
| 8-azaguanine HL60 UP | 54/428 | 6.67E-06 | 6.29E-04 | 2.04 | 24.26 |
| cicloheximide PC3 UP | 49/378 | 8.45E-06 | 7.77E-04 | 2.10 | 24.50 |
| rosiglitazone CTD 00003139 | 52/411 | 9.09E-06 | 8.15E-04 | 2.04 | 23.70 |
| Torcetrapib CTD 00004291 | 44/327 | 9.38E-06 | 8.21E-04 | 2.19 | 25.31 |
| pyrvinium MCF7 UP | 28/170 | 9.90E-06 | 8.46E-04 | 2.76 | 31.81 |
| Methaneseleninic acid CTD 00000412 | 51/402 | 1.02E-05 | 8.55E-04 | 2.05 | 23.52 |
| MG-262 PC3 DOWN | 46/360 | 2.28E-05 | 0.0019 | 2.06 | 22.02 |
| fisetin PC3 DOWN | 62/541 | 3.02E-05 | 0.0024 | 1.83 | 19.00 |
| camptothecin MCF7 DOWN | 142/1513 | 3.03E-05 | 0.0024 | 1.48 | 15.42 |
| NICKEL CHLORIDE CTD 00001064 | 49/399 | 3.57E-05 | 0.0027 | 1.97 | 20.16 |
| dexamethasone CTD 00005779 | 51/421 | 3.60E-05 | 0.0027 | 1.94 | 19.85 |
| cyclosporin A CTD 00007121 | 387/4825 | 4.09E-05 | 0.0030 | 1.29 | 13.00 |
| Irinotecan hydrochloride CTD 00002224 | 63/565 | 5.94E-05 | 0.0043 | 1.77 | 17.22 |
| Silica CTD 00006678 | 91/898 | 6.21E-05 | 0.0043 | 1.60 | 15.48 |
| valsartan CTD 00002971 | 14/63 | 6.23E-05 | 0.0043 | 3.98 | 38.52 |
| Bisphenol A CTD 00000312 | 120/1262 | 7.43E-05 | 0.0051 | 1.50 | 14.22 |
| DICHLOROMETHANE CTD 00006313 | 12/49 | 7.57E-05 | 0.0051 | 4.51 | 42.81 |
| oxygen CTD 00006454 | 59/524 | 7.69E-05 | 0.0051 | 1.79 | 16.93 |
| niclosamide PC3 DOWN | 9/29 | 8.24E-05 | 0.0053 | 6.25 | 58.79 |
| puromycin MCF7 UP | 33/242 | 9.06E-05 | 0.0057 | 2.21 | 20.58 |
| rofecoxib CTD 00003631 | 18/99 | 1.01E-04 | 0.0063 | 3.10 | 28.51 |
| parthenolide PC3 DOWN | 26/174 | 1.10E-04 | 0.0068 | 2.45 | 22.37 |
| TERT-BUTYL HYDROPEROXIDE CTD 00007349 | 125/1341 | 1.24E-04 | 0.0075 | 1.46 | 13.16 |
| atovaquone MCF7 DOWN | 43/354 | 1.34E-04 | 0.0080 | 1.94 | 17.30 |
| LY 294002 CTD 00003061 | 31/227 | 1.42E-04 | 0.0083 | 2.21 | 19.60 |
| LY-294002 HL60 DOWN | 12/53 | 1.70E-04 | 0.0098 | 4.07 | 35.32 |
| luteolin HL60 DOWN | 37/293 | 1.77E-04 | 0.0099 | 2.02 | 17.50 |
| puromycin PC3 UP | 60/552 | 1.78E-04 | 0.0099 | 1.72 | 14.82 |
| daunorubicin MCF7 DOWN | 120/1290 | 1.85E-04 | 0.0101 | 1.46 | 12.53 |
| withaferin A MCF7 DOWN | 9/32 | 1.92E-04 | 0.0103 | 5.44 | 46.51 |
| phenoxybenzamine PC3 DOWN | 18/104 | 1.93E-04 | 0.0103 | 2.92 | 24.94 |
| trichostatin A ssMCF7 DOWN | 24/161 | 2.07E-04 | 0.0109 | 2.45 | 20.75 |
| N-(undeca-2,4,7-trienylideneamino)nitrous amide CTD 00001653 | 13/62 | 2.11E-04 | 0.0109 | 3.69 | 31.24 |
| Decitabine CTD 00000750 | 159/1800 | 2.17E-04 | 0.0111 | 1.38 | 11.67 |
| clomiphene CTD 00005686 | 8/26 | 2.20E-04 | 0.0111 | 6.17 | 51.97 |
| thioridazine | 10/40 | 2.48E-04 | 0.0123 | 4.63 | 38.45 |
| alprostadil HL60 UP | 47/409 | 2.50E-04 | 0.0123 | 1.82 | 15.11 |
| gossypol HL60 UP | 14/72 | 2.82E-04 | 0.0134 | 3.36 | 27.46 |
| risperidone BOSS | 11/48 | 2.82E-04 | 0.0134 | 4.13 | 33.78 |
| levonorgestrel HL60 DOWN | 37/300 | 2.84E-04 | 0.0134 | 1.97 | 16.09 |
| norgestrel CTD 00006422 | 28/205 | 2.91E-04 | 0.0135 | 2.21 | 18.00 |

| | | | | | |
|---|---|---|---|---|---|
| risperidone | 8/27 | 2.94E-04 | 0.0135 | 5.85 | 47.53 |
| TRICHLOROETHYLENE CTD 00006932 | 15/81 | 3.03E-04 | 0.0137 | 3.16 | 25.63 |
| 8-azaguanine PC3 UP | 39/323 | 3.05E-04 | 0.0137 | 1.92 | 15.58 |
| mebendazole HL60 UP | 41/346 | 3.21E-04 | 0.0142 | 1.88 | 15.15 |
| Arsenenous acid CTD 00000922 | 118/1283 | 3.26E-04 | 0.0143 | 1.44 | 11.54 |
| Bexarotene CTD 00003225 | 16/91 | 3.58E-04 | 0.0155 | 2.97 | 23.57 |
| tyrphostin AG-825 MCF7 DOWN | 51/462 | 3.68E-04 | 0.0157 | 1.74 | 13.78 |
| Enterolactone CTD 00001393 | 93/971 | 3.78E-04 | 0.0160 | 1.50 | 11.80 |
| scriptaid PC3 UP | 68/664 | 3.84E-04 | 0.0161 | 1.61 | 12.64 |
| resveratrol CTD 00002483 | 142/1601 | 4.04E-04 | 0.0166 | 1.39 | 10.82 |
| ETHYLBENZENE CTD 00000178 | 11/50 | 4.12E-04 | 0.0166 | 3.92 | 30.56 |
| O-XYLENE CTD 00001228 | 11/50 | 4.12E-04 | 0.0166 | 3.92 | 30.56 |
| Vatalanib succinate TTD 00011775 | 5/11 | 4.55E-04 | 0.0180 | 11.55 | 88.88 |
| 172889-27-9 BOSS | 15/84 | 4.56E-04 | 0.0180 | 3.03 | 23.28 |
| IRON CTD 00006166 | 25/180 | 4.66E-04 | 0.0181 | 2.25 | 17.27 |
| Chromium(II) chloride CTD 00000877 | 13/67 | 4.70E-04 | 0.0181 | 3.35 | 25.66 |
| valproic acid MCF7 DOWN | 19/121 | 4.72E-04 | 0.0181 | 2.60 | 19.88 |
| olanzapine | 8/29 | 5.04E-04 | 0.0191 | 5.29 | 40.16 |
| staurosporine MCF7 DOWN | 66/649 | 5.62E-04 | 0.0210 | 1.59 | 11.92 |
| strophanthidin PC3 UP | 50/459 | 5.67E-04 | 0.0210 | 1.72 | 12.82 |
| scriptaid CTD 00003819 | 12/60 | 5.77E-04 | 0.0212 | 3.48 | 25.92 |
| niclosamide MCF7 UP | 19/123 | 5.82E-04 | 0.0212 | 2.55 | 18.96 |
| retinol CTD 00006991 | 15/86 | 5.90E-04 | 0.0213 | 2.94 | 21.86 |
| meclofenoxate HL60 DOWN | 71/713 | 6.37E-04 | 0.0226 | 1.56 | 11.46 |
| Pentabromodiphenyl ether CTD 00003077 | 46/415 | 6.39E-04 | 0.0226 | 1.75 | 12.86 |
| quercetin CTD 00006679 | 256/3158 | 6.76E-04 | 0.0237 | 1.27 | 9.27 |
| menadione PC3 DOWN | 24/175 | 7.19E-04 | 0.0250 | 2.22 | 16.05 |
| Enzacamene CTD 00001812 | 5/12 | 7.38E-04 | 0.0253 | 9.90 | 71.41 |
| testosterone CTD 00006844 | 113/1247 | 7.49E-04 | 0.0255 | 1.41 | 10.16 |
| 15-delta prostaglandin J2 MCF7 DOWN | 9/38 | 7.73E-04 | 0.0256 | 4.31 | 30.88 |
| Ziprasidone | 7/24 | 7.84E-04 | 0.0256 | 5.71 | 40.85 |
| tanespimycin ssMCF7 DOWN | 7/24 | 7.84E-04 | 0.0256 | 5.71 | 40.85 |
| toluene CTD 00006907 | 12/62 | 7.86E-04 | 0.0256 | 3.34 | 23.85 |
| Chromium(III) oxide CTD 00001091 | 12/62 | 7.86E-04 | 0.0256 | 3.34 | 23.85 |
| methylergometrine HL60 UP | 20/136 | 8.04E-04 | 0.0257 | 2.40 | 17.12 |
| Indeno[1,2,3-cd]pyrene CTD 00001895 | 8/31 | 8.21E-04 | 0.0257 | 4.83 | 34.30 |
| AC-42 TTD 00001459 | 8/31 | 8.21E-04 | 0.0257 | 4.83 | 34.30 |
| 17-Hydroxyandrostan-3-one CTD 00006776 | 25/187 | 8.23E-04 | 0.0257 | 2.15 | 15.29 |

| | | | | | |
|---|---|---|---|---|---|
| ZINC CTD 00007011 | 143/1642 | 8.27E-04 | 0.0257 | 1.36 | 9.62 |
| 4-Hydroxytamoxifen CTD 00000850 | 60/586 | 8.31E-04 | 0.0257 | 1.60 | 11.37 |
| clozapine BOSS | 13/71 | 8.38E-04 | 0.0257 | 3.12 | 22.08 |
| ETHYL METHANESULFONATE CTD 00005938 | 193/2315 | 9.31E-04 | 0.0282 | 1.30 | 9.07 |
| 3-methylcholanthrene CTD 00006310 | 35/297 | 9.37E-04 | 0.0282 | 1.87 | 13.02 |
| clozapine | 10/47 | 9.86E-04 | 0.0292 | 3.75 | 25.99 |
| phenoxybenzamine MCF7 DOWN | 10/47 | 9.86E-04 | 0.0292 | 3.75 | 25.99 |
| Premarin CTD 00005926 | 32/265 | 0.0010 | 0.0297 | 1.92 | 13.23 |
| anisomycin HL60 UP | 104/1142 | 0.0010 | 0.0297 | 1.42 | 9.75 |
| Sphingosine 1-phosphate CTD 00002508 | 7/25 | 0.0010 | 0.0297 | 5.40 | 37.13 |
| alexidine HL60 UP | 16/100 | 0.0010 | 0.0299 | 2.65 | 18.20 |
| doxycycline HL60 UP | 43/390 | 0.0010 | 0.0299 | 1.74 | 11.91 |
| scopolamine PC3 DOWN | 41/368 | 0.0011 | 0.0311 | 1.76 | 11.94 |
| 1H-Pyrrole-2,5-dione, 3,4-diphenyl- TTD 00000418 | 5/13 | 0.0011 | 0.0311 | 8.66 | 58.76 |
| SERTINDOLE TTD 00010935 | 5/13 | 0.0011 | 0.0311 | 8.66 | 58.76 |
| 1H-Pyrrole-2,5-dione, 3,4-bis(4-methoxyphenyl)- TTD 00000417 | 5/13 | 0.0011 | 0.0311 | 8.66 | 58.76 |
| AGN-PC-00BUNE TTD 00001660 | 5/13 | 0.0011 | 0.0311 | 8.66 | 58.76 |
| digoxin HL60 UP | 32/267 | 0.0011 | 0.0312 | 1.90 | 12.88 |
| Oxazolone CTD 00006449 | 19/130 | 0.0012 | 0.0313 | 2.38 | 16.12 |
| Hydroxychlor CTD 00003776 | 6/19 | 0.0012 | 0.0317 | 6.40 | 43.14 |
| metronidazole PC3 UP | 117/1316 | 0.0012 | 0.0319 | 1.38 | 9.28 |
| GONADORELIN BOSS | 25/192 | 0.0012 | 0.0319 | 2.09 | 14.03 |
| papaverine HL60 DOWN | 12/65 | 0.0012 | 0.0319 | 3.15 | 21.12 |
| doxazosin HL60 DOWN | 8/33 | 0.0013 | 0.0334 | 4.44 | 29.58 |
| Dasatinib CTD 00004330 | 52/502 | 0.0014 | 0.0354 | 1.62 | 10.66 |
| h-89 CTD 00002586 | 12/66 | 0.0014 | 0.0354 | 3.09 | 20.30 |
| NSC94017 CTD 00005320 | 14/84 | 0.0014 | 0.0354 | 2.78 | 18.27 |
| Lead(II) acetate CTD 00000394 | 14/84 | 0.0014 | 0.0354 | 2.78 | 18.27 |
| H-7 PC3 DOWN | 120/1361 | 0.0014 | 0.0354 | 1.37 | 8.99 |
| nocodazole HL60 UP | 41/373 | 0.0014 | 0.0359 | 1.73 | 11.31 |
| COUMESTROL CTD 00005717 | 154/1812 | 0.0015 | 0.0363 | 1.32 | 8.61 |
| NICKEL SULFATE CTD 00001417 | 58/576 | 0.0015 | 0.0366 | 1.57 | 10.23 |
| menadione PC3 UP | 17/113 | 0.0015 | 0.0366 | 2.46 | 16.04 |
| curcumin CTD 00000663 | 54/528 | 0.0015 | 0.0368 | 1.60 | 10.38 |
| geldanamycin MCF7 DOWN | 8/34 | 0.0016 | 0.0381 | 4.27 | 27.55 |
| F0447-0125 MCF7 DOWN | 6/20 | 0.0016 | 0.0383 | 5.94 | 38.29 |
| Sorafenib tosylate | 9/42 | 0.0017 | 0.0388 | 3.79 | 24.24 |
| clenbuterol PC3 UP | 9/42 | 0.0017 | 0.0388 | 3.79 | 24.24 |

| | | | | | |
|---|---|---|---|---|---|
| oxymetazoline CTD 00006455 | 5/14 | 0.0017 | 0.0388 | 7.70 | 49.27 |
| cinnarizine TTD 00007193 | 5/14 | 0.0017 | 0.0388 | 7.70 | 49.27 |
| piperlongumine MCF7 DOWN | 31/262 | 0.0017 | 0.0388 | 1.87 | 11.99 |
| doxorubicin MCF7 DOWN | 92/1006 | 0.0017 | 0.0401 | 1.42 | 9.02 |
| lycorine PC3 UP | 52/508 | 0.0018 | 0.0413 | 1.60 | 10.11 |
| vorinostat HL60 DOWN | 57/569 | 0.0018 | 0.0416 | 1.56 | 9.85 |
| H-7 MCF7 DOWN | 117/1332 | 0.0018 | 0.0416 | 1.36 | 8.58 |
| Andriol CTD 00000567 | 25/198 | 0.0019 | 0.0420 | 2.02 | 12.67 |
| etoposide CTD 00005948 | 48/461 | 0.0019 | 0.0420 | 1.63 | 10.22 |
| ciclopirox HL60 UP | 23/177 | 0.0019 | 0.0424 | 2.08 | 13.04 |
| Guanidine hydrochloride | 8/35 | 0.0019 | 0.0426 | 4.11 | 25.71 |
| CP-690334-01 MCF7 DOWN | 6/21 | 0.0021 | 0.0463 | 5.55 | 34.19 |
| GW-8510 PC3 DOWN | 115/1312 | 0.0021 | 0.0470 | 1.36 | 8.34 |
| clonidine HL60 UP | 13/79 | 0.0023 | 0.0502 | 2.74 | 16.62 |
| 4-aminopyridine | 8/36 | 0.0023 | 0.0502 | 3.96 | 24.03 |
| mepacrine HL60 UP | 9/44 | 0.0023 | 0.0502 | 3.57 | 21.63 |
| Doramapimod TTD 00007708 | 5/15 | 0.0024 | 0.0504 | 6.93 | 41.92 |
| clomipramine PC3 DOWN | 16/108 | 0.0024 | 0.0506 | 2.42 | 14.61 |
