## Supplementary Table S13 for "Genetic regulation of the vascular endothelial growth factor receptor 1 during sepsis and association with ARDS susceptibility"

**Table S13. Significant results of gene set enrichment analysis on the GWASCatalog dataset for the list of common genes obtained from the best PGS models at T1 and T7.**

| Term | Overlap | P-value | Adjusted P-value | Odds Ratio | Combined Score |
|---|---|---|---|---|---|
| Educational Attainment | 286/2058 | 1.03E-34 | 3.73E-31 | 2.56 | 200.37 |
| Metabolite Levels | 165/940 | 5.83E-31 | 1.06E-27 | 3.21 | 223.58 |
| Adolescent Idiopathic Scoliosis | 132/685 | 7.89E-29 | 9.55E-26 | 3.55 | 229.50 |
| Educational Attainment (MTAG) | 135/753 | 2.89E-26 | 2.62E-23 | 3.24 | 190.65 |
| Height | 593/6159 | 4.73E-26 | 3.44E-23 | 1.84 | 107.38 |
| Smoking Initiation | 177/1205 | 9.64E-24 | 5.83E-21 | 2.59 | 137.08 |
| Highest Math Class Taken (MTAG) | 113/644 | 2.61E-21 | 1.35E-18 | 3.12 | 147.73 |
| Weight | 103/587 | 1.68E-19 | 7.62E-17 | 3.10 | 134.02 |
| Body Mass Index | 231/1912 | 2.04E-19 | 8.25E-17 | 2.08 | 89.68 |
| Body Mass Index (MTAG) | 126/807 | 3.29E-19 | 1.20E-16 | 2.72 | 115.59 |
| Severe COVID-19 Infection | 81/415 | 2.27E-18 | 7.49E-16 | 3.50 | 142.21 |
| Educational Attainment (Years Of Education) | 122/801 | 9.43E-18 | 2.85E-15 | 2.63 | 103.08 |
| Vertex-wise Sulcal Depth | 97/576 | 3.48E-17 | 9.71E-15 | 2.94 | 111.30 |
| Adult Body Size | 73/368 | 4.50E-17 | 1.17E-14 | 3.56 | 133.88 |
| Obesity-related Traits | 85/498 | 1.51E-15 | 3.66E-13 | 2.97 | 101.26 |
| Cognitive Performance (MTAG) | 67/365 | 4.71E-14 | 1.07E-11 | 3.22 | 98.69 |
| Insomnia | 113/816 | 1.43E-13 | 3.06E-11 | 2.33 | 68.97 |
| Schizophrenia | 130/996 | 1.65E-13 | 3.32E-11 | 2.19 | 64.41 |
| Self-reported Math Ability (MTAG) | 74/440 | 2.34E-13 | 4.46E-11 | 2.90 | 84.26 |
| Refractive Error | 62/337 | 3.79E-13 | 6.57E-11 | 3.22 | 91.99 |
| Drinks Per Week | 72/426 | 3.86E-13 | 6.57E-11 | 2.91 | 83.22 |
| Protein Quantitative Trait Loci (Liver) | 130/1008 | 3.98E-13 | 6.57E-11 | 2.16 | 61.58 |
| DNA Methylation Variation (Age Effect) | 47/215 | 4.71E-13 | 7.44E-11 | 3.97 | 112.63 |
| Gut Microbiota (Bacterial Taxa, Hurdle Binary Method) | 38/168 | 2.74E-11 | 4.15E-09 | 4.13 | 100.35 |
| Age When Finished Full-Time Education (Standard GWA) | 34/142 | 5.81E-11 | 8.43E-09 | 4.44 | 104.55 |
| Cortical Thickness | 64/401 | 9.95E-11 | 1.39E-08 | 2.70 | 62.28 |
| Carotid Intima-media Thickness (Mean Of The Maximum cIMT) | 31/124 | 1.26E-10 | 1.69E-08 | 4.69 | 106.90 |
| Cortical Surface Area | 74/506 | 2.26E-10 | 2.93E-08 | 2.45 | 54.32 |
| 3-Hydroxy-1-Methylpropylmercapturic Acid Levels In Smokers | 31/127 | 2.41E-10 | 3.02E-08 | 4.54 | 100.60 |
| Chronotype | 46/257 | 1.06E-09 | 1.28E-07 | 3.08 | 63.71 |
| Externalizing Behaviour (Multivariate Analysis) | 55/339 | 1.13E-09 | 1.32E-07 | 2.75 | 56.59 |
| Depression | 38/190 | 1.21E-09 | 1.37E-07 | 3.52 | 72.37 |
| General Cognitive Ability | 83/620 | 1.48E-09 | 1.63E-07 | 2.21 | 44.92 |
| Type 2 Diabetes | 110/913 | 1.65E-09 | 1.76E-07 | 1.97 | 39.87 |
| Trauma Exposure | 16/40 | 2.27E-09 | 2.36E-07 | 9.31 | 185.26 |
| Resistance To COVID-19 Infection (Exposed Negative Vs Positive) | 58/375 | 2.53E-09 | 2.56E-07 | 2.60 | 51.39 |

| | | | | | |
|---|---|---|---|---|---|
| Vertex-wise Cortical Surface Area | 63/427 | 3.53E-09 | 3.47E-07 | 2.46 | 47.86 |
| Smoking Initiation (Ever Regular Vs Never Regular) | 44/249 | 3.66E-09 | 3.50E-07 | 3.03 | 58.89 |
| Body Surface Area | 51/317 | 6.05E-09 | 5.63E-07 | 2.71 | 51.35 |
| Smoking Initiation (Ever Regular Vs Never Regular) (MTAG) | 59/401 | 1.23E-08 | 1.11E-06 | 2.45 | 44.56 |
| Morningness | 33/164 | 1.26E-08 | 1.11E-06 | 3.54 | 64.44 |
| Major Depressive Disorder | 40/226 | 1.78E-08 | 1.50E-06 | 3.03 | 54.09 |
| Vertex-wise Cortical Thickness | 59/405 | 1.78E-08 | 1.50E-06 | 2.42 | 43.15 |
| Neuroticism | 51/332 | 2.90E-08 | 2.40E-06 | 2.57 | 44.54 |
| IgG Glycosylation | 38/213 | 3.22E-08 | 2.60E-06 | 3.06 | 52.75 |
| PR Interval | 34/180 | 3.98E-08 | 3.14E-06 | 3.27 | 55.79 |
| Diisocyanate-induced Asthma | 24/101 | 4.41E-08 | 3.41E-06 | 4.37 | 73.94 |
| Spherical Equivalent | 29/142 | 6.73E-08 | 5.09E-06 | 3.60 | 59.47 |
| Interferon-Related Traits | 30/151 | 7.69E-08 | 5.70E-06 | 3.48 | 57.01 |
| Blood Pressure (Pleiotropy Model 2 SBP Adjusted For Estimated Causal Effects X DBP) | 45/285 | 8.27E-08 | 6.01E-06 | 2.65 | 43.14 |
| Self-reported Math Ability | 49/326 | 1.08E-07 | 7.64E-06 | 2.50 | 40.08 |
| QRS Duration | 26/121 | 1.09E-07 | 7.64E-06 | 3.84 | 61.48 |
| Heel Bone Mineral Density | 114/1034 | 1.16E-07 | 7.94E-06 | 1.78 | 28.39 |
| Cigarette Consumption X Hours Spent Using Computers Interaction | 13/34 | 1.39E-07 | 9.32E-06 | 8.63 | 136.21 |
| 3-Hydroxypropylmercapturic Acid Levels In Smokers | 26/124 | 1.83E-07 | 1.21E-05 | 3.72 | 57.67 |
| Reaction Time | 42/265 | 1.99E-07 | 1.29E-05 | 2.65 | 40.93 |
| Peripheral Arterial Disease (Traffic-Related Air Pollution Interaction) | 21/88 | 2.81E-07 | 1.79E-05 | 4.38 | 66.11 |
| COVID-19 | 26/127 | 3.01E-07 | 1.89E-05 | 3.61 | 54.15 |
| Age At First Sexual Intercourse | 30/161 | 3.35E-07 | 2.06E-05 | 3.21 | 47.90 |
| RBC Levels Of Pyruvate (uM) | 17/61 | 3.54E-07 | 2.14E-05 | 5.39 | 80.11 |
| Brain Morphology (MOSTest) | 61/463 | 3.96E-07 | 2.36E-05 | 2.15 | 31.67 |
| Biological Sex | 22/98 | 4.61E-07 | 2.65E-05 | 4.05 | 59.07 |
| R-warfarin Levels | 22/98 | 4.61E-07 | 2.65E-05 | 4.05 | 59.07 |
| Body Size At Age 10 | 33/191 | 5.46E-07 | 3.10E-05 | 2.93 | 42.29 |
| Hair Curvature (Quantitative) | 10/22 | 5.80E-07 | 3.23E-05 | 11.59 | 166.45 |
| Tuberculosis | 17/63 | 5.87E-07 | 3.23E-05 | 5.16 | 74.01 |
| Blood Pressure (Pleiotropy Model 1 DBP Adjusted For Estimated Causal Effects X SBP) | 45/305 | 6.02E-07 | 3.26E-05 | 2.44 | 34.94 |
| Hair Color | 42/277 | 6.79E-07 | 3.62E-05 | 2.52 | 35.74 |
| Chronic Obstructive Pulmonary Disease Liability (Machine Learning-Based Score) | 39/249 | 7.33E-07 | 3.85E-05 | 2.61 | 36.90 |
| Hemoglobin | 63/493 | 7.50E-07 | 3.89E-05 | 2.07 | 29.25 |
| BMI (Standard GWA) | 28/151 | 9.06E-07 | 4.63E-05 | 3.19 | 44.39 |
| Night Sleep Phenotypes | 21/95 | 1.09E-06 | 5.52E-05 | 3.97 | 54.44 |
| Bipolar Disorder And Schizophrenia | 18/73 | 1.18E-06 | 5.85E-05 | 4.57 | 62.38 |

| | | | | | |
|---|---|---|---|---|---|
| Appendicular Lean Mass | 93/836 | 1.21E-06 | 5.94E-05 | 1.78 | 24.29 |
| Hippocampal Volume | 13/40 | 1.23E-06 | 5.94E-05 | 6.71 | 91.28 |
| Depressive Symptoms | 30/171 | 1.27E-06 | 6.06E-05 | 2.98 | 40.51 |
| Response To Antipsychotic Treatment | 11/29 | 1.42E-06 | 6.68E-05 | 8.50 | 114.52 |
| COVID-19 (Hospitalized Vs Tested, Not Hospitalized) | 16/61 | 1.90E-06 | 8.82E-05 | 4.96 | 65.34 |
| Venous Thromboembolism Adjusted For Sickle Cell Variant rs77121243-T | 14/48 | 2.09E-06 | 9.59E-05 | 5.74 | 75.05 |
| Lung Function (FEV1/FVC) | 70/589 | 2.68E-06 | 1.22E-04 | 1.91 | 24.51 |
| Adverse Response To Drug | 15/56 | 2.97E-06 | 1.33E-04 | 5.10 | 64.90 |
| Response To Zileuton Treatment In Asthma (FEV1 Change Interaction) | 8/16 | 3.33E-06 | 1.47E-04 | 13.89 | 175.20 |
| Waist-hip Ratio | 73/628 | 3.68E-06 | 1.61E-04 | 1.86 | 23.32 |
| Systolic Blood Pressure | 111/1073 | 3.80E-06 | 1.63E-04 | 1.65 | 20.56 |
| Morning Person | 29/171 | 3.83E-06 | 1.63E-04 | 2.86 | 35.69 |
| Lung Function (Forced Vital Capacity) | 29/172 | 4.31E-06 | 1.82E-04 | 2.84 | 35.10 |
| Lung Function (FVC) | 58/466 | 4.70E-06 | 1.96E-04 | 2.01 | 24.62 |
| Highest Math Class Taken | 33/211 | 5.26E-06 | 2.17E-04 | 2.60 | 31.61 |
| Colonoscopy-negative Controls Vs Population Controls | 17/73 | 5.38E-06 | 2.19E-04 | 4.23 | 51.38 |
| Breast Cancer | 58/469 | 5.74E-06 | 2.31E-04 | 1.99 | 24.04 |
| Atrial Fibrillation (MTAG) | 19/89 | 6.06E-06 | 2.42E-04 | 3.79 | 45.52 |
| Progression Free Survival In Serous Epithelial Ovarian Cancer Treated With Carboplatin And Paclitaxel | 9/22 | 6.30E-06 | 2.49E-04 | 9.62 | 115.22 |
| Waist Circumference | 27/158 | 7.05E-06 | 2.73E-04 | 2.89 | 34.22 |
| Depressed Affect | 16/67 | 7.15E-06 | 2.73E-04 | 4.37 | 51.83 |
| PR Interval In Tripanosoma Cruzi Seropositivity | 16/67 | 7.15E-06 | 2.73E-04 | 4.37 | 51.83 |
| Diastolic Blood Pressure | 87/803 | 7.65E-06 | 2.89E-04 | 1.73 | 20.33 |
| Cerebellum Cortex Volume Change Rate | 5/6 | 7.88E-06 | 2.92E-04 | 69.33 | 814.67 |
| Ceramide (D17:1/16:0) Levels | 5/6 | 7.88E-06 | 2.92E-04 | 69.33 | 814.67 |
| Cognitive Ability, Years Of Educational Attainment Or Schizophrenia (Pleiotropy) | 27/159 | 7.95E-06 | 2.92E-04 | 2.86 | 33.62 |
| Aspartate Aminotransferase Levels In Low Alcohol Consumption | 10/28 | 8.10E-06 | 2.94E-04 | 7.72 | 90.57 |
| Post Bronchodilator FEV1 In COPD | 13/47 | 9.20E-06 | 2.98E-04 | 5.32 | 61.73 |
| Myopia | 17/76 | 9.59E-06 | 2.98E-04 | 4.02 | 46.44 |
| Stimulated Adipocyte Lipolysis | 9/23 | 9.73E-06 | 2.98E-04 | 8.93 | 103.10 |
| Leisure Sedentary Behaviour (Television Watching) | 20/100 | 9.88E-06 | 2.98E-04 | 3.49 | 40.23 |
| Triglyceride Levels | 84/774 | 1.03E-05 | 2.98E-04 | 1.73 | 19.83 |
| Post Bronchodilator FEV1/FVC Ratio | 37/258 | 1.13E-05 | 2.98E-04 | 2.35 | 26.76 |
| Cognitive Function (Generalized Correlation Coefficient) | 11/35 | 1.17E-05 | 2.98E-04 | 6.38 | 72.39 |
| Nickel Levels | 11/35 | 1.17E-05 | 2.98E-04 | 6.38 | 72.39 |

| | | | | | |
|---|---|---|---|---|---|
| Cobalt Levels | 11/35 | 1.17E-05 | 2.98E-04 | 6.38 | 72.39 |
| Mercuric Chloride-2 Levels | 11/35 | 1.17E-05 | 2.98E-04 | 6.38 | 72.39 |
| Methoxychlor Levels | 11/35 | 1.17E-05 | 2.98E-04 | 6.38 | 72.39 |
| Environmental Pollutants Exposure (POD Low) | 11/35 | 1.17E-05 | 2.98E-04 | 6.38 | 72.39 |
| Potassium Chromate Levels | 11/35 | 1.17E-05 | 2.98E-04 | 6.38 | 72.39 |
| Environmental Pollutants Exposure (RfD High) | 11/35 | 1.17E-05 | 2.98E-04 | 6.38 | 72.39 |
| Parathion Levels | 11/35 | 1.17E-05 | 2.98E-04 | 6.38 | 72.39 |
| Environmental Pollutants Exposure (RfD Low) | 11/35 | 1.17E-05 | 2.98E-04 | 6.38 | 72.39 |
| Heptachlor Levels | 11/35 | 1.17E-05 | 2.98E-04 | 6.38 | 72.39 |
| Heptachlor Epoxide Levels | 11/35 | 1.17E-05 | 2.98E-04 | 6.38 | 72.39 |
| Mercuric Chloride Levels | 11/35 | 1.17E-05 | 2.98E-04 | 6.38 | 72.39 |
| Pentachlorophenol Levels | 11/35 | 1.17E-05 | 2.98E-04 | 6.38 | 72.39 |
| Environmental Pollutants Exposure (POD High) | 11/35 | 1.17E-05 | 2.98E-04 | 6.38 | 72.39 |
| 2,4,5-Trichlorophenol Levels | 11/35 | 1.17E-05 | 2.98E-04 | 6.38 | 72.39 |
| Environmental Pollutants Exposure (AC50 Low) | 11/35 | 1.17E-05 | 2.98E-04 | 6.38 | 72.39 |
| Aldrin Levels | 11/35 | 1.17E-05 | 2.98E-04 | 6.38 | 72.39 |
| Azinphos Methyl Levels | 11/35 | 1.17E-05 | 2.98E-04 | 6.38 | 72.39 |
| Cadmium Chloride Levels | 11/35 | 1.17E-05 | 2.98E-04 | 6.38 | 72.39 |
| Chlorpyrifos Levels | 11/35 | 1.17E-05 | 2.98E-04 | 6.38 | 72.39 |
| DDT Metabolite (DDD P-P') Levels | 11/35 | 1.17E-05 | 2.98E-04 | 6.38 | 72.39 |
| DDT Metabolite (DDT O-P') Levels | 11/35 | 1.17E-05 | 2.98E-04 | 6.38 | 72.39 |
| Environmental Pollutants Exposure (AC50 High) | 11/35 | 1.17E-05 | 2.98E-04 | 6.38 | 72.39 |
| DDT Metabolite (DDT P-P') Levels | 11/35 | 1.17E-05 | 2.98E-04 | 6.38 | 72.39 |
| Disulfoton Levels | 11/35 | 1.17E-05 | 2.98E-04 | 6.38 | 72.39 |
| Endosulfan Levels | 11/35 | 1.17E-05 | 2.98E-04 | 6.38 | 72.39 |
| Endrin Levels | 11/35 | 1.17E-05 | 2.98E-04 | 6.38 | 72.39 |
| 2,4,5-Trichlorophenol-2 Levels | 11/35 | 1.17E-05 | 2.98E-04 | 6.38 | 72.39 |
| 4,6-Dinitro-O-Cresol Levels | 11/35 | 1.17E-05 | 2.98E-04 | 6.38 | 72.39 |
| Ethion Levels | 11/35 | 1.17E-05 | 2.98E-04 | 6.38 | 72.39 |
| Di-n-butyl Phthalate Levels | 11/35 | 1.17E-05 | 2.98E-04 | 6.38 | 72.39 |
| Diazinon Levels | 11/35 | 1.17E-05 | 2.98E-04 | 6.38 | 72.39 |
| Dicofol Levels | 11/35 | 1.17E-05 | 2.98E-04 | 6.38 | 72.39 |
| Dieldrin Levels | 11/35 | 1.17E-05 | 2.98E-04 | 6.38 | 72.39 |
| Environmental Pollutants Exposure (Expo High) | 11/35 | 1.17E-05 | 2.98E-04 | 6.38 | 72.39 |
| Environmental Pollutants Exposure (Expo Low) | 11/35 | 1.17E-05 | 2.98E-04 | 6.38 | 72.39 |
| Attention Deficit Hyperactivity Disorder | 18/86 | 1.41E-05 | 3.52E-04 | 3.69 | 41.24 |
| Gut Microbiota Alpha Diversity (PD Whole Tree Index) | 7/14 | 1.41E-05 | 3.52E-04 | 13.88 | 155.00 |
| Metabolite Levels (Pyroglutamine) | 7/14 | 1.41E-05 | 3.52E-04 | 13.88 | 155.00 |

| | | | | | |
|---|---|---|---|---|---|
| Tonometry | 6/10 | 1.55E-05 | 3.83E-04 | 20.81 | 230.45 |
| Gut Microbiota (Beta Diversity) | 11/36 | 1.59E-05 | 3.89E-04 | 6.12 | 67.64 |
| Immune Response To Smallpox Vaccine (IL-6) | 10/30 | 1.64E-05 | 3.96E-04 | 6.95 | 76.61 |
| Alzheimer's Disease | 26/156 | 1.64E-05 | 3.96E-04 | 2.80 | 30.83 |
| Depression (Broad) | 13/50 | 1.92E-05 | 4.62E-04 | 4.89 | 53.12 |
| Internet Addiction Disorder | 9/25 | 2.15E-05 | 5.14E-04 | 7.82 | 84.00 |
| Well-being Spectrum (Multivariate Analysis) | 23/132 | 2.38E-05 | 5.66E-04 | 2.95 | 31.38 |
| F-sharp Flavour Liking (Derived Food-Liking Factor) | 7/15 | 2.50E-05 | 5.89E-04 | 12.15 | 128.71 |
| Cytomegalovirus Antibody Response | 5/7 | 2.60E-05 | 6.10E-04 | 34.66 | 365.88 |
| Daytime Nap | 16/74 | 2.73E-05 | 6.32E-04 | 3.84 | 40.40 |
| COVID-19 (Hospitalized Vs Not Hospitalized) | 16/74 | 2.73E-05 | 6.32E-04 | 3.84 | 40.40 |
| Number Of Sexual Partners | 14/59 | 2.86E-05 | 6.56E-04 | 4.33 | 45.33 |
| General Risk Tolerance (MTAG) | 26/161 | 2.89E-05 | 6.59E-04 | 2.69 | 28.15 |
| COVID-19 (Hospitalized Vs Population) | 18/91 | 3.17E-05 | 7.14E-04 | 3.44 | 35.63 |
| Retinal Vascular Fractal Density | 18/91 | 3.17E-05 | 7.14E-04 | 3.44 | 35.63 |
| Gut Microbiota Alpha Diversity (Simpson Index) | 6/11 | 3.22E-05 | 7.21E-04 | 16.65 | 172.21 |
| Bipolar Disorder Or Major Depressive Disorder | 15/68 | 3.77E-05 | 8.40E-04 | 3.94 | 40.16 |
| Myopia (Age Of Diagnosis) | 16/76 | 3.87E-05 | 8.57E-04 | 3.72 | 37.75 |
| Refractive Astigmatism | 10/33 | 4.18E-05 | 9.21E-04 | 6.04 | 60.93 |
| Facial Wrinkles | 9/27 | 4.36E-05 | 9.54E-04 | 6.95 | 69.75 |
| S-6-hydroxywarfarin Levels | 22/128 | 4.41E-05 | 9.60E-04 | 2.90 | 29.06 |
| Rate Of Cognitive Decline In Alzheimer's Disease | 16/77 | 4.58E-05 | 9.89E-04 | 3.66 | 36.51 |
| Height (Standard GWA) | 48/392 | 4.60E-05 | 9.89E-04 | 1.96 | 19.59 |
| Menarche (Age At Onset) | 37/276 | 5.07E-05 | 0.0011 | 2.17 | 21.47 |
| Airflow Obstruction | 8/22 | 5.74E-05 | 0.0012 | 7.94 | 77.50 |
| Neutral Antibody Levels In Response To SARS-CoV-2 Vaccination | 6/12 | 6.06E-05 | 0.0013 | 13.87 | 134.70 |
| Anxiety And Stress-Related Disorders | 5/8 | 6.56E-05 | 0.0014 | 23.11 | 222.57 |
| N-methylpipecolate Levels In Elite Athletes | 5/8 | 6.56E-05 | 0.0014 | 23.11 | 222.57 |
| Longevity | 18/96 | 6.65E-05 | 0.0014 | 3.22 | 30.95 |
| Moderate-to-late Spontaneous Preterm Birth | 7/17 | 6.70E-05 | 0.0014 | 9.72 | 93.38 |
| Brain Morphology (min-P) | 16/80 | 7.46E-05 | 0.0015 | 3.48 | 33.10 |
| High-sensitivity Cardiac Troponin I Concentration | 14/64 | 7.48E-05 | 0.0015 | 3.90 | 37.03 |
| Predicted Developmental Stuttering | 12/49 | 7.57E-05 | 0.0015 | 4.51 | 42.81 |
| Adventurousness | 18/97 | 7.66E-05 | 0.0015 | 3.18 | 30.11 |
| Cognitive Performance | 29/200 | 7.93E-05 | 0.0016 | 2.37 | 22.40 |
| Lung Function (FEV1) | 30/210 | 7.95E-05 | 0.0016 | 2.33 | 22.01 |
| Strep Throat | 9/29 | 8.24E-05 | 0.0016 | 6.25 | 58.79 |

| | | | | | |
|---|---|---|---|---|---|
| COVID-19 (Severe Respiratory Symptoms Vs Population) | 9/29 | 8.24E-05 | 0.0016 | 6.25 | 58.79 |
| Intraocular Pressure | 33/241 | 8.36E-05 | 0.0016 | 2.22 | 20.86 |
| Low Myopia Vs Hyperopia | 17/90 | 9.55E-05 | 0.0019 | 3.25 | 30.04 |
| Crohn's Disease (Indolent Vs Progressive) | 4/5 | 9.78E-05 | 0.0019 | 55.42 | 511.68 |
| Middle Childhood And Early Adolescence Aggressive Behavior | 4/5 | 9.78E-05 | 0.0019 | 55.42 | 511.68 |
| Taurolithocholate 3-Sulfate Levels In Elite Athletes | 4/5 | 9.78E-05 | 0.0019 | 55.42 | 511.68 |
| Metabolite Peak Levels (QI9680) | 4/5 | 9.78E-05 | 0.0019 | 55.42 | 511.68 |
| Left Ventricle Wall Thickness | 7/18 | 1.03E-04 | 0.0019 | 8.83 | 81.07 |
| Progression Free Survival In Epithelial Ovarian Cancer Treated With Carboplatin And Paclitaxel | 7/18 | 1.03E-04 | 0.0019 | 8.83 | 81.07 |
| Vaginal Microbiome Relative Abundance (S Dialister Propionicifaciens) | 7/18 | 1.03E-04 | 0.0019 | 8.83 | 81.07 |
| Resting-state Electroencephalogram Vigilance | 7/18 | 1.03E-04 | 0.0019 | 8.83 | 81.07 |
| Stromal-cell-derived Factor 1 Alpha Levels | 6/13 | 1.06E-04 | 0.0020 | 11.89 | 108.79 |
| Dialysis-related Mortality | 6/13 | 1.06E-04 | 0.0020 | 11.89 | 108.79 |
| Intracranial Aneurysm | 14/66 | 1.07E-04 | 0.0020 | 3.75 | 34.28 |
| Anorexia Nervosa | 9/30 | 1.11E-04 | 0.0020 | 5.95 | 54.23 |
| S-7-hydroxywarfarin To S-warfarin Ratio | 9/30 | 1.11E-04 | 0.0020 | 5.95 | 54.23 |
| Multisite Chronic Pain | 12/51 | 1.15E-04 | 0.0021 | 4.28 | 38.82 |
| Red Blood Cell Count | 98/1001 | 1.20E-04 | 0.0022 | 1.54 | 13.89 |
| Waist Circumference Adjusted For Body Mass Index | 95/966 | 1.30E-04 | 0.0023 | 1.55 | 13.83 |
| Pulse Pressure | 76/735 | 1.32E-04 | 0.0024 | 1.63 | 14.55 |
| Risk-taking Behavior (Multivariate Analysis) | 23/147 | 1.33E-04 | 0.0024 | 2.59 | 23.10 |
| Lobe Attachment (Rater-Scored Or Self-Reported) | 16/84 | 1.37E-04 | 0.0024 | 3.28 | 29.16 |
| Hematocrit | 61/559 | 1.42E-04 | 0.0025 | 1.72 | 15.29 |
| Spherical Equivalent Or Myopia (Age Of Diagnosis) | 17/93 | 1.45E-04 | 0.0026 | 3.12 | 27.54 |
| Age When Finished Full-Time Education (Weighted GWA) | 9/31 | 1.47E-04 | 0.0026 | 5.68 | 50.16 |
| Intelligence (MTAG) | 32/238 | 1.50E-04 | 0.0026 | 2.17 | 19.14 |
| Corneal Astigmatism | 11/45 | 1.53E-04 | 0.0026 | 4.50 | 39.52 |
| Major Depressive Disorder (MTAG) | 11/45 | 1.53E-04 | 0.0026 | 4.50 | 39.52 |
| Type 2 Diabetes (Age Of Onset) | 7/19 | 1.54E-04 | 0.0026 | 8.10 | 71.07 |
| Response To Esketamine In Treatment Resistant Depression | 8/25 | 1.62E-04 | 0.0028 | 6.53 | 57.04 |
| Hip Circumference | 21/130 | 1.63E-04 | 0.0028 | 2.69 | 23.45 |

| | | | | | |
|---|---|---|---|---|---|
| Medication Use For Hypertension (Number Of Purchases) | 20/121 | 1.68E-04 | 0.0028 | 2.76 | 24.01 |
| Atopic Dermatitis (Moderate To Severe) | 6/14 | 1.75E-04 | 0.0029 | 10.40 | 89.98 |
| Haemorrhoidal Disease | 14/69 | 1.76E-04 | 0.0029 | 3.54 | 30.62 |
| FEV1 | 22/141 | 1.93E-04 | 0.0032 | 2.58 | 22.07 |
| Balding Type 1 | 26/180 | 1.93E-04 | 0.0032 | 2.36 | 20.17 |
| Whole Brain Restricted Directional Diffusion (Multivariate Analysis) | 26/180 | 1.93E-04 | 0.0032 | 2.36 | 20.17 |
| Subjective Well-Being | 15/78 | 1.97E-04 | 0.0032 | 3.31 | 28.29 |
| R-6-hydroxywarfarin To R-warfarin Ratio | 10/39 | 1.98E-04 | 0.0032 | 4.79 | 40.86 |
| Male-pattern Baldness | 40/328 | 2.08E-04 | 0.0034 | 1.95 | 16.50 |
| Epithelial Ovarian Cancer | 18/105 | 2.19E-04 | 0.0035 | 2.88 | 24.30 |
| Added Salt Consumption | 8/26 | 2.20E-04 | 0.0035 | 6.17 | 51.97 |
| Gestational Age At Birth (Child Effect) | 7/20 | 2.23E-04 | 0.0036 | 7.47 | 62.83 |
| F-strong Flavour Liking (Derived Food-Liking Factor) | 7/20 | 2.23E-04 | 0.0036 | 7.47 | 62.83 |
| Response To Amphetamines | 10/40 | 2.48E-04 | 0.0040 | 4.63 | 38.45 |
| Angioedema In Response To Angiotensin-Converting Enzyme Inhibitor And/Or Angiotensin Receptor Blocker | 5/10 | 2.63E-04 | 0.0041 | 13.86 | 114.27 |
| Western Dietary Pattern | 5/10 | 2.63E-04 | 0.0041 | 13.86 | 114.27 |
| LDL Levels X SSRI Levels (Escitalopram Or Citalopram) Interaction In Schizophrenia Or Bipolar Disorder | 5/10 | 2.63E-04 | 0.0041 | 13.86 | 114.27 |
| Complement Factor H-like 1 Protein Levels | 5/10 | 2.63E-04 | 0.0041 | 13.86 | 114.27 |
| Pyridoxate Levels In Elite Athletes | 5/10 | 2.63E-04 | 0.0041 | 13.86 | 114.27 |
| Glycosphingolipids (D42:2) Levels | 5/10 | 2.63E-04 | 0.0041 | 13.86 | 114.27 |
| Colorectal Cancer | 34/267 | 2.71E-04 | 0.0042 | 2.04 | 16.77 |
| Cardio-cerebrovascular Disease In Diabetes Mellitus | 6/15 | 2.75E-04 | 0.0042 | 9.25 | 75.80 |
| Verbal Declarative Memory | 6/15 | 2.75E-04 | 0.0042 | 9.25 | 75.80 |
| Triiodothyronine Levels | 4/6 | 2.78E-04 | 0.0042 | 27.71 | 226.91 |
| Osteosarcoma | 4/6 | 2.78E-04 | 0.0042 | 27.71 | 226.91 |
| Hip Shape (DXA Scan) | 4/6 | 2.78E-04 | 0.0042 | 27.71 | 226.91 |
| Radiation-Induced Toxicity (Physician-Rated Acute Dysphagia) | 4/6 | 2.78E-04 | 0.0042 | 27.71 | 226.91 |
| Champagne Or White Wine Consumption (Glasses Per Month) (UKB Data Field 1578, 4418) | 4/6 | 2.78E-04 | 0.0042 | 27.71 | 226.91 |
| Ebbinghaus Illusion (Overestimation) | 11/48 | 2.82E-04 | 0.0042 | 4.13 | 33.78 |
| Obesity | 10/41 | 3.09E-04 | 0.0046 | 4.48 | 36.23 |
| Electrocardiogram Morphology (Amplitude At Temporal Datapoints) | 34/269 | 3.12E-04 | 0.0046 | 2.02 | 16.34 |
| Blond Vs. Brown/Black Hair Color | 18/108 | 3.14E-04 | 0.0046 | 2.79 | 22.48 |

| | | | | | |
|---|---|---|---|---|---|
| Ischemic Stroke In Diabetes Mellitus | 7/21 | 3.15E-04 | 0.0046 | 6.94 | 55.94 |
| F-acquired Taste Liking (Derived Food-Liking Factor) | 7/21 | 3.15E-04 | 0.0046 | 6.94 | 55.94 |
| Body Fat Percentage | 30/227 | 3.21E-04 | 0.0047 | 2.13 | 17.12 |
| Atrial Fibrillation | 26/186 | 3.27E-04 | 0.0048 | 2.27 | 18.21 |
| Bipolar Disorder | 29/217 | 3.32E-04 | 0.0048 | 2.16 | 17.27 |
| Stroke | 11/49 | 3.42E-04 | 0.0049 | 4.02 | 32.12 |
| Tinnitus | 11/49 | 3.42E-04 | 0.0049 | 4.02 | 32.12 |
| COVID-19 (Hospitalized Covid Vs Population) | 12/57 | 3.52E-04 | 0.0050 | 3.71 | 29.49 |
| Peak Expiratory Flow | 23/157 | 3.59E-04 | 0.0051 | 2.39 | 19.00 |
| Cognitive Aspects Of Educational Attainment | 23/157 | 3.59E-04 | 0.0051 | 2.39 | 19.00 |
| Smoking Status (Ever Vs Never Smokers) | 20/128 | 3.63E-04 | 0.0051 | 2.58 | 20.45 |
| Neuroblastoma | 11/50 | 4.12E-04 | 0.0058 | 3.92 | 30.56 |
| Ghrelin Levels | 6/16 | 4.16E-04 | 0.0058 | 8.32 | 64.79 |
| Left Unilateral Cleft Lip | 6/16 | 4.16E-04 | 0.0058 | 8.32 | 64.79 |
| Vertigo | 12/58 | 4.17E-04 | 0.0058 | 3.63 | 28.23 |
| Predicted Visceral Adipose Tissue | 20/130 | 4.46E-04 | 0.0062 | 2.53 | 19.55 |
| Interstitial Lung Diseases In Rheumatoid Arthritis | 5/11 | 4.55E-04 | 0.0063 | 11.55 | 88.88 |
| Glycosphingolipids (D40:1) Levels | 5/11 | 4.55E-04 | 0.0063 | 11.55 | 88.88 |
| Gastroesophageal Reflux Disease | 10/43 | 4.67E-04 | 0.0064 | 4.21 | 32.29 |
| Risk-taking Tendency (4-Domain Principal Component Model) | 13/67 | 4.70E-04 | 0.0064 | 3.35 | 25.66 |
| Subcortical Volume (min-P) | 13/67 | 4.70E-04 | 0.0064 | 3.35 | 25.66 |
| Alcohol Consumption (Drinks Per Week) (MTAG) | 19/121 | 4.72E-04 | 0.0064 | 2.60 | 19.88 |
| R-6-hydroxywarfarin Levels | 11/51 | 4.93E-04 | 0.0067 | 3.82 | 29.10 |
| Psychosis In Alzheimer's Disease | 8/29 | 5.04E-04 | 0.0068 | 5.29 | 40.16 |
| Thiazide-induced Adverse Metabolic Effects In Hypertensive Patients | 9/36 | 5.05E-04 | 0.0068 | 4.63 | 35.14 |
| Whole Brain Restricted Isotropic Diffusion (Multivariate Analysis) | 27/202 | 5.22E-04 | 0.0070 | 2.15 | 16.28 |
| Body Fat Distribution (Leg Fat Ratio) | 13/68 | 5.46E-04 | 0.0073 | 3.29 | 24.70 |
| Neuritic Plaque | 7/23 | 5.90E-04 | 0.0078 | 6.07 | 45.14 |
| Rubella | 7/23 | 5.90E-04 | 0.0078 | 6.07 | 45.14 |
| Myringotomy | 7/23 | 5.90E-04 | 0.0078 | 6.07 | 45.14 |
| HDL Cholesterol Levels | 55/519 | 6.03E-04 | 0.0078 | 1.66 | 12.34 |
| Differentiated Thyroid Cancer | 6/17 | 6.06E-04 | 0.0078 | 7.56 | 56.05 |
| Academic Attainment (Maths) | 6/17 | 6.06E-04 | 0.0078 | 7.56 | 56.05 |
| Rheumatic Fever | 6/17 | 6.06E-04 | 0.0078 | 7.56 | 56.05 |
| Lung Function | 4/7 | 6.13E-04 | 0.0078 | 18.47 | 136.63 |
| Normal Facial Asymmetry (Angle Of Surface Orientation Score) | 4/7 | 6.13E-04 | 0.0078 | 18.47 | 136.63 |
| Adverse Response To Lamotrigine And Phenytoin | 4/7 | 6.13E-04 | 0.0078 | 18.47 | 136.63 |

| | | | | | |
|---|---|---|---|---|---|
| Gestational Age At Birth In Premature Rupture Of Membrane-Initiated Deliveries (Maternal Effect) | 4/7 | 6.13E-04 | 0.0078 | 18.47 | 136.63 |
| Age Related Hearing Loss-Related Regional Glucose Metabolism (Cochlear Nucleus) | 4/7 | 6.13E-04 | 0.0078 | 18.47 | 136.63 |
| Facial Skin Gloss | 4/7 | 6.13E-04 | 0.0078 | 18.47 | 136.63 |
| Alkylphosphatidylcholine (O-18:0/18:1) Levels | 4/7 | 6.13E-04 | 0.0078 | 18.47 | 136.63 |
| Cognitive Decline Rate In Late Mild Cognitive Impairment | 9/37 | 6.27E-04 | 0.0079 | 4.46 | 32.91 |
| Myopia (Pathological) | 9/37 | 6.27E-04 | 0.0079 | 4.46 | 32.91 |
| Fat-free Mass | 19/124 | 6.45E-04 | 0.0080 | 2.52 | 18.52 |
| 3-Month Functional Outcome In Ischaemic Stroke (Modified Rankin Score) | 8/30 | 6.47E-04 | 0.0080 | 5.05 | 37.07 |
| Nicotine Dependence | 8/30 | 6.47E-04 | 0.0080 | 5.05 | 37.07 |
| Ischemic Stroke (Cardioembolic) | 8/30 | 6.47E-04 | 0.0080 | 5.05 | 37.07 |
| Waist-to-hip Ratio Adjusted For BMI | 86/900 | 6.67E-04 | 0.0082 | 1.49 | 10.90 |
| Life Satisfaction | 15/87 | 6.70E-04 | 0.0082 | 2.90 | 21.19 |
| Amyotrophic Lateral Sclerosis | 11/53 | 6.97E-04 | 0.0086 | 3.64 | 26.45 |
| Intelligence | 38/326 | 7.16E-04 | 0.0088 | 1.85 | 13.37 |
| Disc Degeneration (Lumbar) | 5/12 | 7.38E-04 | 0.0089 | 9.90 | 71.41 |
| Fractional Exhaled Nitric Oxide (Childhood) | 5/12 | 7.38E-04 | 0.0089 | 9.90 | 71.41 |
| Hoarding Symptoms | 5/12 | 7.38E-04 | 0.0089 | 9.90 | 71.41 |
| Colorectal Cancer X Estrogen-Progesterone Hormone Therapy Interaction | 5/12 | 7.38E-04 | 0.0089 | 9.90 | 71.41 |
| Brain Shape (Segment 1) | 15/88 | 7.58E-04 | 0.0091 | 2.86 | 20.54 |
| Hemoglobin Concentration | 56/536 | 7.60E-04 | 0.0091 | 1.64 | 11.77 |
| COVID-19 (Covid Pneumonia Vs Population) | 9/38 | 7.73E-04 | 0.0092 | 4.31 | 30.88 |
| Prudent Dietary Pattern | 7/24 | 7.84E-04 | 0.0093 | 5.71 | 40.85 |
| Renal Cell Carcinoma | 7/24 | 7.84E-04 | 0.0093 | 5.71 | 40.85 |
| Asparaginase Hypersensitivity In Acute Lymphoblastic Leukemia | 7/24 | 7.84E-04 | 0.0093 | 5.71 | 40.85 |
| Verbal-numerical Reasoning | 12/62 | 7.86E-04 | 0.0093 | 3.34 | 23.85 |
| Bone Mineral Density (Femoral Neck) | 8/31 | 8.21E-04 | 0.0096 | 4.83 | 34.30 |
| Tinnitus-related Distress | 8/31 | 8.21E-04 | 0.0096 | 4.83 | 34.30 |
| Leprosy | 10/46 | 8.25E-04 | 0.0096 | 3.86 | 27.40 |
| Pursuit Maintenance Gain | 10/46 | 8.25E-04 | 0.0096 | 3.86 | 27.40 |
| Suicide Attempts | 6/18 | 8.57E-04 | 0.0099 | 6.93 | 48.97 |
| Vaginal Microbiome MetaCyc Pathway (PWY-6562 \| norspermidine Biosynthesis) | 6/18 | 8.57E-04 | 0.0099 | 6.93 | 48.97 |
| A Body Shape Index | 45/410 | 8.93E-04 | 0.0103 | 1.73 | 12.13 |
| Orofacial Clefts | 9/39 | 9.46E-04 | 0.0109 | 4.17 | 29.01 |

| | | | | | |
|---|---|---|---|---|---|
| High Myopia | 15/90 | 9.64E-04 | 0.0110 | 2.78 | 19.33 |
| Visceral Fat | 11/55 | 9.67E-04 | 0.0110 | 3.47 | 24.11 |
| Bioavailable Testosterone Levels | 36/309 | 9.83E-04 | 0.0112 | 1.84 | 12.77 |
| Alzheimer's Disease (Cognitive Decline) | 10/47 | 9.86E-04 | 0.0112 | 3.75 | 25.99 |
| Birth Weight | 24/179 | 9.93E-04 | 0.0112 | 2.16 | 14.93 |
| Loneliness | 7/25 | 0.0010 | 0.0115 | 5.40 | 37.13 |
| Coronary Artery Aneurysm In Kawasaki Disease | 7/25 | 0.0010 | 0.0115 | 5.40 | 37.13 |
| Migraine - Clinic-Based | 7/25 | 0.0010 | 0.0115 | 5.40 | 37.13 |
| Age Of Smoking Initiation | 8/32 | 0.0010 | 0.0115 | 4.63 | 31.82 |
| Acceptance Of An Invitation To Participate In A Mental Health Questionnaire | 8/32 | 0.0010 | 0.0115 | 4.63 | 31.82 |
| Total Testosterone Levels | 45/413 | 0.0010 | 0.0115 | 1.71 | 11.77 |
| Metabolic Biomarkers (Multivariate Analysis) | 24/180 | 0.0011 | 0.0119 | 2.15 | 14.67 |
| Pelvic Organ Prolapse (Moderate/Severe) | 5/13 | 0.0011 | 0.0123 | 8.66 | 58.76 |
| Vaginal Microbiome MetaCyc Pathway (PWY-6397 \| mycolyl-arabinogalactan-peptidoglycan Complex Biosynthesis) | 5/13 | 0.0011 | 0.0123 | 8.66 | 58.76 |
| Endometrial Cancer | 9/40 | 0.0011 | 0.0123 | 4.03 | 27.29 |
| Bipolar Disorder Or Body Mass Index | 4/8 | 0.0012 | 0.0123 | 13.85 | 93.63 |
| Gut Microbiota Relative Abundance (Unclassified Genus Belonging To Family Ruminococcaceae) | 4/8 | 0.0012 | 0.0123 | 13.85 | 93.63 |
| Autism Spectrum Disorder (MTAG) | 4/8 | 0.0012 | 0.0123 | 13.85 | 93.63 |
| QT Interval (Drug Interaction) | 4/8 | 0.0012 | 0.0123 | 13.85 | 93.63 |
| Joint Destruction In Rheumatoid Arthritis (Rapid Vs Slow) | 4/8 | 0.0012 | 0.0123 | 13.85 | 93.63 |
| Response To Chemotherapy In Breast Cancer Hypertensive Cases (Cumulative Dose) (Bevacizumab) | 4/8 | 0.0012 | 0.0123 | 13.85 | 93.63 |
| Facial Morphology (Factor 4, Facial Height Related To Vertical Position Of Gnathion) | 4/8 | 0.0012 | 0.0123 | 13.85 | 93.63 |
| Dihexosylceramide (D16:1/16:0) Levels | 4/8 | 0.0012 | 0.0123 | 13.85 | 93.63 |
| Glycosphingolipids (D42:1) Levels | 4/8 | 0.0012 | 0.0123 | 13.85 | 93.63 |
| Vaginal Microbiome MetaCyc Pathway (GLUCOSE1PMETAB-PWY \| glucose And Glucose-1-Phosphate Degradation) | 4/8 | 0.0012 | 0.0123 | 13.85 | 93.63 |
| F-salty Food Liking (Derived Food-Liking Factor) | 4/8 | 0.0012 | 0.0123 | 13.85 | 93.63 |
| Serum Iron Levels | 4/8 | 0.0012 | 0.0123 | 13.85 | 93.63 |

| | | | | | |
|---|---|---|---|---|---|
| 1,2-dipalmitoyl-GPE (16:0/16:0) Levels In Elite Athletes | 4/8 | 0.0012 | 0.0123 | 13.85 | 93.63 |
| Brain Region Volumes | 27/213 | 0.0012 | 0.0124 | 2.03 | 13.66 |
| Response To Paliperidone In Schizophrenia (Negative Marder Score) | 8/33 | 0.0013 | 0.0134 | 4.44 | 29.58 |
| Central Corneal Thickness | 12/66 | 0.0014 | 0.0146 | 3.09 | 20.30 |
| Opioid Addiction | 12/66 | 0.0014 | 0.0146 | 3.09 | 20.30 |
| Pancreatic Cancer | 13/75 | 0.0014 | 0.0148 | 2.92 | 19.11 |
| Positive Affect | 17/113 | 0.0015 | 0.0155 | 2.46 | 16.04 |
| Edge-level Brain Connectivity (Multivariate Analysis) | 19/133 | 0.0015 | 0.0158 | 2.32 | 15.06 |
| S-6-hydroxywarfarin To S-warfarin Ratio | 8/34 | 0.0016 | 0.0163 | 4.27 | 27.55 |
| Exacerbations Requiring Hospitalisation In Asthma | 6/20 | 0.0016 | 0.0163 | 5.94 | 38.29 |
| Urate Levels In Obese Individuals | 6/20 | 0.0016 | 0.0163 | 5.94 | 38.29 |
| Femur Total Bone Mineral Density X Gut Microbiota (?Genus Lactococcus) Interaction | 6/20 | 0.0016 | 0.0163 | 5.94 | 38.29 |
| COVID-19 (Covid Vs Negative) | 20/144 | 0.0016 | 0.0168 | 2.25 | 14.40 |
| Response To Antidepressants (Symptom Improvement) | 5/14 | 0.0017 | 0.0168 | 7.70 | 49.27 |
| Adipsin Levels | 5/14 | 0.0017 | 0.0168 | 7.70 | 49.27 |
| Upper Eyelid Sagging Severity | 5/14 | 0.0017 | 0.0168 | 7.70 | 49.27 |
| F-fish Liking (Derived Food-Liking Factor) | 5/14 | 0.0017 | 0.0168 | 7.70 | 49.27 |
| Whole Brain Free Water Diffusion (Multivariate Analysis) | 18/124 | 0.0017 | 0.0168 | 2.36 | 15.12 |
| Pulmonary Function Decline | 7/27 | 0.0017 | 0.0168 | 4.86 | 31.01 |
| Photic Sneeze Reflex | 7/27 | 0.0017 | 0.0168 | 4.86 | 31.01 |
| Calcium Levels | 35/308 | 0.0018 | 0.0176 | 1.79 | 11.36 |
| Coronary Artery Disease | 52/508 | 0.0018 | 0.0179 | 1.60 | 10.11 |
| Smoking Status | 17/115 | 0.0018 | 0.0180 | 2.41 | 15.24 |
| Anorexia Nervosa, Attention-Deficit/Hyperactivity Disorder, Autism Spectrum Disorder, Bipolar Disorder, Major Depression, Obsessive-Compulsive Disorder, Schizophrenia, Or Tourette Syndrome (Pleiotropy) | 15/96 | 0.0019 | 0.0187 | 2.58 | 16.16 |
| Neurofibrillary Tangles | 8/35 | 0.0019 | 0.0189 | 4.11 | 25.71 |
| Velopharyngeal Dysfunction | 9/43 | 0.0020 | 0.0189 | 3.67 | 22.89 |
| Gut Microbiota Relative Abundance (Ruminococcus Belonging To Family Erysipelotrichaceae) | 4/9 | 0.0020 | 0.0189 | 11.08 | 68.99 |
| Caudal Anterior-Cingulate Cortex Volume | 4/9 | 0.0020 | 0.0189 | 11.08 | 68.99 |

| | | | | | |
|---|---|---|---|---|---|
| Stress Sensitivity (Neuroticism Score X Major Depressive Disorder Status Interaction) | 4/9 | 0.0020 | 0.0189 | 11.08 | 68.99 |
| Lewy Body Disease | 4/9 | 0.0020 | 0.0189 | 11.08 | 68.99 |
| Tumor Necrosis Factor Alpha Levels | 4/9 | 0.0020 | 0.0189 | 11.08 | 68.99 |
| Heart Rate Response To Beta Blockers (Atenolol Add-On Therapy) | 4/9 | 0.0020 | 0.0189 | 11.08 | 68.99 |
| Docetaxel-induced Peripheral Neuropathy In Metastatic Castrate-Resistant Prostate Cancer | 4/9 | 0.0020 | 0.0189 | 11.08 | 68.99 |
| Serum IgE Levels | 4/9 | 0.0020 | 0.0189 | 11.08 | 68.99 |
| Vaginal Microbiome MetaCyc Pathway (PWY-5837 \| 1,4-dihydroxy-2-naphthoate Biosynthesis I) | 4/9 | 0.0020 | 0.0189 | 11.08 | 68.99 |
| COVID-19 (Recovered Vs Asymptomatic) | 4/9 | 0.0020 | 0.0189 | 11.08 | 68.99 |
| Choline Phosphate Levels In Elite Athletes | 4/9 | 0.0020 | 0.0189 | 11.08 | 68.99 |
| Atrial Fibrillation/Atrial Flutter | 11/60 | 0.0020 | 0.0194 | 3.12 | 19.33 |
| Body Fat Distribution (Trunk Fat Ratio) | 13/78 | 0.0021 | 0.0195 | 2.78 | 17.21 |
| Serum Platinum Levels After Completion Of Cisplatin Chemotherapy | 6/21 | 0.0021 | 0.0198 | 5.55 | 34.19 |
| Antipsychotic Drug-Induced QTc Interval Change In Schizophrenia | 6/21 | 0.0021 | 0.0198 | 5.55 | 34.19 |
| Selective IgA Deficiency | 6/21 | 0.0021 | 0.0198 | 5.55 | 34.19 |
| Epstein-Barr Virus Copy Number In Lymphoblastoid Cell Lines | 7/28 | 0.0021 | 0.0199 | 4.62 | 28.47 |
| Arterial Stiffness Index | 7/28 | 0.0021 | 0.0199 | 4.62 | 28.47 |
| Malaria | 8/36 | 0.0023 | 0.0217 | 3.96 | 24.03 |
| COVID-19 (Severe Vs Tested, Not Severe) | 9/44 | 0.0023 | 0.0217 | 3.57 | 21.63 |
| Gut Microbiome Composition (Winter) | 5/15 | 0.0024 | 0.0217 | 6.93 | 41.92 |
| Human Papilloma Virus 16 Negative Oropharyngeal Cancer | 5/15 | 0.0024 | 0.0217 | 6.93 | 41.92 |
| HOMA-B (Corrected For HOMA-IR) | 5/15 | 0.0024 | 0.0217 | 6.93 | 41.92 |
| Migraine With Aura | 5/15 | 0.0024 | 0.0217 | 6.93 | 41.92 |
| Shellfish Liking | 5/15 | 0.0024 | 0.0217 | 6.93 | 41.92 |
| Headache Or Type 2 Diabetes | 5/15 | 0.0024 | 0.0217 | 6.93 | 41.92 |
| Cognitive Function (Immediate Memory) (Longitudinal) | 7/29 | 0.0026 | 0.0239 | 4.41 | 26.22 |
| Nicotine Dependence And Major Depression (Severity Of Comorbidity) | 7/29 | 0.0026 | 0.0239 | 4.41 | 26.22 |
| Asthma | 34/304 | 0.0026 | 0.0239 | 1.76 | 10.44 |
| Gut Microbiota (Bacterial Taxa, Rank Normal Transformation Method) | 12/71 | 0.0027 | 0.0239 | 2.83 | 16.75 |
| Nasal Polyps | 6/22 | 0.0027 | 0.0239 | 5.20 | 30.70 |

| | | | | | |
|---|---|---|---|---|---|
| Gut Microbiome Composition (Summer) | 6/22 | 0.0027 | 0.0239 | 5.20 | 30.70 |
| Vaginal Microbiome MetaCyc Pathway (CATECHOL-ORTHO-CLEAVAGE-PWY\|catechol Degradation To Β-Ketoadipate) | 6/22 | 0.0027 | 0.0239 | 5.20 | 30.70 |
| Cholesterol Levels In Medium VLDL | 9/45 | 0.0027 | 0.0239 | 3.47 | 20.46 |
| Concentration Of Large LDL Particles | 9/45 | 0.0027 | 0.0239 | 3.47 | 20.46 |
| Cytokine Levels | 3/5 | 0.0028 | 0.0239 | 20.77 | 122.31 |
| Sleep Duration (> 10 Hours) | 3/5 | 0.0028 | 0.0239 | 20.77 | 122.31 |
| Anxiety In Major Depressive Disorder | 3/5 | 0.0028 | 0.0239 | 20.77 | 122.31 |
| Midgestational Circulating Levels Of PBDEs | 3/5 | 0.0028 | 0.0239 | 20.77 | 122.31 |
| Residual Cognition | 3/5 | 0.0028 | 0.0239 | 20.77 | 122.31 |
| Ceramide (D17:1/18:0) Levels | 3/5 | 0.0028 | 0.0239 | 20.77 | 122.31 |
| Ceramide (D19:1/26:0) Levels | 3/5 | 0.0028 | 0.0239 | 20.77 | 122.31 |
| Dihydroorotate Levels In Elite Athletes | 3/5 | 0.0028 | 0.0239 | 20.77 | 122.31 |
| Indoleacetate Levels In Elite Athletes | 3/5 | 0.0028 | 0.0239 | 20.77 | 122.31 |
| Ceramide (D17:1/20:0) Levels | 3/5 | 0.0028 | 0.0239 | 20.77 | 122.31 |
| Sphingomyelin (D16:1/23:0) levels/Sphingomyelin (D17:1/22:0) Levels | 3/5 | 0.0028 | 0.0239 | 20.77 | 122.31 |
| Sphingomyelin (D17:1/16:0) Levels | 3/5 | 0.0028 | 0.0239 | 20.77 | 122.31 |
| Neck Circumference | 3/5 | 0.0028 | 0.0239 | 20.77 | 122.31 |
| 4-Vinylphenol Sulfate Levels In Elite Athletes | 3/5 | 0.0028 | 0.0239 | 20.77 | 122.31 |
| Gamma-glutamylphenylalanine Levels In Elite Athletes | 3/5 | 0.0028 | 0.0239 | 20.77 | 122.31 |
| 2-stearoyl-GPE (18:0) Levels In Elite Athletes | 3/5 | 0.0028 | 0.0239 | 20.77 | 122.31 |
| Daytime Sleep Phenotypes | 8/37 | 0.0028 | 0.0241 | 3.83 | 22.50 |
| Cardioembolic Stroke (MTAG) | 8/37 | 0.0028 | 0.0241 | 3.83 | 22.50 |
| Asthma (Adult Onset) | 10/54 | 0.0030 | 0.0255 | 3.16 | 18.37 |
| Chronic Lymphocytic Leukemia | 12/72 | 0.0030 | 0.0257 | 2.78 | 16.14 |
| Post Bronchodilator FEV1/FVC Ratio In COPD | 11/63 | 0.0030 | 0.0259 | 2.94 | 17.03 |
| Cannabis Use (Initiation) | 4/10 | 0.0031 | 0.0262 | 9.23 | 53.28 |
| Response To Cognitive-Behavioural Therapy In Anxiety Disorder | 4/10 | 0.0031 | 0.0262 | 9.23 | 53.28 |
| Aggressiveness In Attention Deficit Hyperactivity Disorder | 4/10 | 0.0031 | 0.0262 | 9.23 | 53.28 |
| Gestational Age At Birth In Labor-Initiated Deliveries (Child Effect) | 4/10 | 0.0031 | 0.0262 | 9.23 | 53.28 |
| Response To Cytidine Analogues (Gemcitabine) | 4/10 | 0.0031 | 0.0262 | 9.23 | 53.28 |
| Glycosphingolipids (D41:1) Levels | 4/10 | 0.0031 | 0.0262 | 9.23 | 53.28 |

| | | | | | |
|---|---|---|---|---|---|
| Vaginal Microbiome MetaCyc Pathway (PWY-5861 \| superpathway Of Demethylmenaquinol-8 Biosynthesis) | 4/10 | 0.0031 | 0.0262 | 9.23 | 53.28 |
| Vaginal Microbiome MetaCyc Pathway (PWY-5863 \| superpathway Of Phylloquinol Biosynthesis) | 4/10 | 0.0031 | 0.0262 | 9.23 | 53.28 |
| Neurociticism | 15/101 | 0.0031 | 0.0262 | 2.43 | 13.99 |
| Stem Cell Growth Factor Beta Levels | 7/30 | 0.0032 | 0.0267 | 4.22 | 24.21 |
| Attention Deficit Hyperactivity Disorder (Inattention Symptoms) | 5/16 | 0.0032 | 0.0267 | 6.30 | 36.11 |
| Lung Cancer In Never Smokers | 5/16 | 0.0032 | 0.0267 | 6.30 | 36.11 |
| Facial Morphology (Factor 19) | 5/16 | 0.0032 | 0.0267 | 6.30 | 36.11 |
| Caudate Activity During Reward | 5/16 | 0.0032 | 0.0267 | 6.30 | 36.11 |
| Strabismus | 5/16 | 0.0032 | 0.0267 | 6.30 | 36.11 |
| Hip Circumference Adjusted For BMI | 82/900 | 0.0033 | 0.0274 | 1.41 | 8.04 |
| Cholesteryl Ester Levels In Small LDL | 8/38 | 0.0033 | 0.0274 | 3.70 | 21.10 |
| Depressive Symptoms (MTAG) | 12/73 | 0.0034 | 0.0277 | 2.73 | 15.55 |
| High-sensitivity Cardiac Troponin T Levels | 10/55 | 0.0034 | 0.0279 | 3.09 | 17.53 |
| Cerebrospinal Fluid AB1-42 Levels | 6/23 | 0.0035 | 0.0282 | 4.89 | 27.69 |
| Smooth-surface Caries | 6/23 | 0.0035 | 0.0282 | 4.89 | 27.69 |
| Bipolar Disorder (Body Mass Index Interaction) | 6/23 | 0.0035 | 0.0282 | 4.89 | 27.69 |
| Alzheimer's Disease In non-APOE E4 Carriers | 6/23 | 0.0035 | 0.0282 | 4.89 | 27.69 |
| Age At First Birth | 9/47 | 0.0037 | 0.0302 | 3.29 | 18.37 |
| COVID-19 (Severe Vs Population) | 9/47 | 0.0037 | 0.0302 | 3.29 | 18.37 |
| Optic Disc Size | 12/74 | 0.0038 | 0.0305 | 2.69 | 14.99 |
| Waist Circumference Adjusted For BMI (Adjusted For Smoking Behaviour) | 10/56 | 0.0039 | 0.0313 | 3.02 | 16.74 |
| Spatial QRS-T Angle | 10/56 | 0.0039 | 0.0313 | 3.02 | 16.74 |
| Phospholipids To Total Lipids Ratio In Small HDL | 8/39 | 0.0040 | 0.0317 | 3.58 | 19.81 |
| Serum Uric Acid Levels | 32/290 | 0.0042 | 0.0339 | 1.73 | 9.45 |
| COVID-19 (Covid Respiratory Support Vs Population) | 9/48 | 0.0043 | 0.0341 | 3.20 | 17.43 |
| Coenzyme Q10 Levels | 5/17 | 0.0043 | 0.0341 | 5.77 | 31.41 |
| Spherical Equivalent (Joint Analysis Main Effects And Education Interaction) | 5/17 | 0.0043 | 0.0341 | 5.77 | 31.41 |
| Frontal Fibrosing Alopecia | 5/17 | 0.0043 | 0.0341 | 5.77 | 31.41 |
| Alcohol Dependence (Age At Onset) | 5/17 | 0.0043 | 0.0341 | 5.77 | 31.41 |
| F-seafood Liking (Derived Food-Liking Factor) | 5/17 | 0.0043 | 0.0341 | 5.77 | 31.41 |
| Vaginal Microbiome Relative Abundance (S Aerococcus Christensenii) | 5/17 | 0.0043 | 0.0341 | 5.77 | 31.41 |

| | | | | | |
|---|---|---|---|---|---|
| Emphysema Annual Change Measurement In Smokers (Adjusted Lung Density) | 6/24 | 0.0044 | 0.0342 | 4.62 | 25.09 |
| Mononucleosis | 6/24 | 0.0044 | 0.0342 | 4.62 | 25.09 |
| Depression Severity X Hours Spent Watching Television Interaction | 6/24 | 0.0044 | 0.0342 | 4.62 | 25.09 |
| Hyperopia | 10/57 | 0.0045 | 0.0347 | 2.95 | 15.99 |
| Response To Norepinephrine-Dopamine Reuptake Inhibitors (Responders Vs Non-Responders) | 4/11 | 0.0046 | 0.0355 | 7.91 | 42.52 |
| Gut Microbiota Relative Abundance (Faecalibacterium) | 4/11 | 0.0046 | 0.0355 | 7.91 | 42.52 |
| Medial Orbital Frontal Cortex Volume | 4/11 | 0.0046 | 0.0355 | 7.91 | 42.52 |
| Oppositional Defiant Disorder Dimensions In Attention-Deficit Hyperactivity Disorder | 4/11 | 0.0046 | 0.0355 | 7.91 | 42.52 |
| Alcohol Consumption (Max-Drinks) | 4/11 | 0.0046 | 0.0355 | 7.91 | 42.52 |
| Right Hippocampal Volume | 4/11 | 0.0046 | 0.0355 | 7.91 | 42.52 |
| Vaginal Microbiome MetaCyc Pathway (PWY-7371 \| 1,4-dihydroxy-6-naphthoate Biosynthesis II) | 4/11 | 0.0046 | 0.0355 | 7.91 | 42.52 |
| Vaginal Microbiome MetaCyc Pathway (PWY1G-0 \| mycothiol Biosynthesis) | 4/11 | 0.0046 | 0.0355 | 7.91 | 42.52 |
| Unilateral Cleft Lip | 4/11 | 0.0046 | 0.0355 | 7.91 | 42.52 |
| Osteoarthritis (Hip) | 8/40 | 0.0047 | 0.0355 | 3.47 | 18.62 |
| Migraine | 17/126 | 0.0048 | 0.0364 | 2.17 | 11.59 |
| Alzheimer Disease And Age Of Onset | 14/96 | 0.0050 | 0.0379 | 2.37 | 12.57 |
| Itch Intensity From Mosquito Bite Adjusted By Bite Size | 17/127 | 0.0052 | 0.0379 | 2.15 | 11.31 |
| Chronic Rhinosinusitis | 3/6 | 0.0053 | 0.0379 | 13.84 | 72.65 |
| Non-del(5q) Myelodysplastic Syndromes | 3/6 | 0.0053 | 0.0379 | 13.84 | 72.65 |
| Autism And Major Depressive Disorder (MTAG) | 3/6 | 0.0053 | 0.0379 | 13.84 | 72.65 |
| Crohn's Disease (Time To Progression) | 3/6 | 0.0053 | 0.0379 | 13.84 | 72.65 |
| Evening Vs. Morning Chronotype (sMEQ Score) | 3/6 | 0.0053 | 0.0379 | 13.84 | 72.65 |
| Parkinson's Disease Progression (Motor) | 3/6 | 0.0053 | 0.0379 | 13.84 | 72.65 |
| Caudate Volume Change Rate X Age Interaction (1Df) | 3/6 | 0.0053 | 0.0379 | 13.84 | 72.65 |
| Postprandial Triglyceride Response | 3/6 | 0.0053 | 0.0379 | 13.84 | 72.65 |
| Radiation Response | 3/6 | 0.0053 | 0.0379 | 13.84 | 72.65 |
| N6,N6-dimethyllysine Levels | 3/6 | 0.0053 | 0.0379 | 13.84 | 72.65 |
| Ceramide (D16:1/24:0) Levels | 3/6 | 0.0053 | 0.0379 | 13.84 | 72.65 |
| Ceramide (D16:1/24:1) Levels | 3/6 | 0.0053 | 0.0379 | 13.84 | 72.65 |
| Ceramide (D16:1/22:0) Levels | 3/6 | 0.0053 | 0.0379 | 13.84 | 72.65 |
| Ceramide (D16:1/18:0) Levels | 3/6 | 0.0053 | 0.0379 | 13.84 | 72.65 |

| | | | | | |
|---|---|---|---|---|---|
| P-cresol Sulfate Levels In Elite Athletes | 3/6 | 0.0053 | 0.0379 | 13.84 | 72.65 |
| Sphingomyelin(39:1) [M+H]1+ Levels | 3/6 | 0.0053 | 0.0379 | 13.84 | 72.65 |
| Sphingomyelin (D18:1/14:0) levels/Sphingomyelin (D16:1/16:0) Levels | 3/6 | 0.0053 | 0.0379 | 13.84 | 72.65 |
| Vaginal Microbiome MetaCyc Pathway (FASYN-INITIAL-PWY \| superpathway Of Fatty Acid Biosynthesis Initiation (E. Coli)) | 3/6 | 0.0053 | 0.0379 | 13.84 | 72.65 |
| Vaginal Microbiome MetaCyc Pathway (PWY0-1241 \| ADP-L-glycero-β-D-manno-heptose Biosynthesis) | 3/6 | 0.0053 | 0.0379 | 13.84 | 72.65 |
| Time To Recovery From Peripheral Sensory Neuropathy In Colon Cancer | 3/6 | 0.0053 | 0.0379 | 13.84 | 72.65 |
| Vaginal Microbiome Relative Abundance (O Campylobacterales) | 3/6 | 0.0053 | 0.0379 | 13.84 | 72.65 |
| Angiopoietin-1 Receptor Levels | 3/6 | 0.0053 | 0.0379 | 13.84 | 72.65 |
| Plasma Selenium Levels | 3/6 | 0.0053 | 0.0379 | 13.84 | 72.65 |
| Immunoglobulin Lambda-Like Polypeptide 1 Levels | 3/6 | 0.0053 | 0.0379 | 13.84 | 72.65 |
| Multiple Sclerosis | 30/271 | 0.0053 | 0.0379 | 1.74 | 9.11 |
| Nicotine Dependence Symptom Count | 13/87 | 0.0054 | 0.0390 | 2.44 | 12.73 |
| Emphysema Imaging Phenotypes | 6/25 | 0.0054 | 0.0390 | 4.38 | 22.82 |
| Tea Intake (UKB Data Field 1488) | 6/25 | 0.0054 | 0.0390 | 4.38 | 22.82 |
| S-warfarin Levels | 8/41 | 0.0055 | 0.0390 | 3.36 | 17.53 |
| Gut Microbiota Alpha Diversity (Shannon Index) | 5/18 | 0.0057 | 0.0399 | 5.33 | 27.56 |
| Osteoarthritis (Self-Reported) | 5/18 | 0.0057 | 0.0399 | 5.33 | 27.56 |
| Familial Lung Adenocarcinoma | 5/18 | 0.0057 | 0.0399 | 5.33 | 27.56 |
| Aortic Root Size | 5/18 | 0.0057 | 0.0399 | 5.33 | 27.56 |
| Blood Pressure (Smoking Interaction) | 5/18 | 0.0057 | 0.0399 | 5.33 | 27.56 |
| Bronchopulmonary Dysplasia | 5/18 | 0.0057 | 0.0399 | 5.33 | 27.56 |
| Hepatitis A | 5/18 | 0.0057 | 0.0399 | 5.33 | 27.56 |
| Blood Osmolality (Transformed Sodium) | 5/18 | 0.0057 | 0.0399 | 5.33 | 27.56 |
| Corneal Curvature | 7/33 | 0.0057 | 0.0399 | 3.73 | 19.30 |
| Mean Arterial Pressure | 29/261 | 0.0057 | 0.0400 | 1.74 | 9.00 |
| Total Protein Levels X Insomnia Interaction | 10/59 | 0.0057 | 0.0402 | 2.83 | 14.62 |
| Oily Fish Consumption | 8/42 | 0.0063 | 0.0442 | 3.26 | 16.51 |
| Bone Stiffness Index | 8/42 | 0.0063 | 0.0442 | 3.26 | 16.51 |
| BMI In Smokers | 4/12 | 0.0066 | 0.0453 | 6.92 | 34.77 |
| Toxicity Response To Radiotherapy In Prostate Cancer (Decreased Urine Stream) (Time To Event) | 4/12 | 0.0066 | 0.0453 | 6.92 | 34.77 |
| Thoracic Aortic Aneurysms | 4/12 | 0.0066 | 0.0453 | 6.92 | 34.77 |
| Facial Morphology (Factor 6, Height Of Vermillion Lower Lip) | 4/12 | 0.0066 | 0.0453 | 6.92 | 34.77 |

| | | | | | |
|---|---|---|---|---|---|
| Facial Morphology (Factor 16) | 4/12 | 0.0066 | 0.0453 | 6.92 | 34.77 |
| Vaginal Microbiome MetaCyc Pathway (PWY-5747\|2-methylcitrate Cycle II) | 4/12 | 0.0066 | 0.0453 | 6.92 | 34.77 |
| Horseradish Liking | 4/12 | 0.0066 | 0.0453 | 6.92 | 34.77 |
| Congenital Solitary Functioning Kidney | 4/12 | 0.0066 | 0.0453 | 6.92 | 34.77 |
| Post-traumatic Stress Disorder | 6/26 | 0.0067 | 0.0458 | 4.16 | 20.83 |
| Aerodigestive Squamous Cell Cancer (Pleiotropy) | 15/110 | 0.0070 | 0.0481 | 2.19 | 10.88 |
| Adipose Morphology | 5/19 | 0.0073 | 0.0494 | 4.95 | 24.36 |
| Total Cerebellar Volume (Excluding Crus I Vermis) | 5/19 | 0.0073 | 0.0494 | 4.95 | 24.36 |
| Unilateral Cleft Lip And Palate | 5/19 | 0.0073 | 0.0494 | 4.95 | 24.36 |
| Bisphosphonate-associated Atypical Femoral Fracture | 5/19 | 0.0073 | 0.0494 | 4.95 | 24.36 |
| Takayasu Arteritis | 10/61 | 0.0073 | 0.0494 | 2.72 | 13.39 |
| Body Fat Mass | 8/43 | 0.0073 | 0.0496 | 3.17 | 15.58 |
| Household Income (MTAG) | 19/153 | 0.0073 | 0.0496 | 1.97 | 9.69 |
| Tea Consumption | 9/52 | 0.0075 | 0.0503 | 2.90 | 14.22 |
| Uterine Fibroids | 12/81 | 0.0079 | 0.0532 | 2.42 | 11.68 |
