## Supplementary Table S14 for "Genetic regulation of the vascular endothelial growth factor receptor 1 during sepsis and association with ARDS susceptibility"

**Table S14. Significant results of gene set enrichment analysis on the Reactome dataset for the list of common genes obtained from the best PGS models at T1 and T7.**

| Term | Overlap | P-value | Adjusted P-value | Odds Ratio | Combined Score |
|---|---|---|---|---|---|
| Signal Transduction R-HSA-162582 | 239/2465 | 1.91E-09 | 2.62E-06 | 1.587 | 31.86 |
| Neuronal System R-HSA-112316 | 54/386 | 2.73E-07 | 1.86E-04 | 2.299 | 34.75 |
| Signaling By Receptor Tyrosine Kinases R-HSA-9006934 | 63/496 | 9.25E-07 | 4.22E-04 | 2.059 | 28.61 |
| Transcriptional Regulation Of Pluripotent Stem Cells R-HSA-452723 | 11/30 | 2.10E-06 | 7.18E-04 | 8.056 | 105.31 |
| PI3K/AKT Signaling In Cancer R-HSA- | 21/105 | 5.97E-06 | 0.002 | 3.492 | 42.01 |
| Constitutive Signaling By Aberrant PI3K In Cancer R-HSA-2219530 | 17/78 | 1.38E-05 | 0.003 | 3.886 | 43.48 |
| Protein-protein Interactions At Synapses R-HSA-6794362 | 18/88 | 1.97E-05 | 0.004 | 3.587 | 38.87 |
| Glycosaminoglycan Metabolism R-HSA- | 21/120 | 4.97E-05 | 0.008 | 2.961 | 29.34 |
| Negative Regulation Of PI3K/AKT Network R-HSA-199418 | 20/113 | 6.31E-05 | 0.010 | 3.001 | 29.02 |
| PI5P, PP2A And IER3 Regulate PI3K/AKT Signaling R-HSA-6811558 | 19/106 | 7.99E-05 | 0.010 | 3.046 | 28.74 |
| Adherens Junctions Interactions R-HSA- | 9/29 | 8.24E-05 | 0.010 | 6.252 | 58.79 |
| MECP2 Regulates Transcription Factors R-HSA-9022707 | 4/5 | 9.78E-05 | 0.010 | 55.421 | 511.68 |
| Intracellular Signaling By Second Messengers R-HSA-9006925 | 39/306 | 9.83E-05 | 0.010 | 2.048 | 18.90 |
| Defective B3GALTL Causes PpS R-HSA- | 10/37 | 1.23E-04 | 0.012 | 5.147 | 46.35 |
| O-glycosylation Of TSR Domain-Containing Proteins R-HSA-5173214 | 10/38 | 1.57E-04 | 0.014 | 4.963 | 43.48 |
| cGMP Effects R-HSA-418457 | 6/15 | 2.75E-04 | 0.024 | 9.247 | 75.80 |
| Nitric Oxide Stimulates Guanylate Cyclase R-HSA-392154 | 7/21 | 3.15E-04 | 0.025 | 6.938 | 55.94 |
| Neurexins And Neuroligins R-HSA- | 12/57 | 3.52E-04 | 0.025 | 3.708 | 29.49 |
| Signaling By ERBB4 R-HSA-1236394 | 12/57 | 3.52E-04 | 0.025 | 3.708 | 29.49 |
| Voltage Gated Potassium Channels R-HSA-1296072 | 10/43 | 4.67E-04 | 0.032 | 4.210 | 32.29 |
| Developmental Biology R-HSA-1266738 | 100/1073 | 5.95E-04 | 0.038 | 1.453 | 10.80 |
| Gastrulation R-HSA-9758941 | 6/17 | 6.06E-04 | 0.038 | 7.565 | 56.05 |
| Cell-cell Junction Organization R-HSA-421270 | 12/61 | 6.75E-04 | 0.040 | 3.405 | 24.86 |
| Nuclear Signaling By ERBB4 R-HSA-1251985 | 8/32 | 0.00103 | 0.057 | 4.626 | 31.82 |
